# Association between phthalate exposure and early puberty: An updated meta-analysis exploring sex and exposure time variations

**DOI:** 10.1101/2025.06.26.25330269

**Authors:** Hsuan-Tung Lee, Yen-Ting Kuo, Uyanga Munkhbayar, Yi-No Kang, Yang-Ching Chen

## Abstract

Puberty is a critical developmental stage, and its early onset, termed precocious puberty, has garnered attention because of its potential health and psychosocial implications. Environmental exposure to endocrine-disrupting chemicals (EDCs), including phthalates, has been associated with alterations in pubertal timing; however, the evidence remains inconsistent. This meta-analysis investigated the association between exposure to specific phthalate metabolites and early puberty; the study focused on the effects of sex and exposure timing (prenatal vs. postnatal).

We conducted a systematic search to identify studies examining phthalate exposure and early puberty, covering research published between 2014 and 2024. This search yielded 29 relevant studies, of which 13 met our inclusion criteria and were selected for analysis. A previously published meta-analysis had synthesized data from these same 13 studies, aligning with our research objectives. Due to reporting heterogeneity across the additional studies, our analysis focused on these 13 studies. Random-effects models were applied to estimate pooled relative risks (RRs), with data stratified by sex and exposure timing. Meta-regression and subgroup analyses were performed to evaluate the effects of demographic and temporal factors.

The results indicated limited associations between most phthalates and early puberty risk. Postnatal exposure to mono-n-butyl phthalate was modestly associated with an increased risk of early puberty in boys (RR: 1.03; 95% confidence interval [CI]: 1.01–1.06), whereas exposure to mono-(3-carboxypropyl) phthalate was associated with a slight reduction in the risk of early puberty in girls (RR: 0.955; 95% CI: 0.917–0.995). However, these associations were not significant after adjustment for urine specific gravity, suggesting the presence of measurement variability or residual confounding. Subgroup and meta-regression analyses revealed no significant modifying effects of sex or exposure timing. Publication bias assessments indicated no substantial asymmetry.

Despite these observed patterns, definitive conclusions cannot be drawn because of data limitations, small sample sizes, and methodological inconsistencies. Future research should prioritize standardized exposure and outcome reporting, larger cohorts, and investigation into the cumulative effects of phthalates with other EDCs. This study demonstrates the complexity of the effects of phthalates on pubertal timing and the need for more rigorous investigations to guide public health interventions.

## Introduction

The onset of puberty marks a critical stage in human development, and it has critical implications for physical, psychological, and social well-being. Over the past few decades, concerns have been growing regarding the increasing prevalence of precocious puberty, which is characterized by the early onset of secondary sexual characteristics ^1^. Emerging evidence indicates that environmental factors, particularly exposure to endocrine-disrupting chemicals (EDCs), may play a pivotal role in this trend ^2^. Among EDCs, phthalates—a class of synthetic chemicals widely used in plastics, personal care products, and industrial applications—have attracted considerable attention owing to their potential to disrupt hormonal regulation ^3^.

Phthalates disrupt the hypothalamic–pituitary–gonadal (HPG) axis through various mechanisms, including the modulation of androgen and estrogen signaling ^3^. Although phthalates have attracted extensive research attention, evidence regarding their role in precocious or early puberty remains inconclusive ^4–9^. Some studies have reported significant associations between exposure to certain phthalate metabolites and earlier pubertal onset, whereas others have observed no such association. These discrepancies may be due to variations in study design, population characteristics, and phthalate exposure timing or levels. Furthermore, some studies have investigated the potential modifying effects of sex and exposure timing (prenatal vs. postnatal), but the overall evidence remains weak and often conflicting ^9–15^.

To address these gaps, we conducted a meta-analysis focusing on individual phthalates instead of combined measures. In particular, this meta-analysis examined sex-specific differences and the effects of exposure timing in order to provide a deeper understanding of the association between phthalate exposure and early puberty.

## Methods

### Protocol and registration

This review was conducted in accordance with the Preferred Reporting Items for Systematic Reviews and Meta-Analysis (PRISMA) guidelines. The protocol for this meta-analysis was prospectively registered in PROSPERO on December 21, 2024 (ID: CRD42024625792).

### Systematic search and study selection

In accordance with the PRISMA statement, we conducted a systematic search to identify studies examining the association between phthalate exposure and early puberty. We systematically searched for relevant research published in PubMed and Embase from their inception to December 2024. The search was conducted using a combination of keywords, including (“phthalate*” OR “2OH-MiBP” OR “2OH-mono-isobutyl phthalate” OR “3OH-MnBP” OR “3OH-mono-n-butyl phthalate” OR “5cx-MEPP” OR “mono(2-ethyl-5-carboxypentyl) phthalate” OR “5OH-MEHP” OR “mono(2-ethyl-5-hydroxyhexyl) phthalate” OR “5oxo-MEHP” OR “mono(2-ethyl-5-oxohexyl) phthalate” OR “cx-MiOP” OR “monocarboxy isooctyl phthalate” OR “MBzP” OR “monobenzyl phthalate” OR “MCPP” OR “mono-(3-carboxypropyl) phthalate” OR “MEHP” OR “mono(2-ethylhexyl) phthalate” OR “MEP” OR “monoethyl phthalate” OR “MiBP” OR “mono-isobutyl phthalate” OR “MMP” OR “monomethyl phthalate” OR “MnBP” OR “mono-n-butyl phthalate” OR “DEHP” OR “DBP” OR “BBP” OR “DINP” OR “DIDP” OR “DnOP” OR “EDC*” OR “endocrine disruptors”[MeSH] OR “environmental exposure”[MeSH]) AND (“precocious puberty” OR “early puberty” OR “pubertal timing” OR “menarche” OR “thelarche” OR “pubarche” OR “gonadarche” OR “testicular volume” OR “puberty”[MeSH] OR “puberty, precocious”[MeSH]).

The inclusion criteria were as follows: (1) reporting risk estimates (relative risk [RR], odds ratio [OR], and hazard ratio [HR]) for the association between exposure to individual phthalate metabolites and early puberty, (2) examining prenatal or postnatal phthalate exposure, (3) providing sufficient data for extracting effect sizes and 95% confidence intervals (CIs), and (4) assessing outcomes related to puberty timing (e.g., thelarche, menarche, or testicular volume). The exclusion criteria were as follows: (1) not being published in English, (2) not examining exposure to phthalates, (3) combining multiple phthalates into a single exposure measure, and (4) not reporting early or precocious puberty as the outcome.

Two independent reviewers screened titles and abstracts to identify potentially eligible studies. Full-text articles were assessed for inclusion on the basis of the inclusion criteria. The study selection process is illustrated in Figure 1. The systematic search yielded 29 studies. After further excluding studies with significant reporting heterogeneity (e.g., inconsistent outcomes and risk estimates, varied log scales) and those that combine multiple phthalates as a single measure, 13 studies remained.

**Figure 1.**
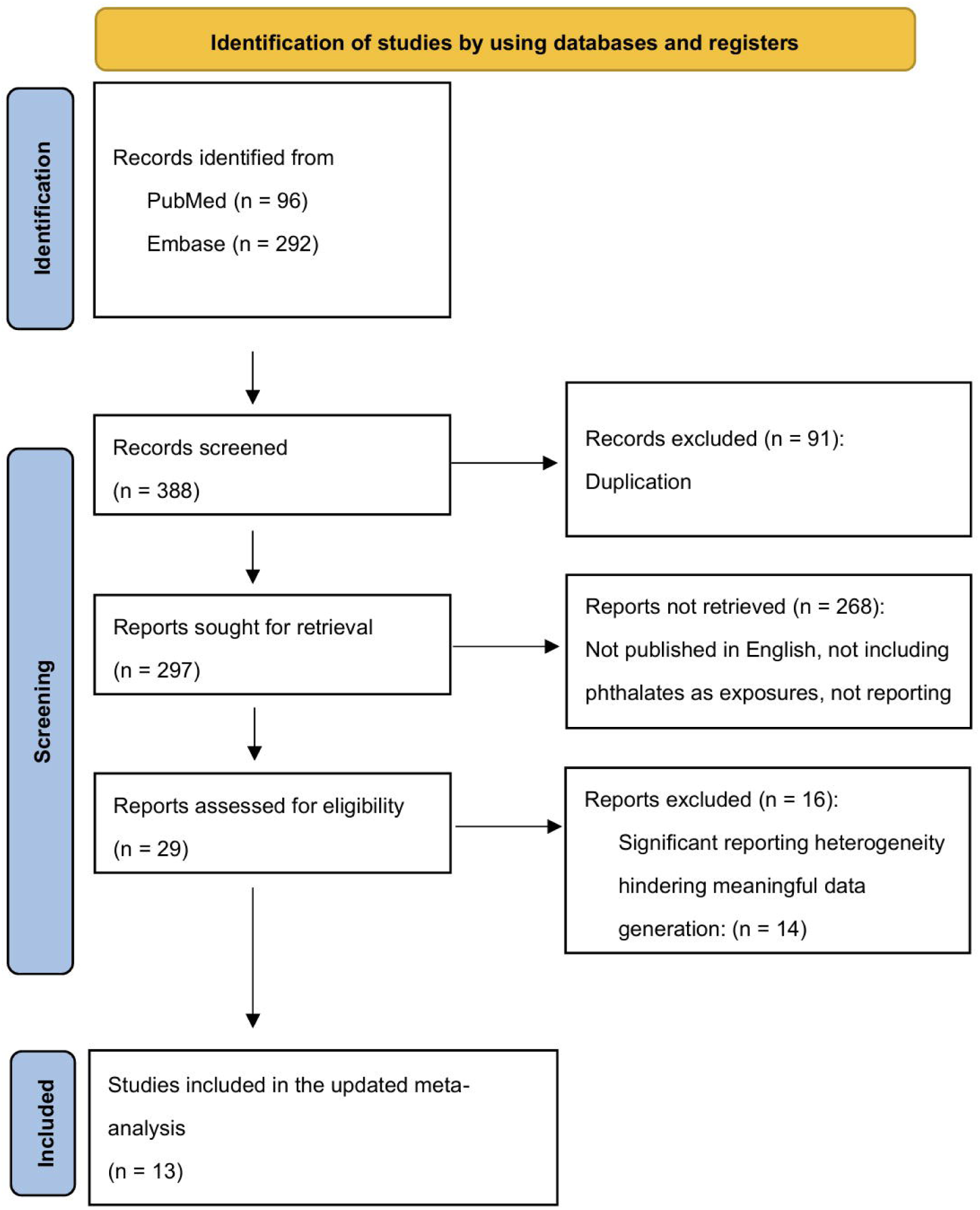
Flowchart of study selection process

After identifying 13 studies that met our inclusion criteria, we evaluated their methodological approaches and statistical measures. Given the variations in outcome reporting––such as differences in effect size measures (e.g., OR, RR, HR), adjustments for urine SG and creatinine, and log-scale transformations––we sought a standardized approach to data synthesis. A previously published comprehensive meta-analysis closely aligned with our search findings and included the same 13 studies within its dataset^9^.

This meta-analysis had already applied statistical harmonization to standardize effect sizes across studies. After carefully reviewing its methodology, we determined that its dataset provided a robust foundation for further investigation. Building upon this work, we conducted an updated meta-analysis to refine and expand upon the existing findings, with a particular focus on individual phthalates, sex-specific differences, and exposure timing.

By leveraging this established dataset, our study aims to provide a more detailed and comprehensive assessment of the association between phthalate exposure and early puberty.

### Study design

This meta-analysis examined the association between phthalate exposure and early puberty. We first conducted a systematic review and identified 13 relevant studies. After evaluating previous research, we found that a previously published meta-analysis had already harmonized these studies into a standardized dataset^9^. This meta-analysis pooled studies investigating the association between phthalate exposure and various indicators of early puberty, including thelarche and menarche in girls and testicular volume in boys. The study reported standardized risk estimates (RR, OR, and HR) as RRs on a log10 scale based on a 10-fold increase in exposure, along with corresponding 95% CIs.

For our updated analysis, we used this existing dataset as a foundation to further investigate the effects of individual phthalates, focusing on sex-specific differences and exposure timing (prenatal vs. postnatal). By analyzing the previously combined data in more detail, we aimed to provide a clearer understanding of how specific phthalates influence puberty timing.

### Data extraction and quality assessment

Three authors independently extracted data by using a predefined data extraction form. RRs and their corresponding 95% CIs were obtained from the existing dataset for each study. In addition, sample sizes for exposed groups were manually extracted from the individual studies. The phthalate levels in each study were reported as either unadjusted or adjusted for urine SG. We performed separate analyses for each group to minimize potential confounding and ensure that differences in concentration adjustments did not introduce systemic bias into the pooled estimates. Pubertal onset in girls was defined as thelarche; if thelarche data were unavailable, early menarche was used as a proxy. For boys, pubertal onset was defined as a testicular volume of >3 mL, as determined through clinical assessment.

Risk of bias was assessed using the Newcastle–Ottawa Scale (NOS) for cohort studies and an adapted version of the NOS for cross-sectional studies. For cohort studies, the NOS was applied to assess bias in three domains, namely selection, comparability, and outcome assessment, by using criteria specifically tailored to the longitudinal design of cohort studies. However, for cross-sectional studies, the adapted NOS was used to assess bias in these three domains, but the criteria were tailored to the cross-sectional study design, emphasizing the representativeness of the sample and assessment of exposure and outcomes at a single time point. In both scales, high-quality responses are assigned stars, with a maximum possible score of 9. In the NOS for cohort studies, up to two stars can be awarded for the comparability domain. For the adapted NOS for cross-sectional studies, up to two stars can be awarded for the ascertainment of the screening or surveillance tool and assessment of outcomes. Studies were categorized as high quality if they scored ≥7, moderate quality if they scored 5 or 6, and low quality if they scored <5.

### Statistical analysis

Separate meta-analyses were conducted for each phthalate, including mono-n-butyl phthalate (MnBP) and mono(2-ethylhexyl) phthalate (MEHP). RRs and 95% CIs were pooled from studies reporting data on the specific phthalate. A random-effects model was used to account for between-study heterogeneity. The Paule–Mandel estimator was employed to calculate the variance between studies (τ^2^). Heterogeneity between studies was determined using the *I*^2^ statistic, with values of <25%, 25%–75%, and >75% indicating low, moderate, and high heterogeneity, respectively. Pooled RRs with corresponding 95% CIs were calculated for each phthalate exposure.

Subgroup analyses were performed to determine potential differences in risk estimates based on sex (female vs. male) and exposure timing (prenatal vs. postnatal). This stratification enabled a more detailed evaluation of demographic and temporal factors that might modify the association between phthalate exposure and early puberty. We conducted meta-regression analyses for several phthalate metabolites to examine the effects of study characteristics on the observed effect sizes. Specifically, these analyses were conducted for mono-(2-ethyl-5-hydroxyhexyl) phthalate, mono-(2-ethyl-5-oxohexyl) phthalate, mono-benzyl phthalate (MBzP), MEHP, mono-ethyl phthalate (MEP), and MnBP in the overall group. Additional analyses were conducted for MBzP and MEP in the unadjusted group; this is because the sample size of studies assessing these metabolites was sufficient. In the meta-regression model, we included sex (male or female) and exposure timing (prenatal or postnatal) as predictors.

Meta-regression was performed using a random-effects model. A p value of <0.05 indicated statistical significance.

Publication bias was evaluated using funnel plots for analyses that included 10 or more studies. All analyses were conducted using R software (version 4.4.1).

## Results

### Characteristics of included studies

Details of the individual studies included in the meta-analysis are presented in Table I. The studies were published between 2014 and 2018, and their sample sizes ranged from 103 to 1141 participants. These studies examined the markers of pubertal onset (thelarche and menarche in girls and testicular enlargement in boys) as primary outcomes. Among the included studies, nine evaluated postnatal phthalate exposure, whereas four focused on prenatal exposure.

**Table 1:**
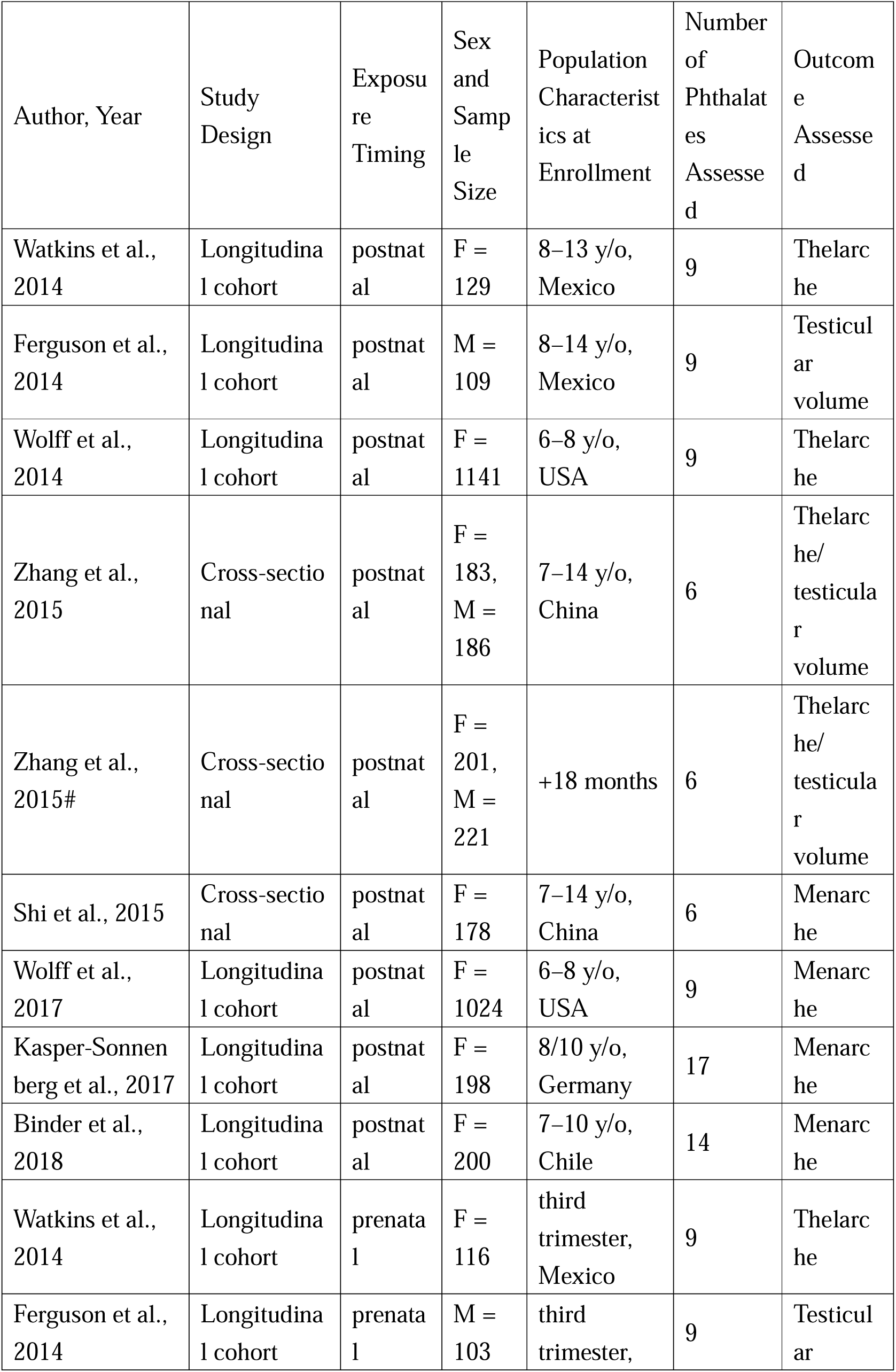
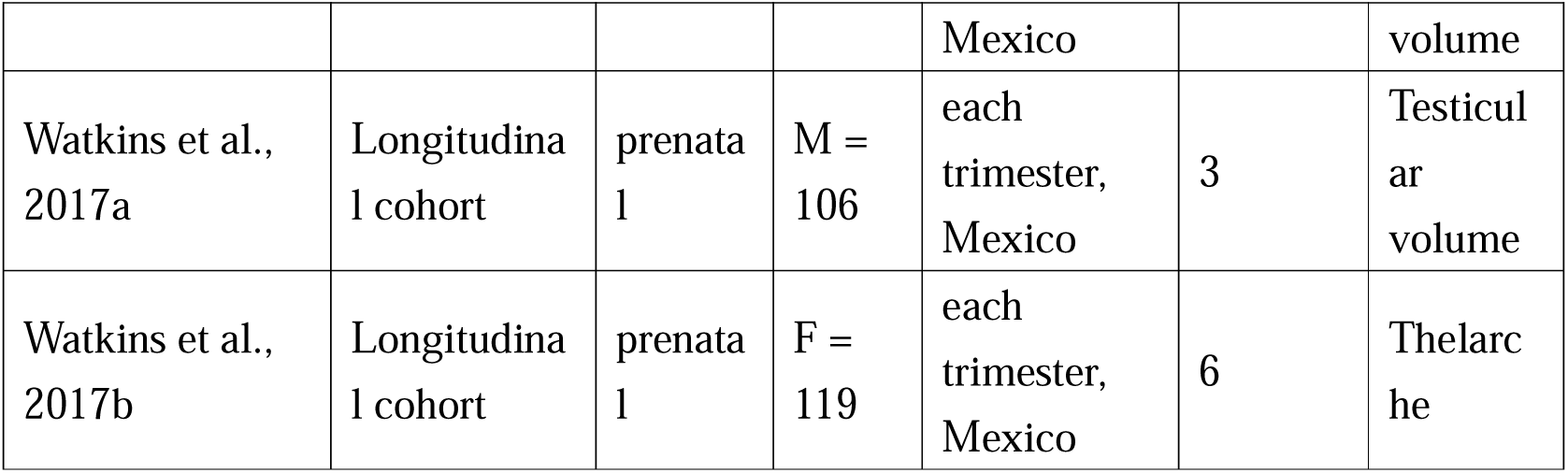
Characteristics of included studies.

| Author, Year | Study Design | Exposure Timing | Sex and Sample Size | Population Characteristics at Enrollment | Number of Phthalates Assessed | Outcome Assessed |
| --- | --- | --- | --- | --- | --- | --- |
| Watkins et al., 2014 | Longitudinal cohort | postnatal | F = 129 | 8–13 y/o, Mexico | 9 | Thelarche |
| Ferguson et al., 2014 | Longitudinal cohort | postnatal | M = 109 | 8–14 y/o, Mexico | 9 | Testicular volume |
| Wolff et al., 2014 | Longitudinal cohort | postnatal | F = 1141 | 6–8 y/o, USA | 9 | Thelarche |
| Zhang et al., 2015 | Cross-sectional | postnatal | F = 183, M = 186 | 7–14 y/o, China | 6 | Thelarche/<br>testicular volume |
| Zhang et al., 2015# | Cross-sectional | postnatal | F = 201, M = 221 | +18 months | 6 | Thelarche/<br>testicular volume |
| Shi et al., 2015 | Cross-sectional | postnatal | F = 178 | 7–14 y/o, China | 6 | Menarche |
| Wolff et al., 2017 | Longitudinal cohort | postnatal | F = 1024 | 6–8 y/o, USA | 9 | Menarche |
| Kasper-Sonnenberg et al., 2017 | Longitudinal cohort | postnatal | F = 198 | 8/10 y/o, Germany | 17 | Menarche |
| Binder et al., 2018 | Longitudinal cohort | postnatal | F = 200 | 7–10 y/o, Chile | 14 | Menarche |
| Watkins et al., 2014 | Longitudinal cohort | prenatal | F = 116 | third trimester, Mexico | 9 | Thelarche |
| Ferguson et al., 2014 | Longitudinal cohort | prenatal | M = 103 | third trimester, | 9 | Testicular |
|  |  |  |  | Mexico |  | volume |
| Watkins et al.,<br>2017a | Longitudinal cohort | prenatal | M =<br>106 | each trimester,<br>Mexico | 3 | Testicular<br>volume |
| Watkins et al.,<br>2017b | Longitudinal cohort | prenatal | F =<br>119 | each trimester,<br>Mexico | 6 | Thelarche |

### Assessment of study quality

The results of the NOS evaluation for cohort studies are summarized in Supplementary Table 1 ^16^. Six of the eight studies were classified as high quality, whereas the remaining two were classified as moderate quality. The results of the adapted NOS evaluation for cross-sectional studies are presented in Supplementary Table 2 ^16^. Both studies were classified as high quality.

### Main findings

This meta-analysis investigated the association between various phthalate metabolites and the risk of early puberty. RRs, 95% CIs, and heterogeneity were calculated using R software (version 4.4.1). The association between multiple phthalate types and the risk of early puberty was analyzed. A summary of the outcomes is provided in Figure 2, and separate forest and funnel plots are presented in Supplementary Figures 1–4 ^16^; additional plots for specific outcomes, namely thelarche, menarche, and testicular volume, are included in Supplementary Figures 5–7 ^16^. The results indicated that most metabolites were not significantly associated with the risk of early puberty in the overall, unadjusted, or urine-SG-adjusted analyses. However, two metabolites, mono-(3-carboxypropyl) phthalate (MCPP) and MnBP, exhibited modest associations in the unadjusted analyses. Specifically, MCPP exposure was associated with a slight reduction in the risk of early puberty (pooled RR: 0.955, 95% CI: 0.917–0.995; Figure 3), whereas postnatal MnBP exposure was associated with an increased risk of early puberty in boys (RR: 1.03, 95% CI: 1.01–1.06; Figure 4). These findings demonstrate the potential sex-specific effects of these metabolites on pubertal timing.

**Figure 2.**
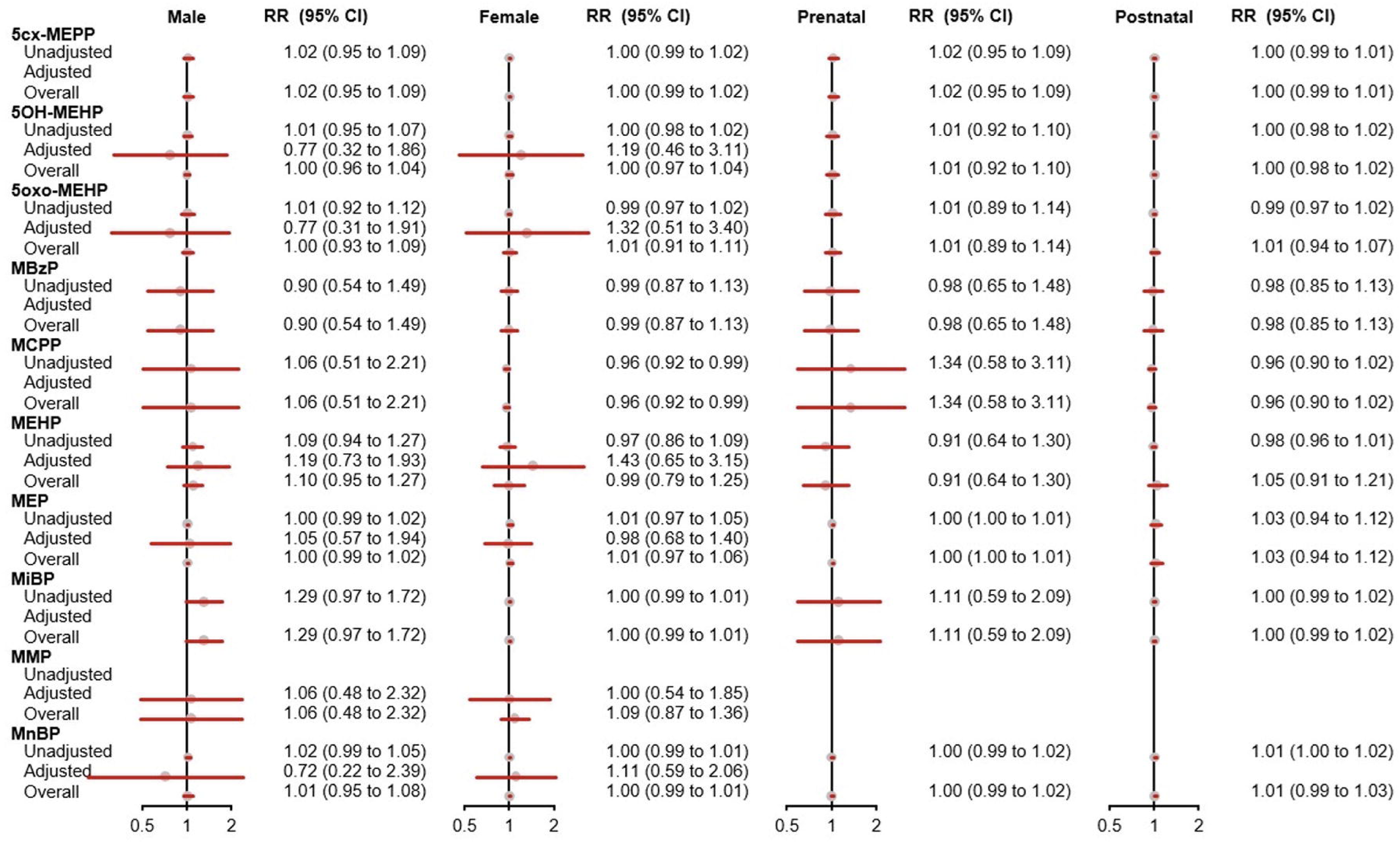
Summary of outcomes

**Figure 3.**
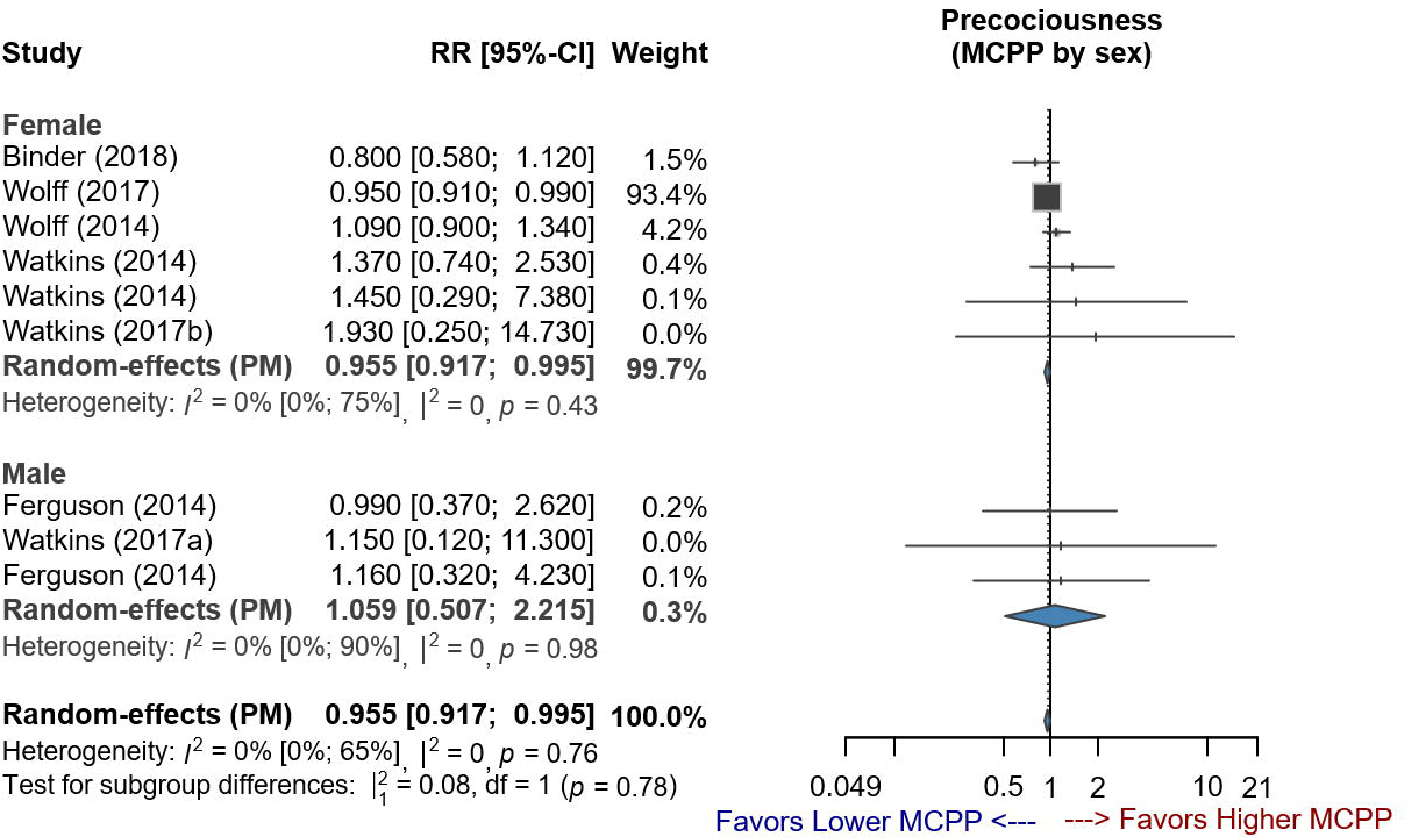
Forest plot of studies demonstrating the RR of MCPP exposure for early puberty stratified by sex

**Figure 4.**
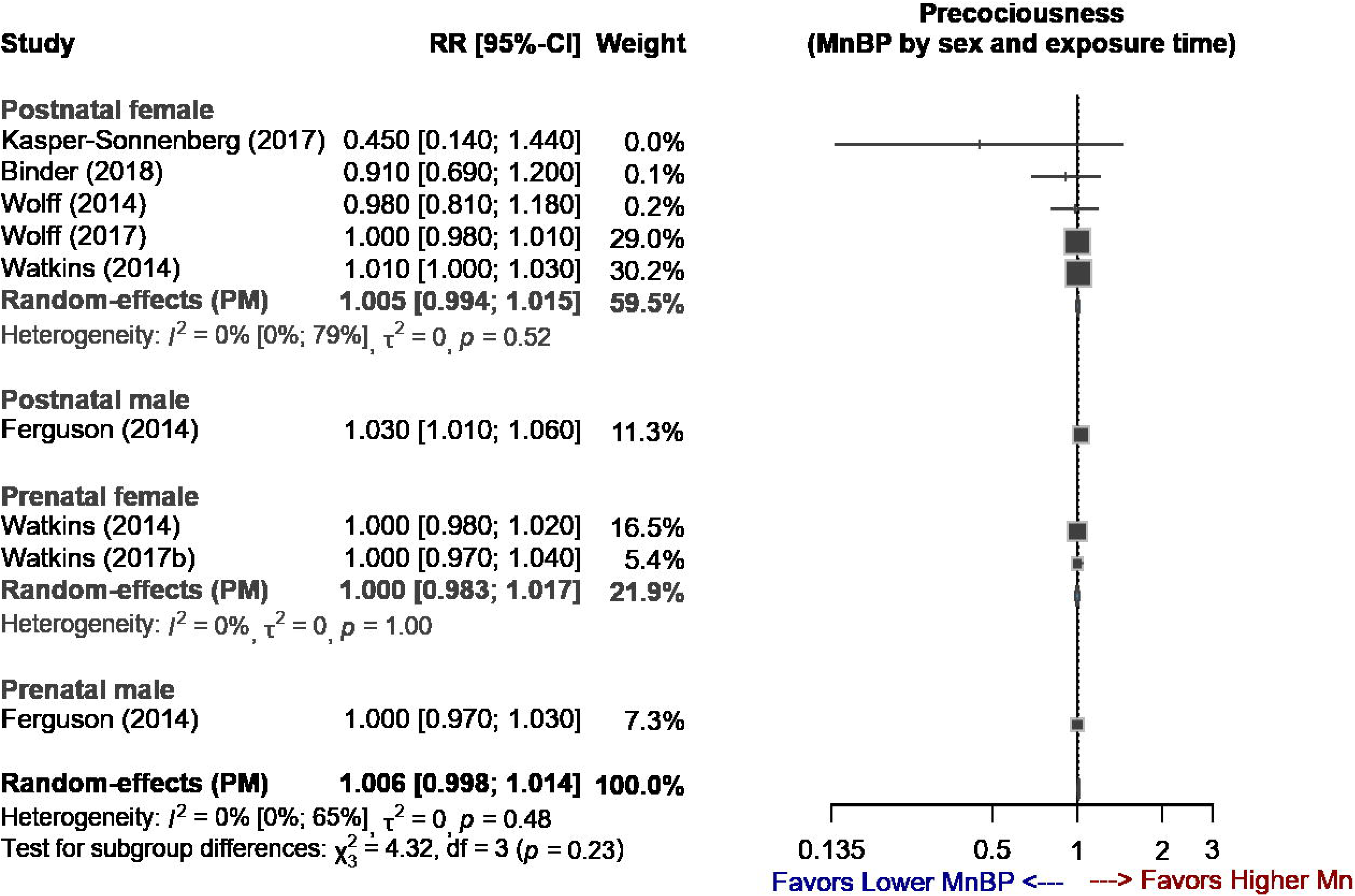
Forest plot of studies demonstrating the RR of MnBP exposure for early puberty stratified by sex and exposure time

The association of MCPP exposure with the risk of early puberty was particularly pronounced in the female subgroup, which accounted for the majority of the weight in the meta-analysis. By contrast, only MnBP exposure was significantly associated with the risk of early puberty in boys. In particular, MnBP exposure resulted in a modest but statistically significant increase in the risk of early puberty. No significant associations between other metabolites and the risk of early puberty were observed in boys. However, the limited number of studies and small sample sizes reduced the statistical power to detect potential effects.

After adjustment for urine SG, which accounts for variability in urine concentration, none of the metabolites, including MCPP and MnBP, were observed to be significantly associated with early puberty(Supplementary Figure 2) ^16^. These findings indicate that measurement variability or residual confounding factors may have affected the modest associations observed in the unadjusted analyses. In all analyses, the strength of evidence for male subgroups and some metabolites was limited owing to small sample sizes and wide CIs.

### Subgroup analyses

Subgroup analyses revealed that when the data were stratified by sex or exposure timing, no significant differences were observed in the effects of all phthalates (Supplementary Figures 1–3) ^16^.

### Meta-regression

Meta-regression analysis results revealed that neither sex nor exposure timing significantly affected the association between phthalate exposure and early puberty (Supplementary Table 3) ^16^.

### Publication bias

To evaluate potential publication bias, funnel plots were generated for mono-(2-ethyl-5-hydroxyhexyl) phthalate, mono-(2-ethyl-5-oxohexyl) phthalate, MBzP, MEHP, MEP, and MnBP in the overall group and for MBzP and MEP in the unadjusted group. These metabolites were selected because they were reported in 10 or more studies. Visual inspection of the funnel plots did not reveal any substantial asymmetry, indicating a low likelihood of publication bias. In addition, Egger’s linear regression test was conducted to examine funnel plot asymmetry, and nonsignificant *p* values were obtained for all phthalate metabolites, confirming the absence of substantial publication bias (Supplementary Figure 4) ^16^.

## Discussion

The findings of this meta-analysis provide insights into the association between phthalate exposure and early puberty. Although the overall results did not demonstrate strong and consistent associations between exposure to all phthalates and early puberty, specific patterns were observed for MnBP and MCPP exposure.

Our analysis revealed a modest association between postnatal MnBP exposure and an increased risk of early puberty in boys; however, this finding was based on data from a single study. By contrast, we observed that exposure to MCPP was associated with a slight reduction in the risk of early puberty among girls. The female subgroup contributed the majority of the weight in this analysis. These associations weakened after adjustment for urine SG, indicating that variability in exposure measurement may have influenced the observed effects. We determined no significant associations between exposure to other phthalates and early pubertal markers across subgroups. Furthermore, our subgroup and meta-regression analyses revealed that sex or exposure timing did not significantly affect the association between phthalate exposure and early puberty, indicating the need for more focused investigations into these potential modifiers.

Our findings differ from those of previous studies. For example, Burns et al. reported no significant association between postnatal MnBP exposure and pubertal timing in boys, and Zhou et al. observed an association between MnBP exposure and decreased pubertal development in boys ^17,18^. This discrepancy in findings may partly be due to our reliance on data from a single study for MnBP in our analysis, which may have limited the robustness of the observed associations. The small evidence base introduces the risk of sampling variability and may not fully capture the range of potential effects observed in larger populations.

The effects of MCPP on pubertal development are challenging to interpret because of the limited number of new studies available for comparison. The observed association between MCPP exposure and a reduced risk of early puberty in girls was unexpected because of the antiandrogenic effects of phthalates, which are generally considered to advance pubertal onset in girls. This discrepancy highlights the scarcity of recent research on MCPP and the potential involvement of unaccounted-for biological mechanisms or measurement artifacts. Targeted investigations into biological and epidemiological pathways affected by MCPP exposure are necessary.

Potential mechanisms underlying the effects of phthalates on pubertal timing involve the disruption of endocrine signaling through the HPG axis. Phthalates affect gonadotropin receptors, including gonadotropin-releasing, follicle-stimulating, and luteinizing hormones, and nuclear receptors, such as androgen, estrogen, and peroxisome proliferator-activated receptors. These disruptions affect hormone regulation and gene expression in reproductive tissues ^19–21^. In addition, phthalates induce epigenetic modifications, including DNA methylation and alterations in the expression of genes in gonadal cells, which may engender transgenerational effects ^22^. The dose of phthalates may also contribute to the variability in results because phthalates exhibit nonmonotonic toxicity; this implies that they can exert different physiological effects at low and high doses ^23^. These mechanisms may collectively explain the observed associations across studies, with nonmonotonic toxicity providing a plausible reason for the variability in results. Other contributing factors, such as ethnic differences in exposure levels and susceptibility, variability in study design, co-exposure to other EDCs, and lifestyle or socioeconomic factors, may also account for the inconsistencies in findings ^24^.

New studies have reported strong associations between phthalate metabolites and pubertal timing, which were not observed in our analysis. For example, Freire et al. reported that prenatal exposure to di-iso-nonyl phthalate metabolites, such as mono-hydroxy-isononyl phthalate and mono-oxo-isononyl phthalate, was associated with accelerated puberty in boys, particularly those with higher body mass index; they also observed an association between prenatal MEHP exposure and delayed gonadarche in girls ^25^. Kim et al. examined the effects of high-and low-molecular-weight phthalates and reported that although both types were inversely correlated with male pubertal development, low-molecular-weight phthalate exposure was associated with advanced pubertal stages in girls ^26^. Similarly, Zheng et al. suggested that the levels of di-(2-ethylhexyl) phthalate metabolites are associated with progression from isolated premature thelarche to central precocious puberty or early puberty ^27^. Liu et al. demonstrated that persistent phthalate exposure substantially affects pubertal onset; synergistic interactions between phthalates and estradiol were determined to accelerate puberty in both sexes, whereas antagonistic interactions between phthalates and testosterone in boys appeared to delay their development ^28^. These findings indicate the variability in the effects of phthalate on pubertal timing.

Overall, our findings align with those of the original meta-analysis, which highlighted variability in the association between phthalate exposure and pubertal timing ^9^. By focusing on individual metabolites and applying meta-regression analyses, we were able to identify nuanced effects that were less apparent in the broader analysis. This reinforces the need for more targeted investigations with standardized methodologies.

Our study has several limitations. First, while we incorporated data from a previously published meta-analysis, the lack of standardized reporting across newer studies posed challenges in directly integrating them. This highlights the ongoing need for harmonization in exposure assessments and outcome reporting. Second, small sample sizes in some subgroups, particularly among boys, reduced the statistical power to detect significant associations. Third, the variability in exposure assessment methods, such as urine-SG adjustments, underscores the challenges of standardizing phthalate measurement across studies. Additionally, although our assessment of publication bias by using funnel plots and Egger’s test did not reveal substantial asymmetry, the small number of studies included in some analyses weakens the robustness of these findings and leaves the possibility of undetected bias. Finally, we cannot rule out residual confounding because the associations observed in unadjusted analyses may reflect unmeasured factors instead of direct causal effects.

Future studies should prioritize the standardization of exposure measurement and outcome reporting to improve comparability across studies. Longitudinal designs with larger and more diverse populations are warranted to identify critical windows of vulnerability and elucidate the mediating role of sex-specific hormonal pathways in the effects of phthalates on puberty. In addition, the interactions between phthalates and other EDCs should be investigated to better understand the cumulative impact of mixed exposures.

## Conclusion

This updated meta-analysis demonstrated the complex association between phthalate exposure and early puberty. Although we did not identify strong and consistent associations for most metabolites, our findings indicate modest sex-specific effects for MnBP and MCPP, which warrant further investigation. These results demonstrate the importance of standardized methodologies and larger, more comprehensive studies to fully elucidate the effects of phthalates on pubertal development. Addressing these gaps in knowledge will enable future research to provide a stronger evidence base for public health policies designed to reduce exposure to EDCs.

## Data Availability

All data produced in the present study are available upon reasonable request to the authors

https://zenodo.org/records/15308932

## Abbreviations

2OH-MiBP: 2OH-mono-isobutyl phthalate
3OH-MnBP: 3OH-mono-n-butyl phthalate
5cx-MEPP: mono(2-ethyl-5-carboxypentyl)phthalate
5OH-MEHP: mono(2-ethyl-5-hydroxyhexyl) phthalate
5oxo-MEHP: mono(2-ethyl-5-oxohexyl)phthalate
cx-MiOP: monocarboxy isooctyl phthalate
MBzP: monobenzyl phthalate
MCPP: mono-(3-carboxypropyl) phthalate
MEHP: mono(2-ethylhexyl) phthalate
MEP: monoethyl phthalate
MiBP: mono-isobutyl phthalate
MMP: monomethyl phthalate
MnBP: mono-n-butyl phthalate
EDCs: endocrine-disrupting chemicals
HPG: hypothalamic–pituitary–gonadal

## Acknowledgements

This manuscript was edited by Wallace Academic Editing.

## Conflict of interest

The authors have no conflicts of interest relevant to this article to disclose. All authors declare that there are no relationships or activities that might bias or be perceived to bias their work.

The data underlying this article are available in the article and in its online supplementary material.

