## Supplementary Files for "Association between phthalate exposure and early puberty: An updated meta-analysis exploring sex and exposure time variations"

**Supplementary File**

### **Supplementary Table 1: Quality assessment for longitudinal cohort studies**

|  | Selection | | | | Comparability | Outcome | | |  |
| --- | --- | --- | --- | --- | --- | --- | --- | --- | --- |
| Study | Representativeness of the exposed cohort | Selection of the nonexposed cohort | Ascertainment of exposure | Demonstration that the outcome of interest was not present at start of the study | Comparability of cohorts on the basis of the design or analysis | Assessment of the outcome | Was the follow-up period sufficient for outcomes to occur? | Adequacy of the follow-up of cohorts | Total quality scores |
| Watkins et al., 2014 | * | - | * | * | ** | - | * | * | 7/9 |
| Ferguson et al., 2014 | * | - | * | * | ** | - | * | * | 7/9 |
| Wolff et al., 2014 | * | - | * | * | * | - | * | * | 6/9 |
| Wolff et al., 2017 | * | - | * | * | * | - | * | * | 6/9 |
| Kasper-Sonnenberg et al., 2017 | * | - | * | * | ** | - | * | * | 7/9 |
| Binder et al., 2018 | * | - | * | * | ** | - | * | * | 7/9 |
| Watkins et al., 2017a | * | - | * | * | ** | - | * | * | 7/9 |
| Watkins et al., 2017b | * | - | * | * | ** | - | * | * | 7/9 |

### **Supplementary Table 2: Quality assessment for cross-sectional studies**

|  | Selection | | | | Comparability | Outcome | |  |
| --- | --- | --- | --- | --- | --- | --- | --- | --- |
| Study | Representativeness of cases | Sample size | Nonresponse rate | Ascertainment of the screening/surveillance tool | Potential confounders were investigated by performing subgroup or multivariable analyses | Assessment of the outcome | Statistical test | Total quality scores |
| Zhang et al., 2015 | * | * | * | * | * | * | * | 7/9 |
| Shi et al., 2015 | * | * | * | * | * | * | * | 7/9 |

### **Supplementary Table 3: Results of meta-regression**

Supplementary Table 3-1: Meta-regression for 5OH-MEHP (combined).

| **Term** | **Estimate** | **se** | **uci** | **lci** | **p** | **AIC** | **BIC** |
| --- | --- | --- | --- | --- | --- | --- | --- |
| Intercept | 0.0061 | 0.0740 | 0.1512 | −0.1389 | 0.9340 | 7.7432 | 9.4381 |
| Sex | −0.0067 | 0.1388 | 0.2653 | −0.2787 | 0.9610 |  |  |
| Intercept | 0.0036 | 0.0698 | 0.1405 | −0.1332 | 0.9580 | 7.7452 | 9.4401 |
| Exposure_Time | 0.0030 | 0.1578 | 0.3122 | −0.3062 | 0.9850 |  |  |
| Intercept | 0.0340 | 0.2121 | 0.4496 | −0.3816 | 0.8730 | 6.6433 | 7.5511 |
| Baseline 5OH-MEHP (linear overall) | −0.0007 | 0.0052 | 0.0095 | −0.0110 | 0.8870 |  |  |

Abbreviations: se, standard error; uci, upper confidence interval; lci, lower confidence interval; AIC, Akaike Information Criterion; BIC, Bayesian Information Criterion; 5OH-MEHP, mono(2-ethyl-5-hydroxyhexyl) phthalate

Supplementary Table 3-2: Meta-regression for 5oxo-MEHP (combined).

| **Term** | **Estimate** | **se** | **uci** | **lci** | **p** | **AIC** | **BIC** |
| --- | --- | --- | --- | --- | --- | --- | --- |
| Intercept | 0.0040 | 0.0826 | 0.1659 | −0.1579 | 0.9610 | 10.8638 | 12.5587 |
| Sex | −0.0072 | 0.1836 | 0.3526 | −0.3670 | 0.9690 |  |  |
| Intercept | 0.0010 | 0.0811 | 0.1599 | −0.1578 | 0.9900 | 10.8633 | 12.5582 |
| Exposure_Time | 0.0088 | 0.1957 | 0.3923 | −0.3747 | 0.9640 |  |  |
| Intercept | 0.0927 | 0.2663 | 0.6146 | −0.4291 | 0.7280 | 9.3994 | 10.3072 |
| Baseline 5oxo-MEHP (linear overall) | −0.0043 | 0.0126 | 0.0205 | −0.0290 | 0.7360 |  |  |

Abbreviations: se, standard error; uci, upper confidence interval; lci, lower confidence interval; AIC, Akaike Information Criterion; BIC, Bayesian Information Criterion; 5oxo-MEHP, mono(2-ethyl-5-oxohexyl)phthalate

Supplementary Table 3-3: Meta-regression for MBzP (combined).

| **Term** | **Estimate** | **se** | **uci** | **lci** | **p** | **AIC** | **BIC** |
| --- | --- | --- | --- | --- | --- | --- | --- |
| Intercept | −0.0115 | 0.1077 | 0.1995 | −0.2225 | 0.9150 | 9.6073 | 10.5150 |
| Sex | −0.0755 | 0.2805 | 0.4742 | −0.6252 | 0.7880 |  |  |
| Intercept | −0.0231 | 0.1095 | 0.1915 | −0.2377 | 0.8330 | 9.6797 | 10.5874 |
| Exposure_Time | 0.0027 | 0.2615 | 0.5152 | −0.5097 | 0.9920 |  |  |

Abbreviations: se, standard error; uci, upper confidence interval; lci, lower confidence interval; AIC, Akaike Information Criterion; BIC, Bayesian Information Criterion; MBzP, monobenzyl phthalate

Supplementary Table 3-4: Meta-regression for MEHP (combined).

| **Term** | **Estimate** | **se** | **uci** | **lci** | **p** | **AIC** | **BIC** |
| --- | --- | --- | --- | --- | --- | --- | --- |
| Intercept | −0.0124 | 0.0897 | 0.1634 | −0.1883 | 0.8900 | 13.5428 | 15.4600 |
| Sex | 0.1126 | 0.2247 | 0.5531 | −0.3278 | 0.6160 |  |  |
| Intercept | 0.0154 | 0.0887 | 0.1894 | −0.1585 | 0.8620 | 13.7053 | 15.6225 |
| Exposure_Time | −0.0705 | 0.2367 | 0.3934 | −0.5344 | 0.7660 |  |  |
| Intercept | 0.1822 | 0.2825 | 0.7358 | −0.3715 | 0.5190 | 12.0282 | 13.2219 |
| Baseline MEHP  (linear overall) | −0.0433 | 0.0694 | 0.0926 | −0.1793 | 0.5320 |  |  |

Abbreviations: se, standard error; uci, upper confidence interval; lci, lower confidence interval; AIC, Akaike Information Criterion; BIC, Bayesian Information Criterion; MEHP, mono(2-ethylhexyl) phthalate

Supplementary Table 3-5: Meta-regression for MEP (combined).

| **Term** | **Estimate** | **se** | **uci** | **lci** | **p** | **AIC** | **BIC** |
| --- | --- | --- | --- | --- | --- | --- | --- |
| Intercept | 0.0048 | 0.0399 | 0.0831 | −0.0735 | 0.9040 | −4.0925 | −1.9683 |
| Sex | 0.0022 | 0.0761 | 0.1513 | −0.1468 | 0.9770 |  |  |
| Intercept | 0.0076 | 0.0496 | 0.1049 | −0.0897 | 0.8780 | −4.0953 | −1.9711 |
| Exposure_Time | −0.0041 | 0.0681 | 0.1294 | −0.1376 | 0.9520 |  |  |
| Intercept | −0.0095 | 0.1210 | 0.2278 | −0.2467 | 0.9380 | −3.4649 | −1.7701 |
| Baseline MEP (linear overall) | 0.0001 | 0.0012 | 0.0024 | −0.0022 | 0.9170 |  |  |

Abbreviations: se, standard error; uci, upper confidence interval; lci, lower confidence interval; AIC, Akaike Information Criterion; BIC, Bayesian Information Criterion; MEP, monoethyl phthalate

Supplementary Table 3-6: Meta-regression for MnBP (combined).

| **Term** | **Estimate** | **se** | **uci** | **lci** | **P** | **AIC** | **BIC** |
| --- | --- | --- | --- | --- | --- | --- | --- |
| Intercept | −0.0001 | 0.0475 | 0.0930 | −0.0931 | 0.9990 | 0.6739 | 2.5911 |
| Sex | 0.0085 | 0.0945 | 0.1936 | −0.1767 | 0.9280 |  |  |
| Intercept | 0.0033 | 0.0517 | 0.1046 | −0.0980 | 0.9490 | 0.6805 | 2.5977 |
| Exposure_Time | −0.0033 | 0.0850 | 0.1633 | −0.1699 | 0.9690 |  |  |
| Intercept | −0.0148 | 0.1104 | 0.2016 | −0.2312 | 0.8930 | −1.2096 | −0.0159 |
| Baseline MnBP (linear overall) | 0.0003 | 0.0014 | 0.0031 | −0.0025 | 0.8390 |  |  |

Abbreviations: se, standard error; uci, upper confidence interval; lci, lower confidence interval; AIC, Akaike Information Criterion; BIC, Bayesian Information Criterion; MnBP, mono-n-butyl phthalate

Supplementary Table 3-7: Meta-regression for MBzP (unadjusted).

| **Term** | **Estimate** | **se** | **uci** | **lci** | **p** | **AIC** | **BIC** |
| --- | --- | --- | --- | --- | --- | --- | --- |
| Intercept | −0.0115 | 0.1077 | 0.1995 | −0.2225 | 0.9150 | 9.6073 | 10.515 |
| Sex | −0.0755 | 0.2805 | 0.4742 | −0.6252 | 0.7880 |  |  |
| Intercept | −0.0231 | 0.1095 | 0.1915 | −0.2377 | 0.8330 | 9.6797 | 10.5874 |
| Exposure_Time | 0.0027 | 0.2615 | 0.5152 | −0.5097 | 0.9920 |  |  |

Abbreviations: se, standard error; uci, upper confidence interval; lci, lower confidence interval; AIC, Akaike Information Criterion; BIC, Bayesian Information Criterion; MBzP, monobenzyl phthalate

Supplementary Table 3-8: Meta-regression for MEP (unadjusted).

| **Term** | **Estimate** | **se** | **uci** | **lci** | **p** | **AIC** | **BIC** |
| --- | --- | --- | --- | --- | --- | --- | --- |
| Intercept | 0.0054 | 0.0403 | 0.0845 | −0.0736 | 0.8930 | −8.0281 | −7.1203 |
| Sex | 0.0001 | 0.0771 | 0.1513 | −0.1511 | 0.9990 |  |  |
| Intercept | 0.0078 | 0.0509 | 0.1076 | −0.0919 | 0.8780 | −8.032 | −7.1242 |
| Exposure_Time | −0.0043 | 0.0690 | 0.1310 | −0.1396 | 0.9500 |  |  |

Abbreviations: se, standard error; uci, upper confidence interval; lci, lower confidence interval; AIC, Akaike Information Criterion; BIC, Bayesian Information Criterion; MEP, monoethyl phthalate

**Supplementary Figure 1 (1-1~1-33) : Forest plots (combined-reported phthalate level for urine-SG)**

Abbreviations:

urine-SG, urine specific gravity; 2OH-MiBP, 2OH-mono-isobutyl phthalate; 3OH-MnBP, 3OH-mono-n-butyl phthalate; 5cx-MEPP, mono(2-ethyl-5-carboxypentyl)phthalate; 5OH-MEHP, mono(2-ethyl-5-hydroxyhexyl) phthalate; 5oxo-MEHP, mono(2-ethyl-5-oxohexyl)phthalate; cx-MiOP, monocarboxy isooctyl phthalate; MBzP, monobenzyl phthalate; MCPP, mono-(3-carboxypropyl) phthalate; MEHP, mono(2-ethylhexyl) phthalate; MEP, monoethyl phthalate; MiBP, mono-isobutyl phthalate; MMP, monomethyl phthalate; MnBP, mono-n-butyl phthalate

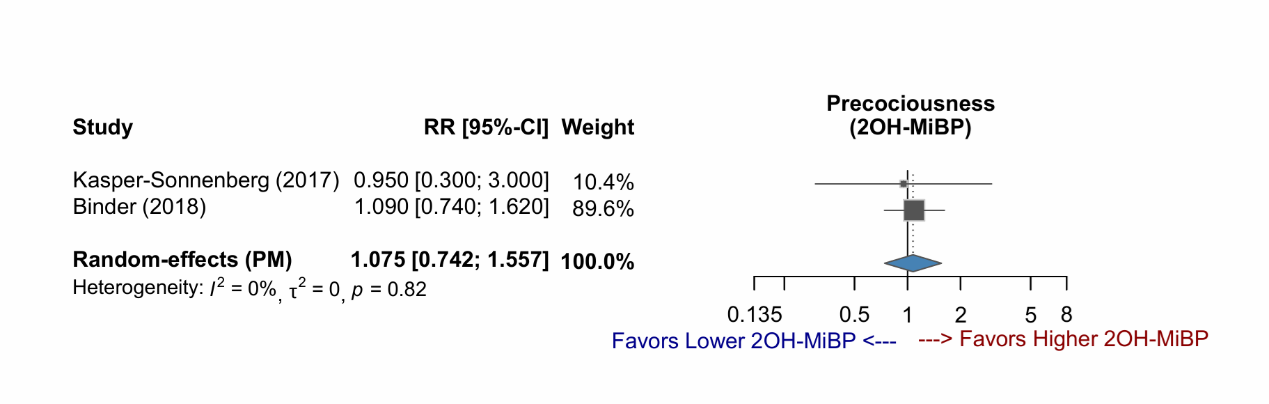

Abbrivia

Supplementary Figure 1-2: Forest plot of studies demonstrating the RR of 3OH-MnBP exposure for early puberty.

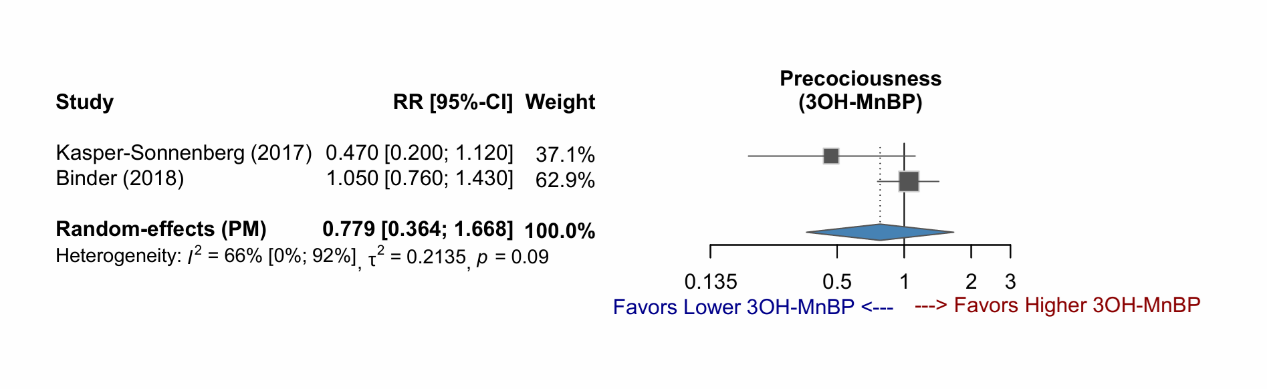

Supplementary Figure 1-3: Forest plot of studies demonstrating the RR of 5cx-MEPP exposure for early puberty, stratified by sex.

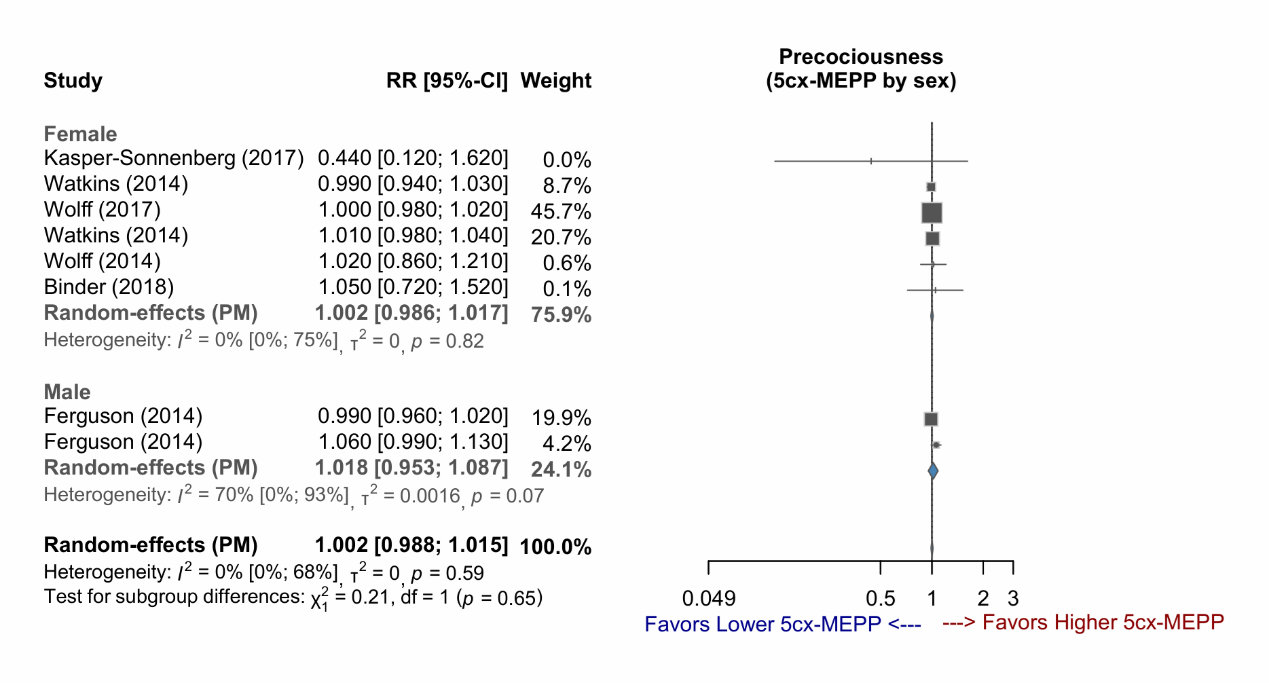

Supplementary Figure 1-4: Forest plot of studies demonstrating the RR of 5cx-MEPP exposure for early puberty, stratified by exposure time.

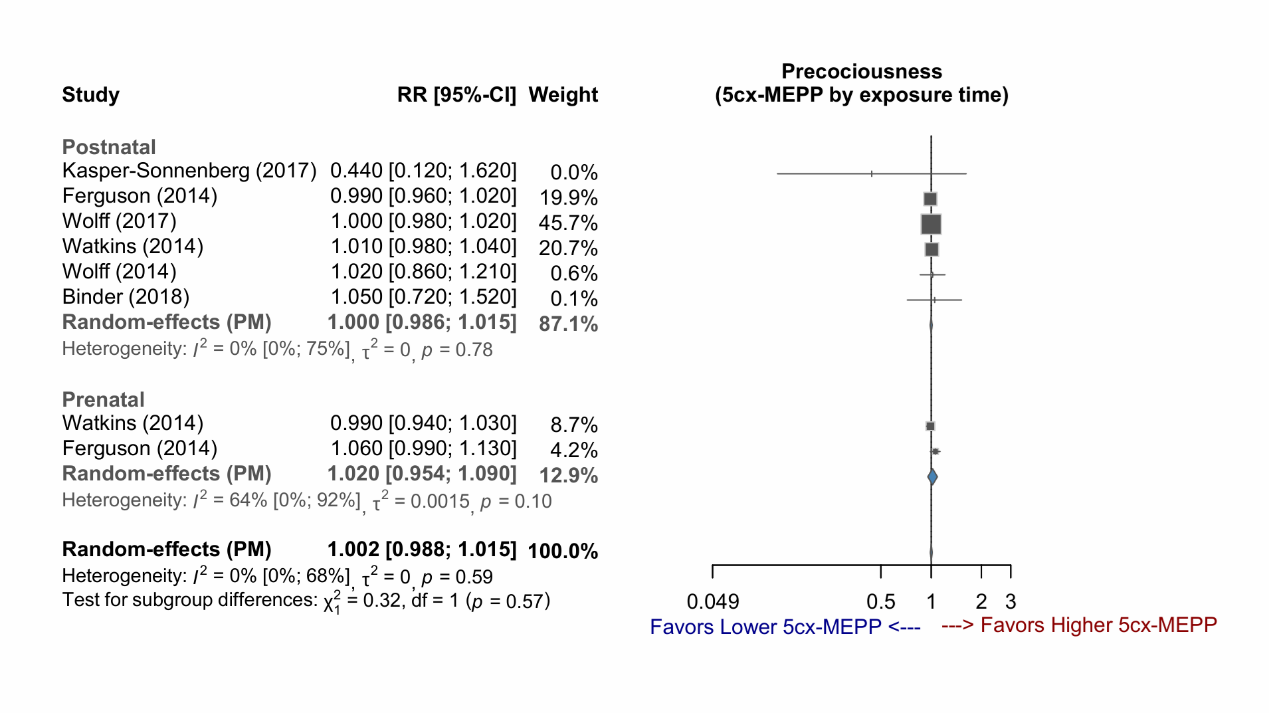

Supplementary Figure 1-5: Forest plot of studies demonstrating the RR of 5cx-MEPP exposure for early puberty, stratified by sex and exposure time.

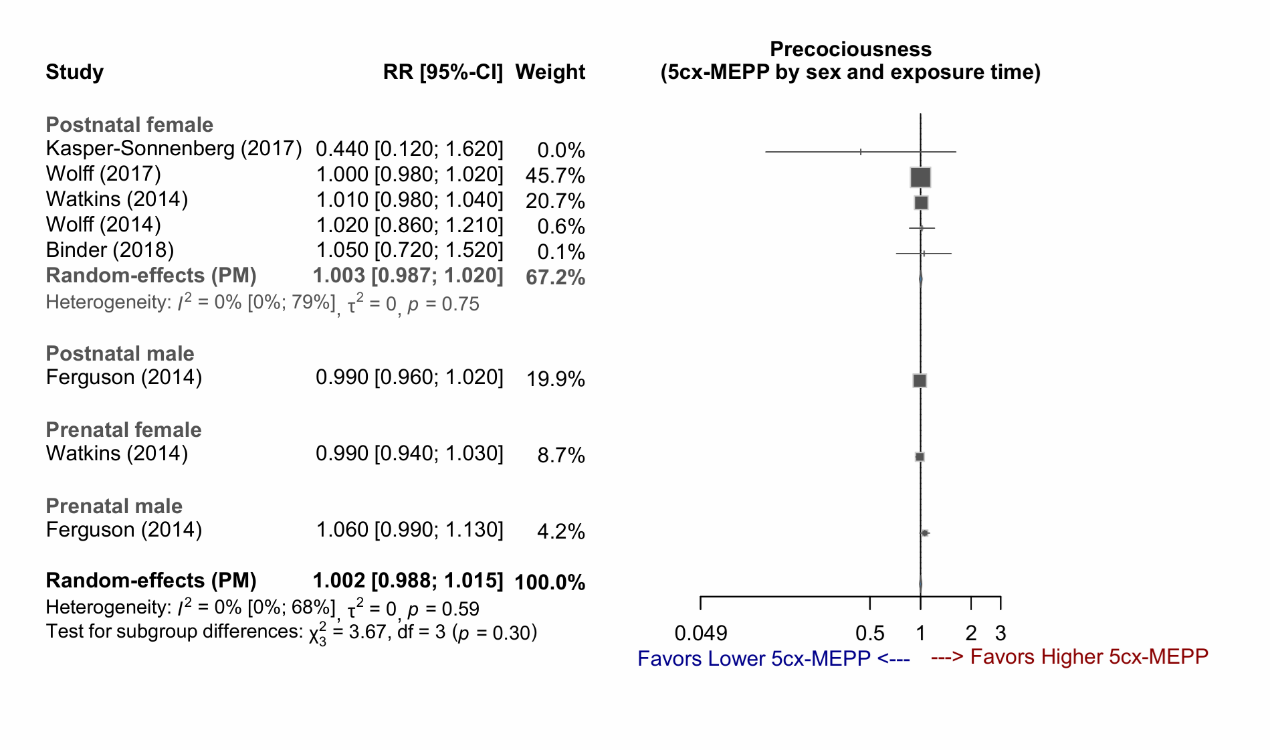

Supplementary Figure 1-6: Forest plot of studies demonstrating the RR of 5OH-MEHP exposure for early puberty, stratified by sex.

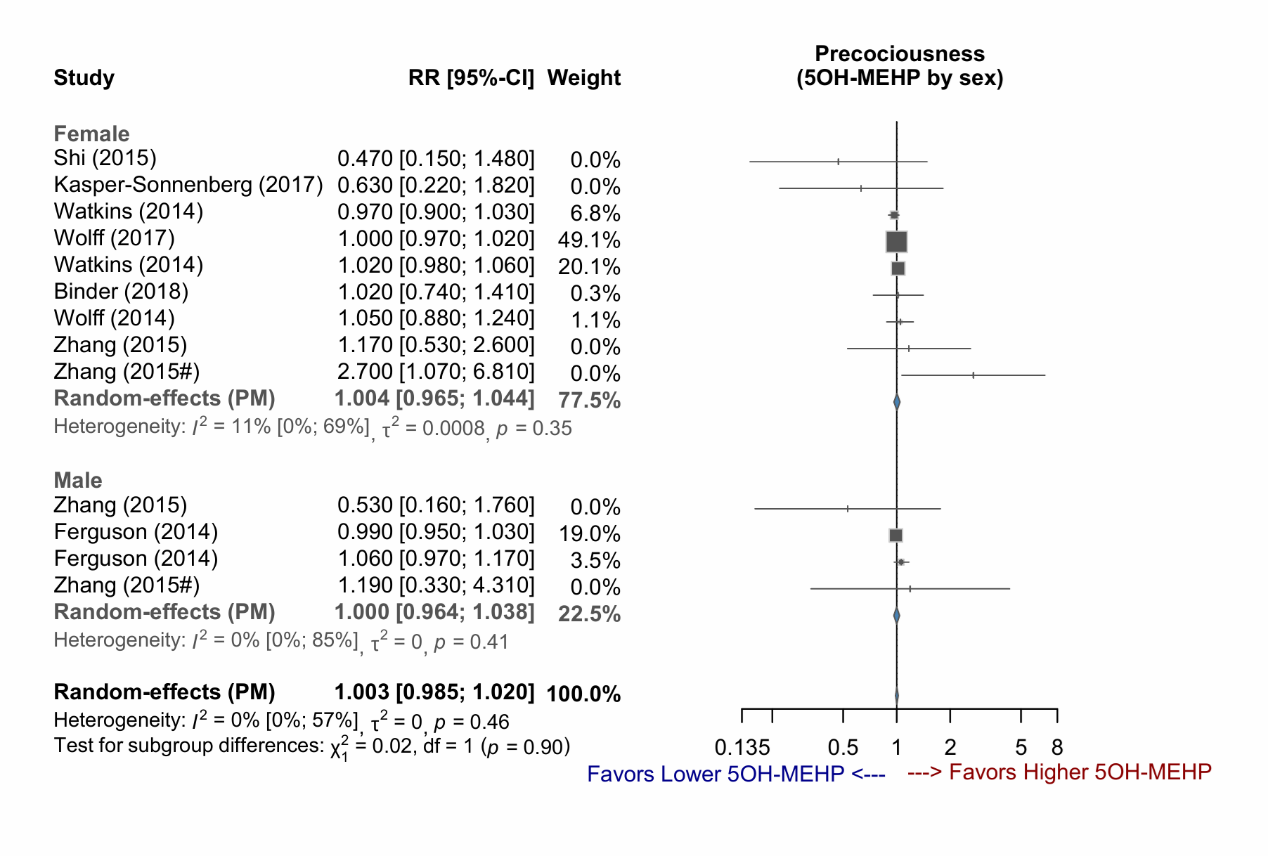

Supplementary Figure 1-7: Forest plot of studies demonstrating the RR of 5OH-MEHP exposure for early puberty, stratified by exposure time.

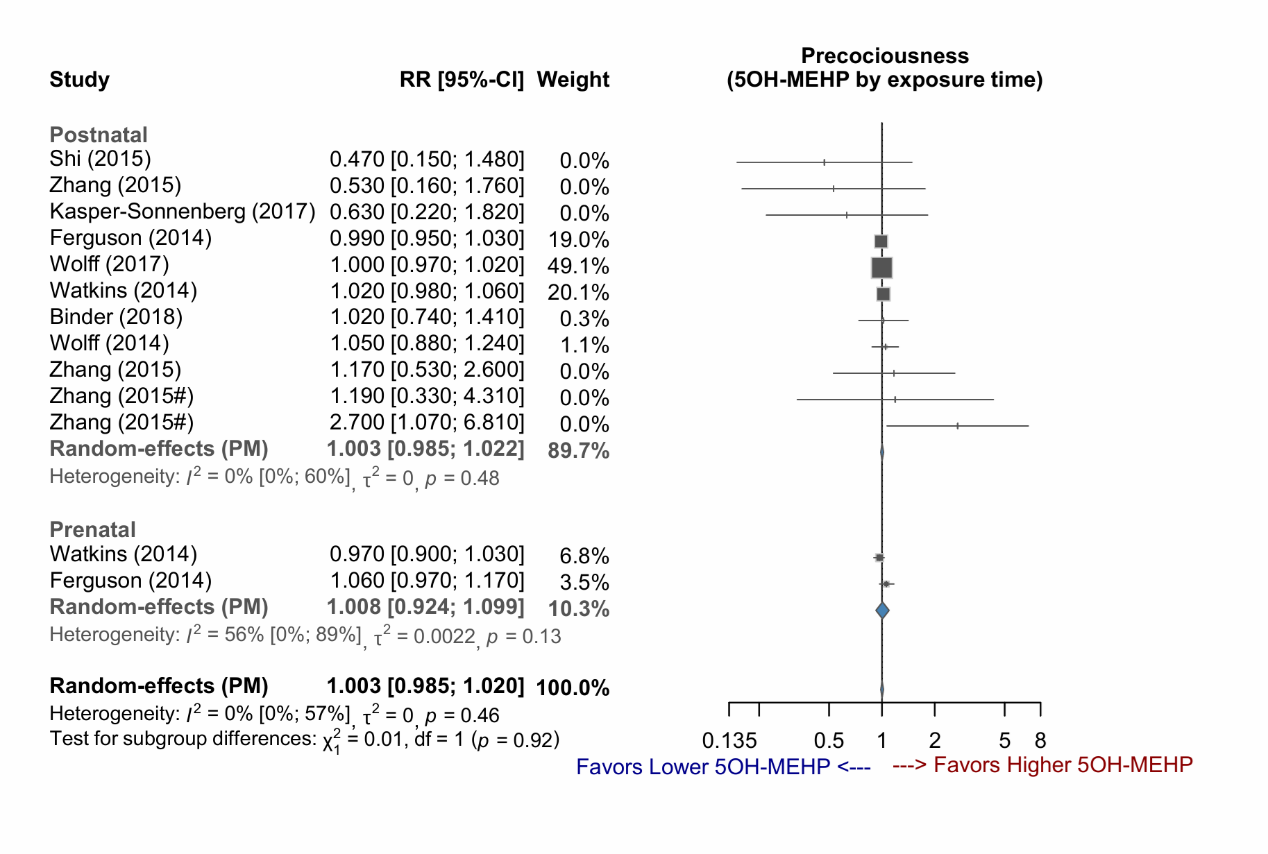

Supplementary Figure 1-8: Forest plot of studies demonstrating the RR of 5OH-MEHP exposure for early puberty, stratified by sex and exposure time.

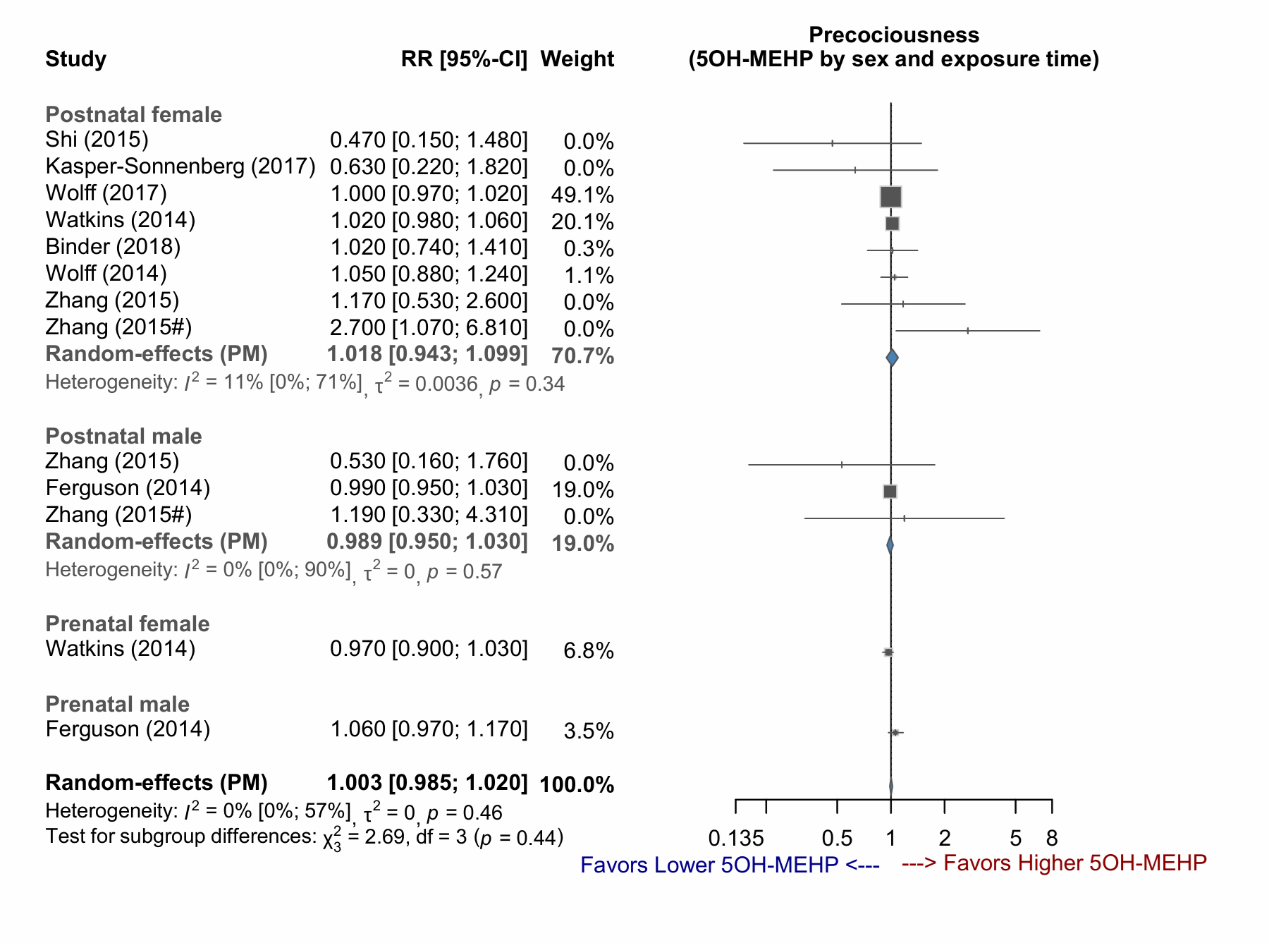

Supplementary Figure 1-9: Forest plot of studies demonstrating the RR of 5oxo-MEHP exposure for early puberty, stratified by sex.

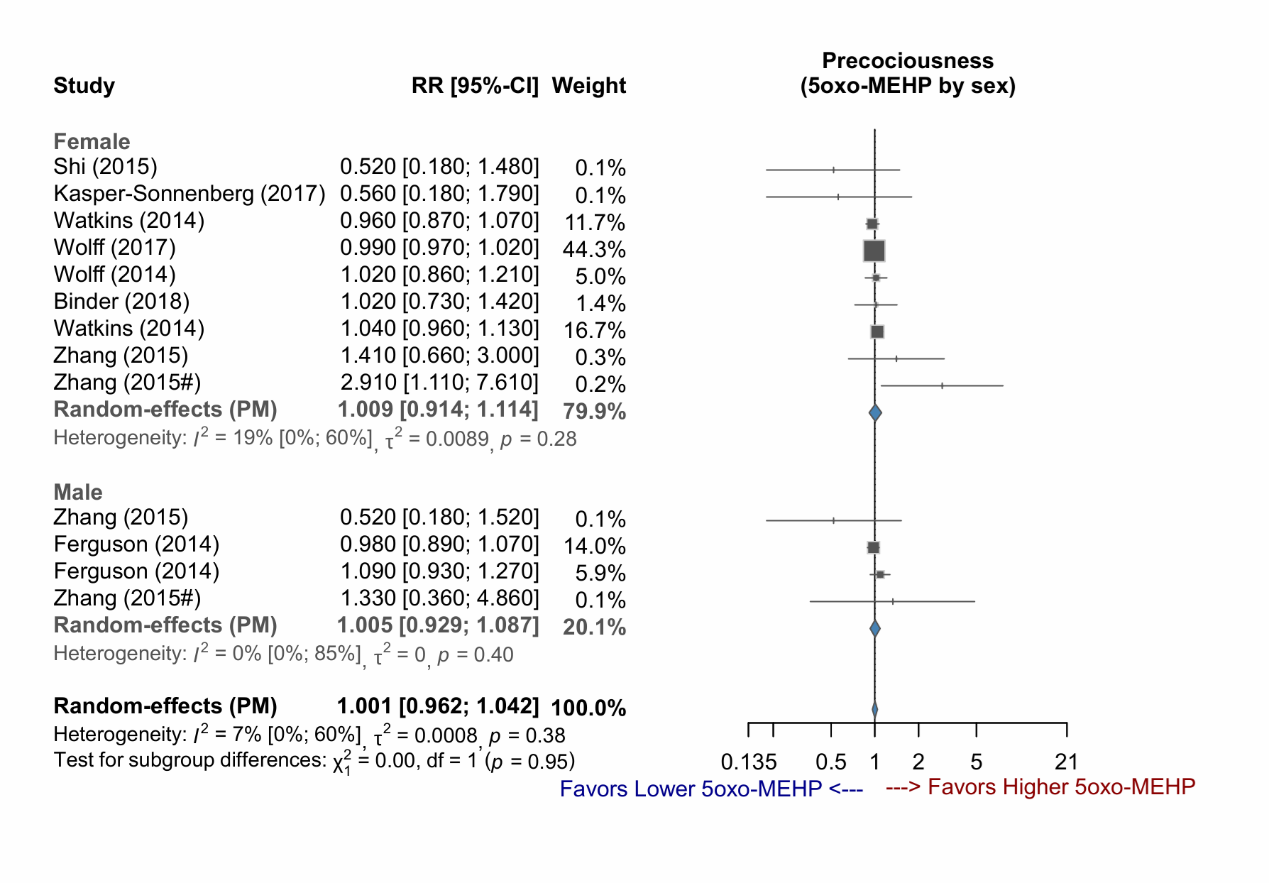

Supplementary Figure 1-10: Forest plot of studies demonstrating the RR of 5oxo-MEHP exposure for early puberty, stratified by exposure time.

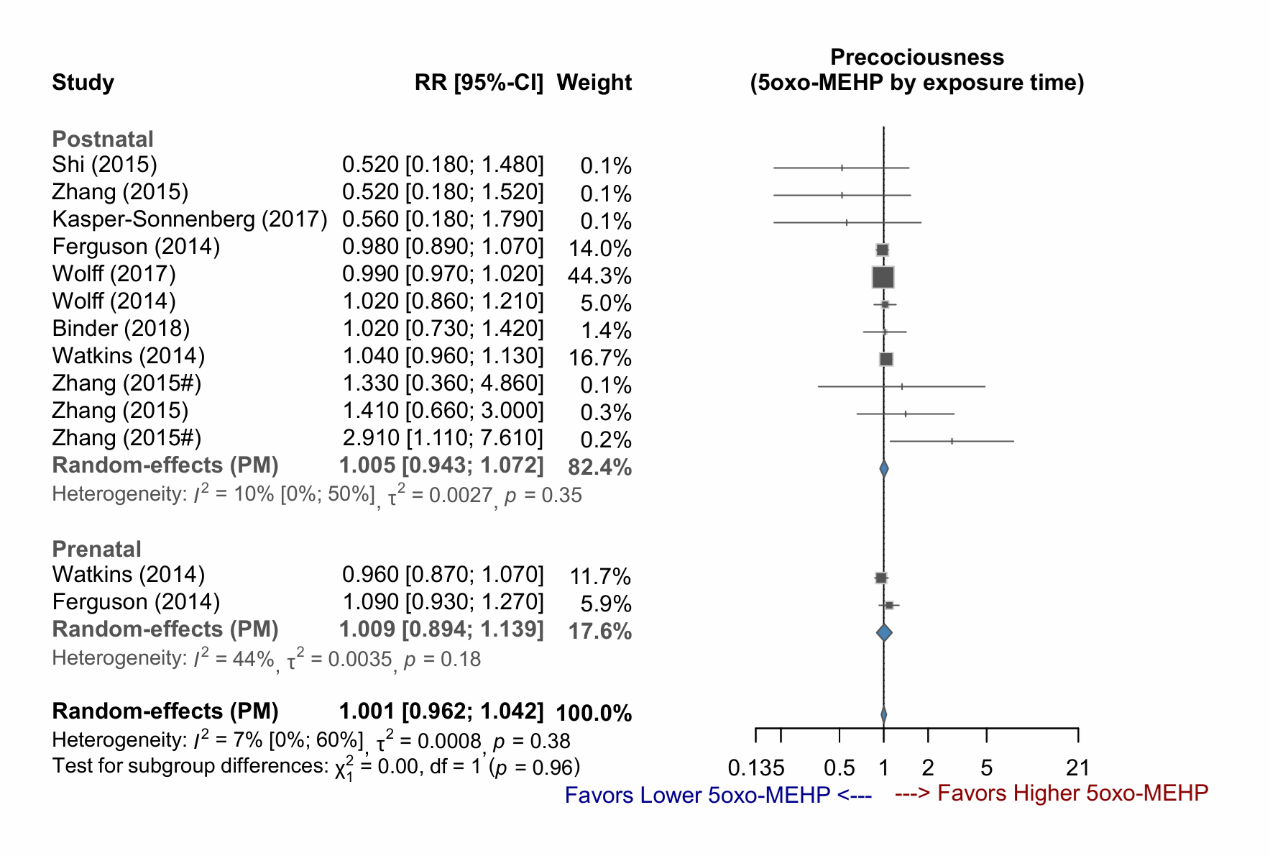

Supplementary Figure 1-11: Forest plot of studies demonstrating the RR of 5oxo-MEHP exposure for early puberty, stratified by sex and exposure time.

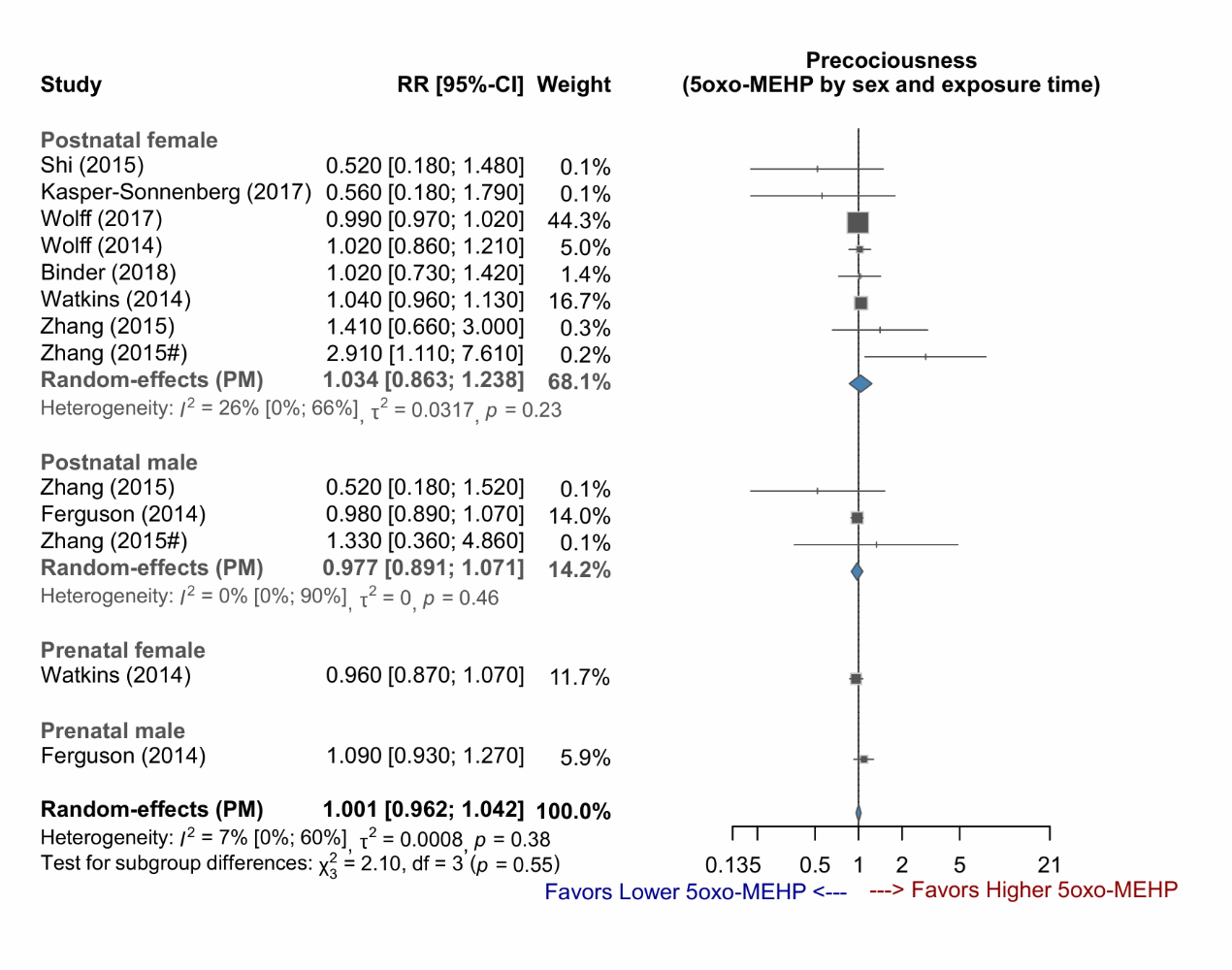

Supplementary Figure 1-12: Forest plot of studies demonstrating the RR of cx-MiOP exposure for early puberty.

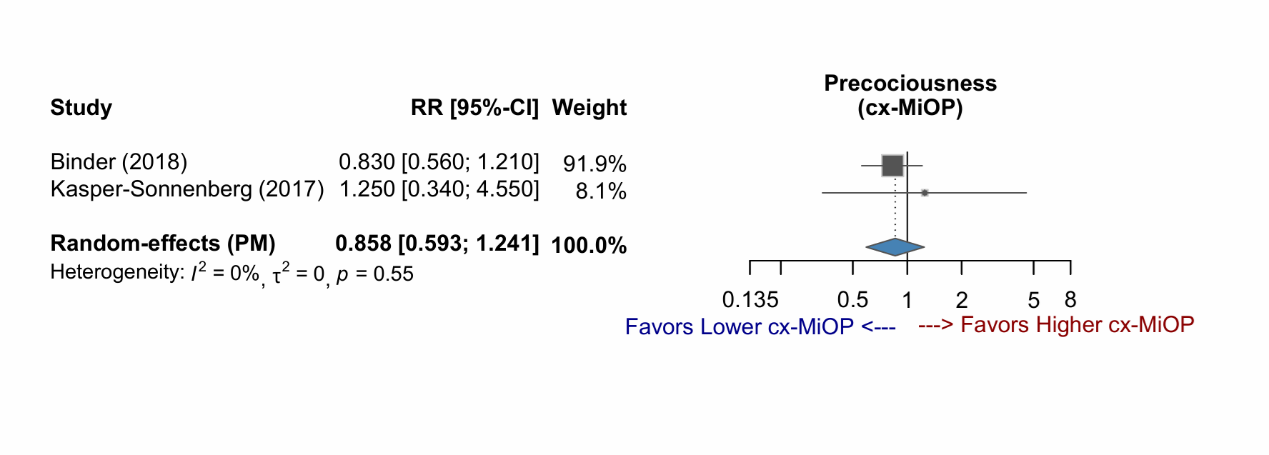

Supplementary Figure 1-13: Forest plot of studies demonstrating the RR of MBzP exposure for early puberty, stratified by sex.

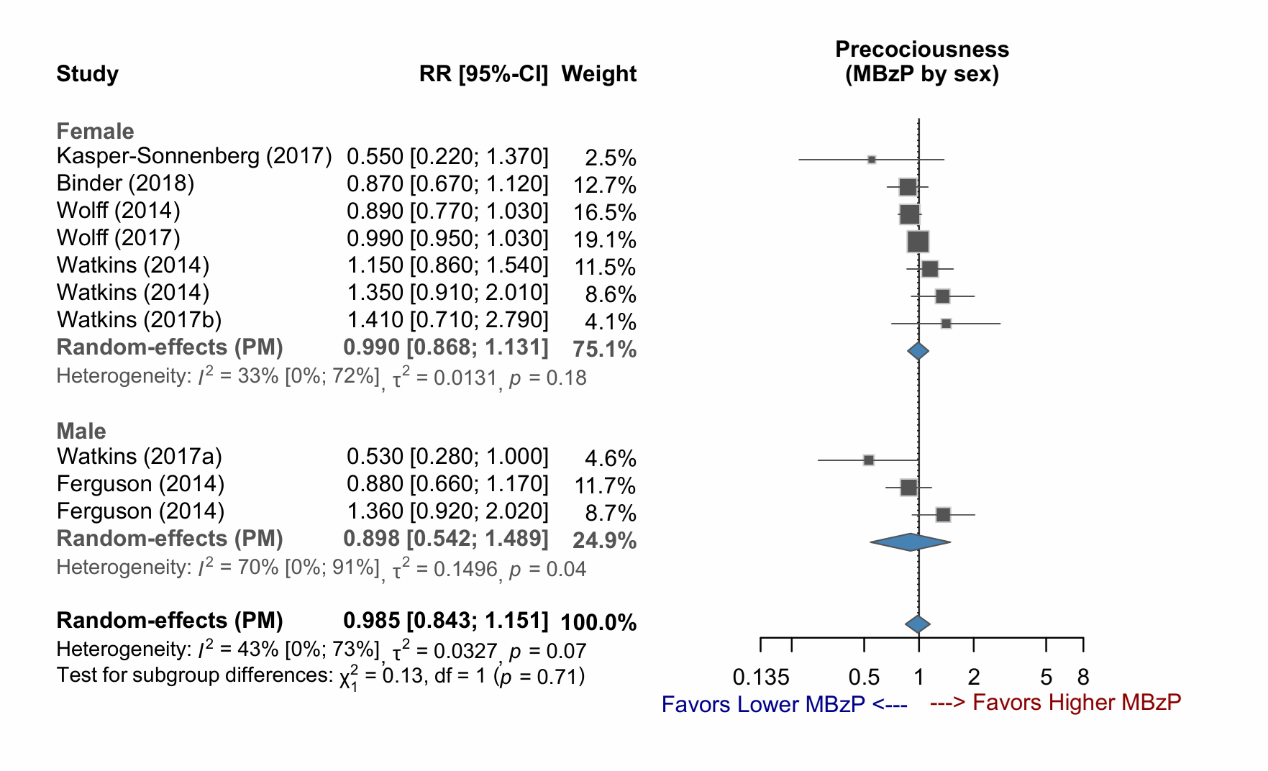

Supplementary Figure 1-14: Forest plot of studies demonstrating the RR of MBzP exposure for early puberty, stratified by exposure time.

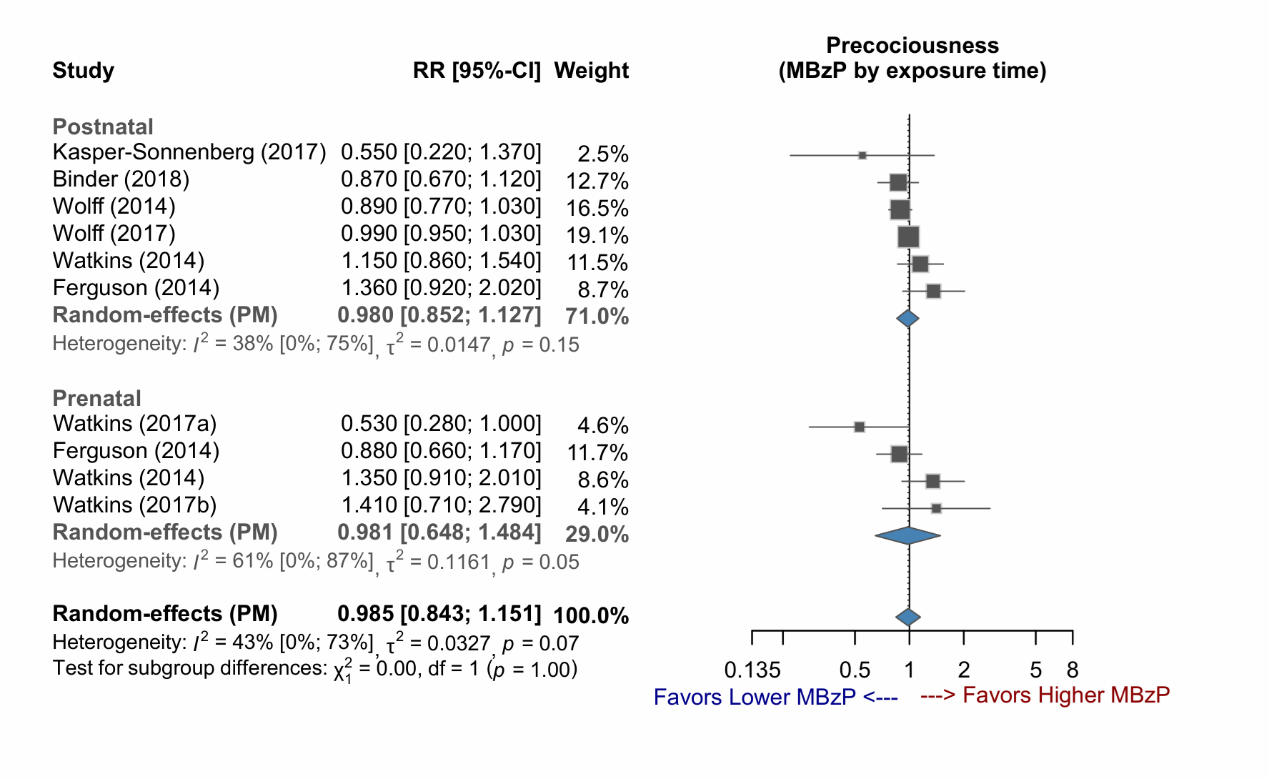

Supplementary Figure 1-15: Forest plot of studies demonstrating the RR of MBzP exposure for early puberty, stratified by sex and exposure time.

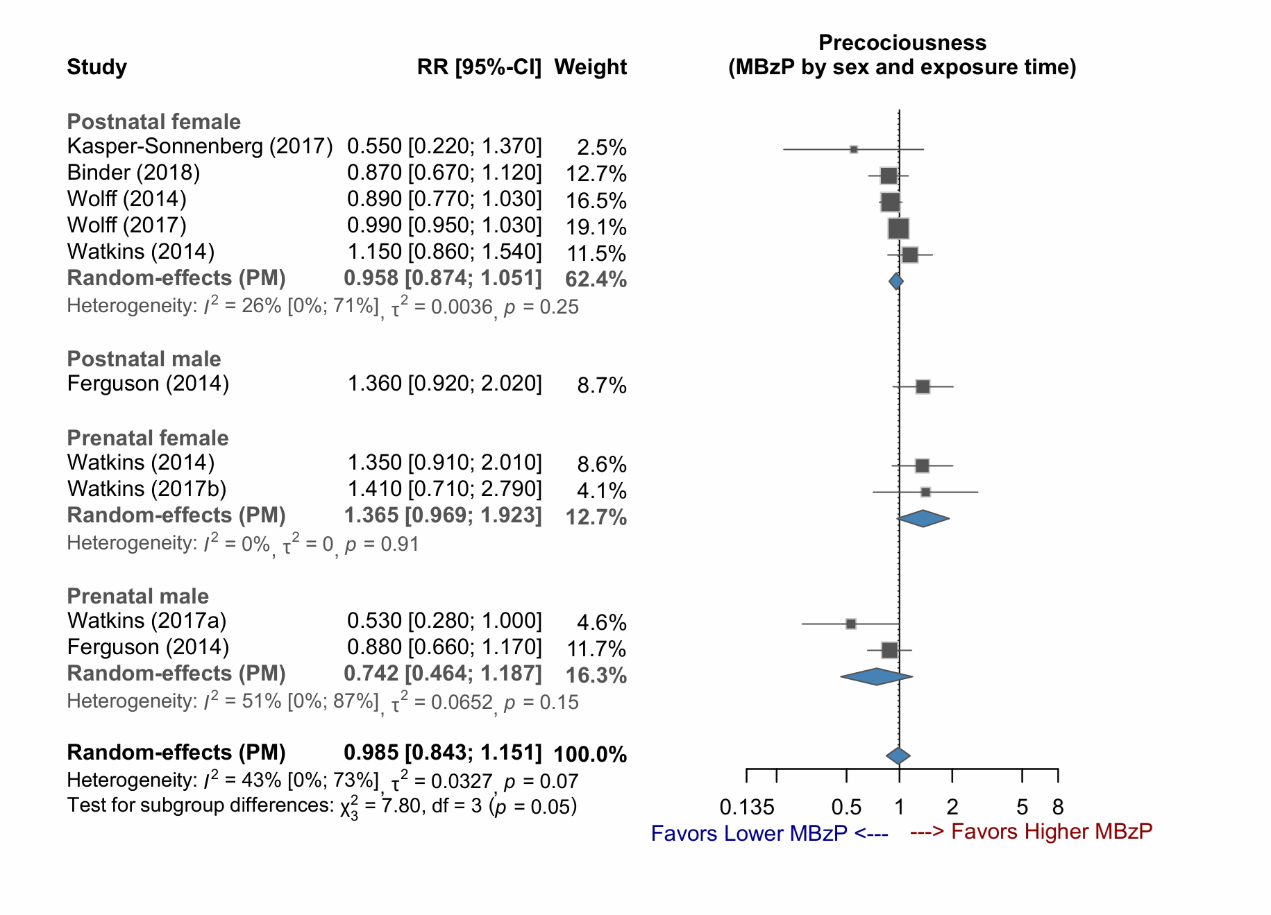

Supplementary Figure 1-16: Forest plot of studies demonstrating the RR of MCPP exposure for early puberty, stratified by sex.

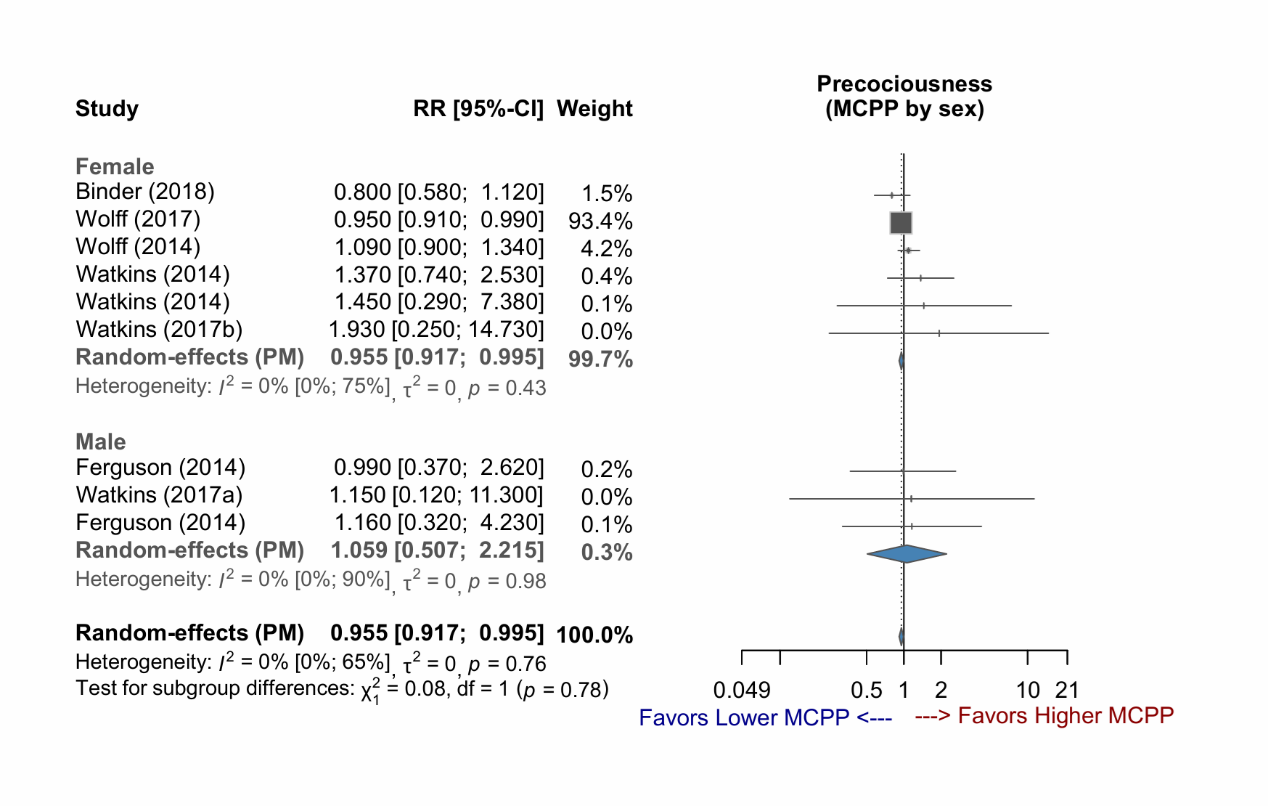

Supplementary Figure 1-17: Forest plot of studies demonstrating the RR of MCPP exposure for early puberty, stratified by exposure time.

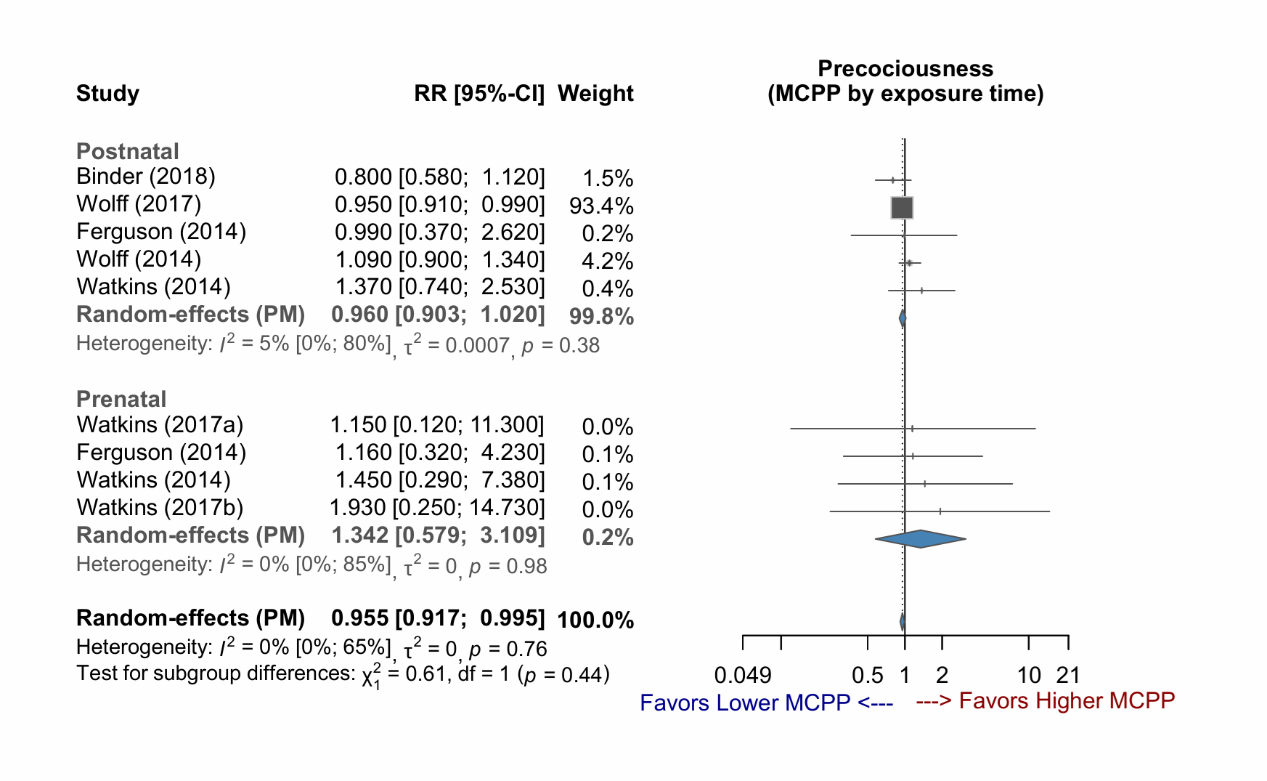

Supplementary Figure 1-18: Forest plot of studies demonstrating the RR of MCPP exposure for early puberty, stratified by sex and exposure time.

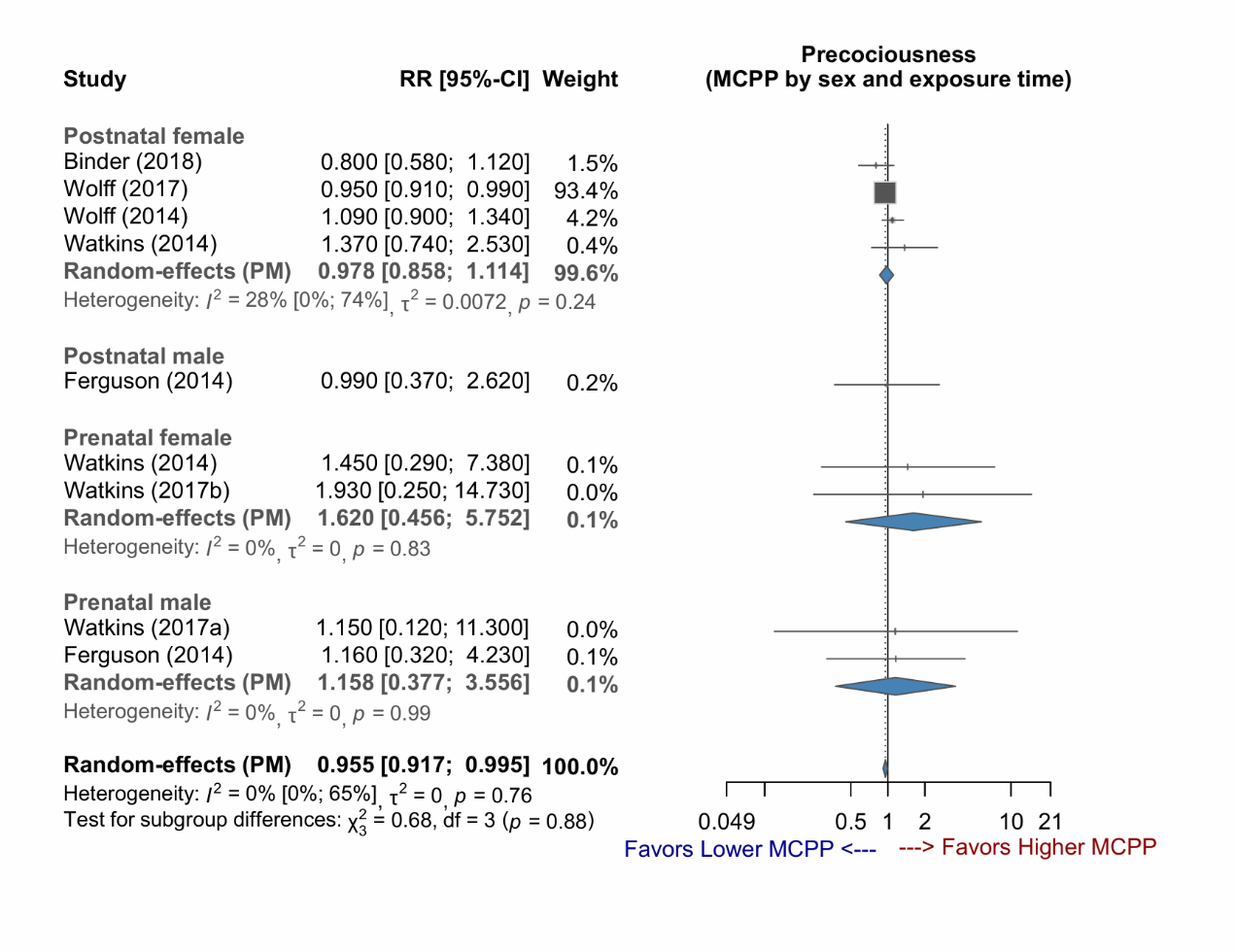

Supplementary Figure 1-19: Forest plot of studies demonstrating the RR of MEHP exposure for early puberty, stratified by sex.

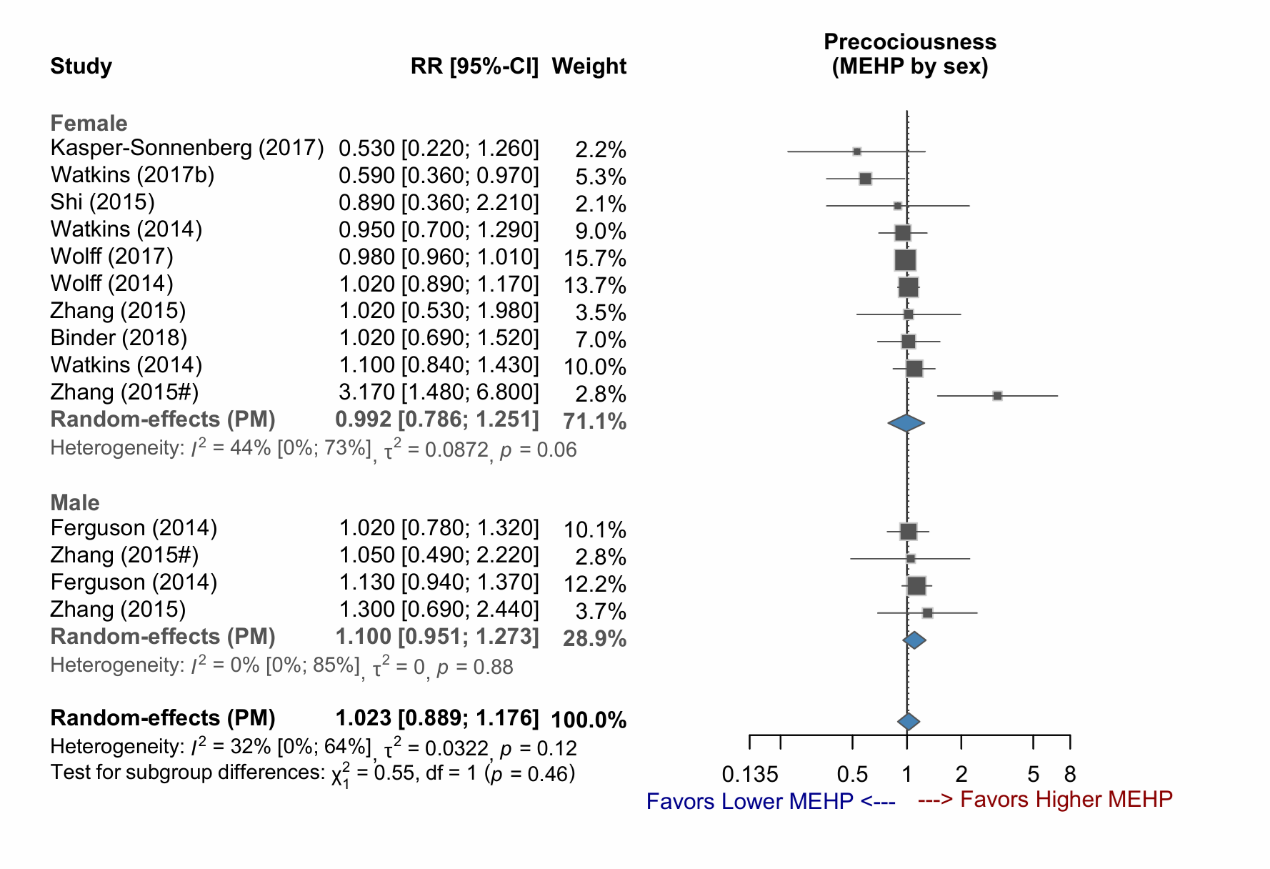

Supplementary Figure 1-20: Forest plot of studies demonstrating the RR of MEHP exposure for early puberty, stratified by exposure time.

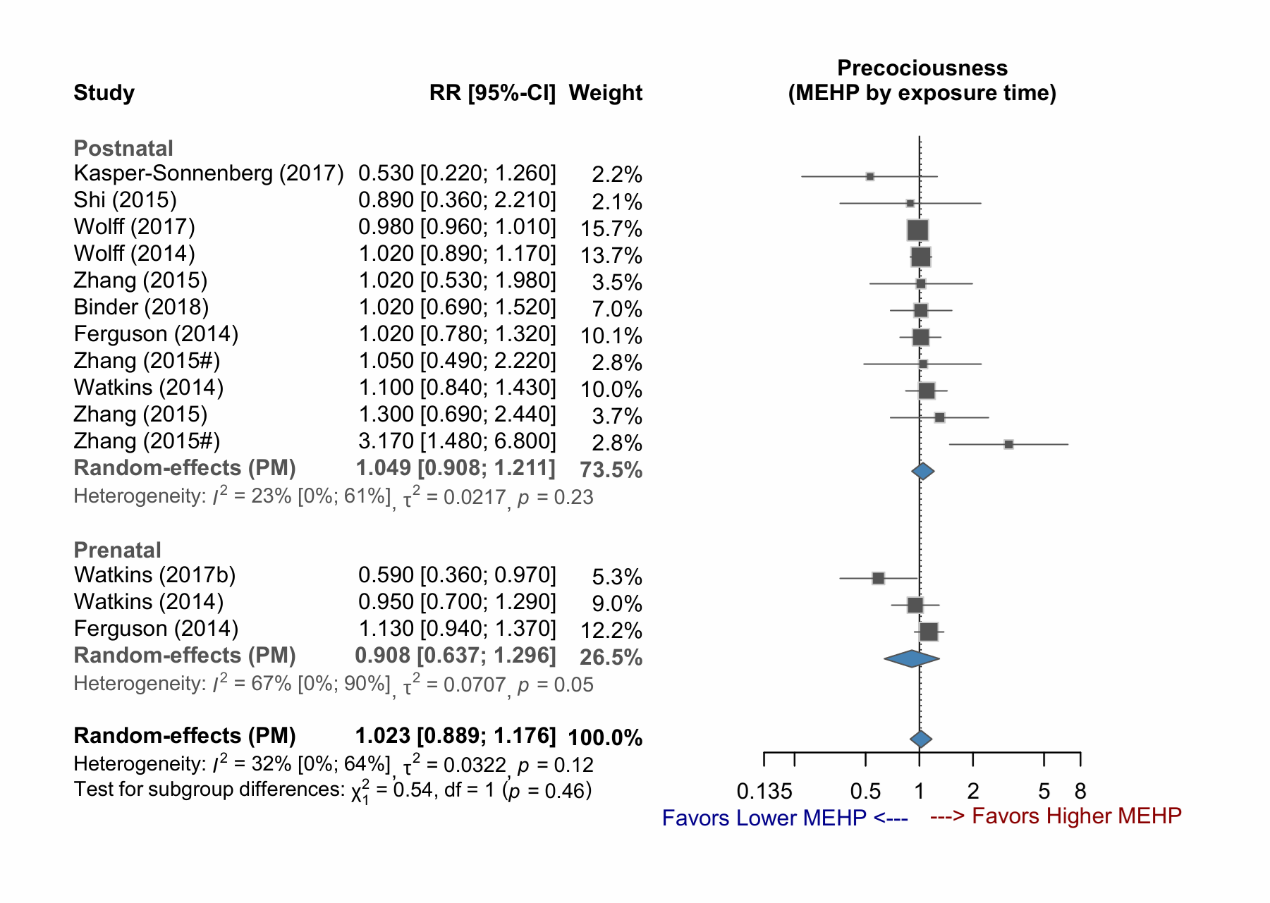

Supplementary Figure 1-21: Forest plot of studies demonstrating the RR of MEHP exposure for early puberty, stratified by sex and exposure time.

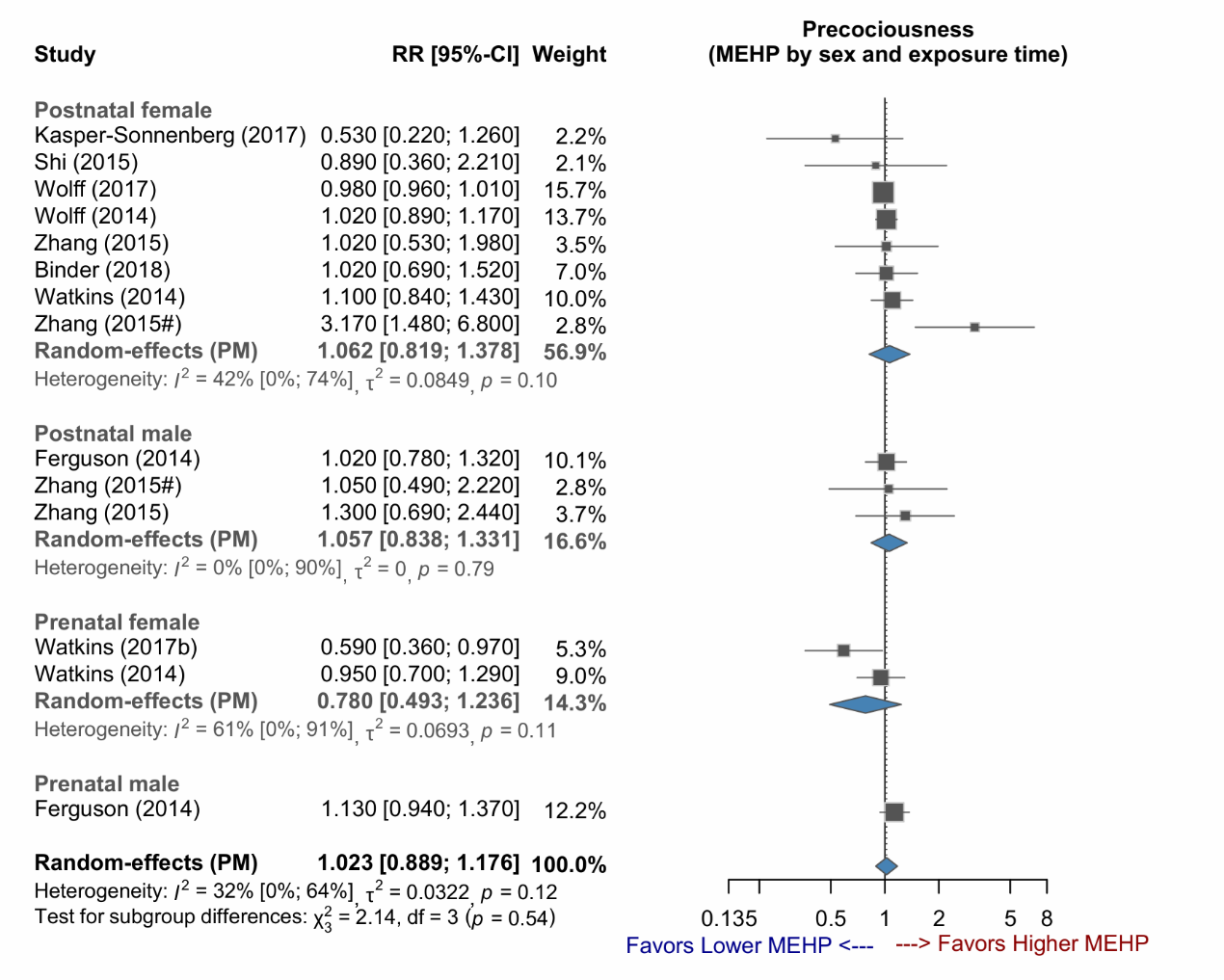

Supplementary Figure 1-22: Forest plot of studies demonstrating the RR of MEP exposure for early puberty, stratified by sex.

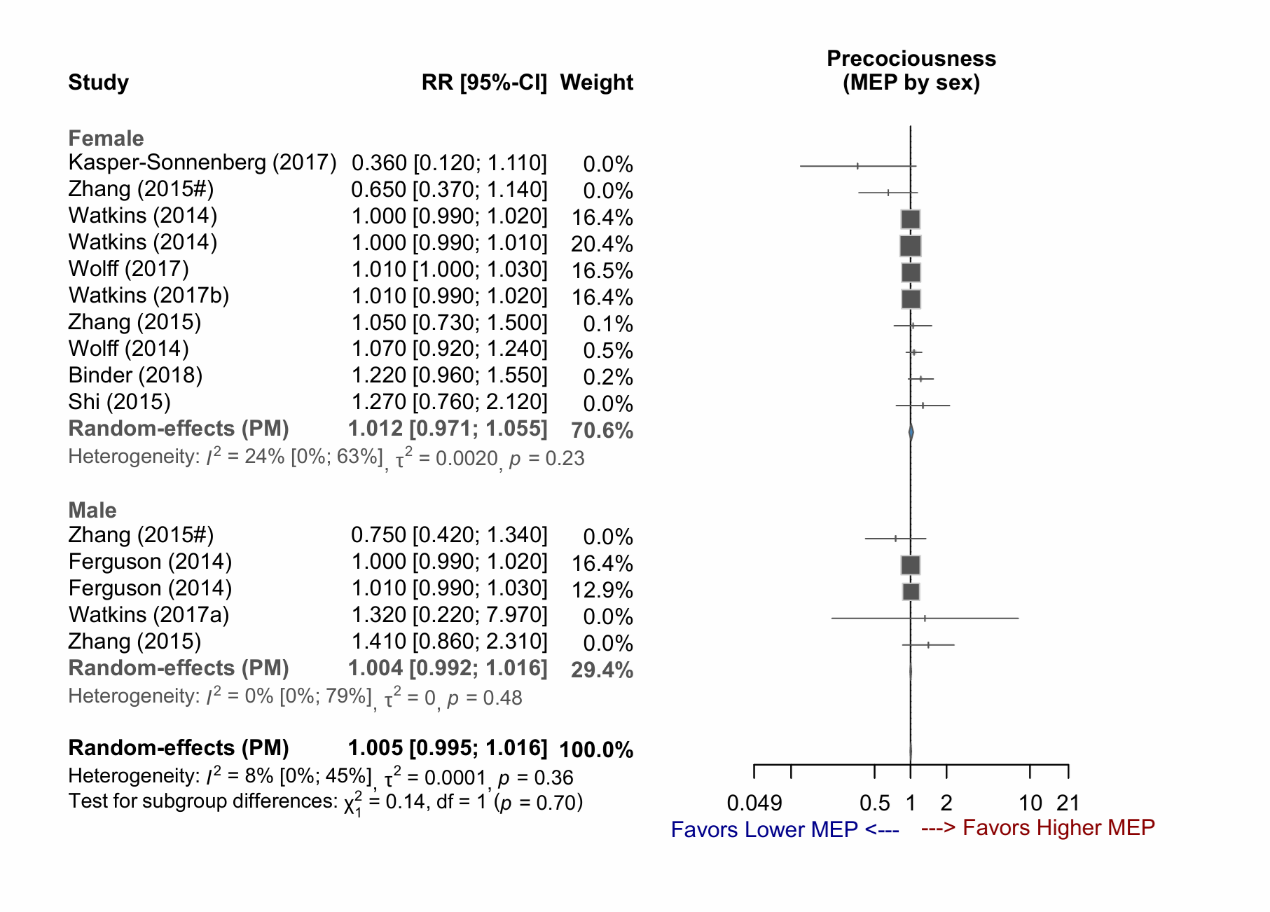

Supplementary Figure 1-23: Forest plot of studies demonstrating the RR of MEP exposure for early puberty, stratified by exposure time.

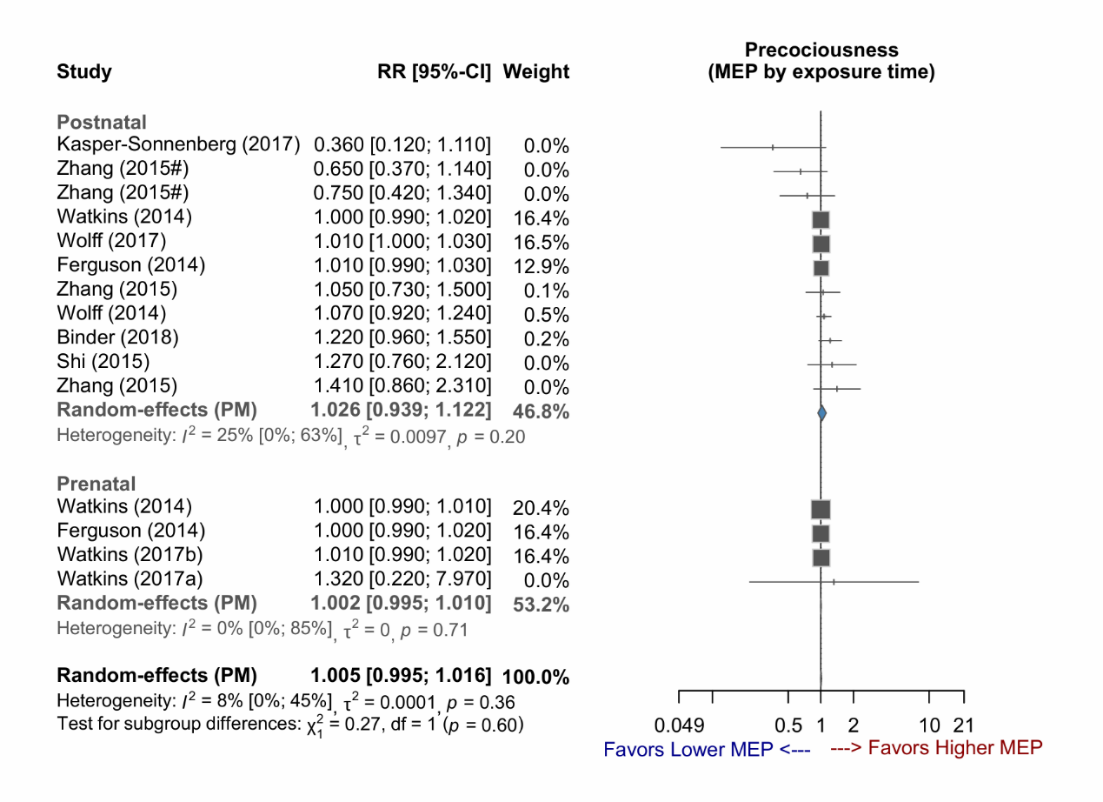

Supplementary Figure 1-24: Forest plot of studies demonstrating the RR of MEP exposure for early puberty, stratified by sex and exposure time.

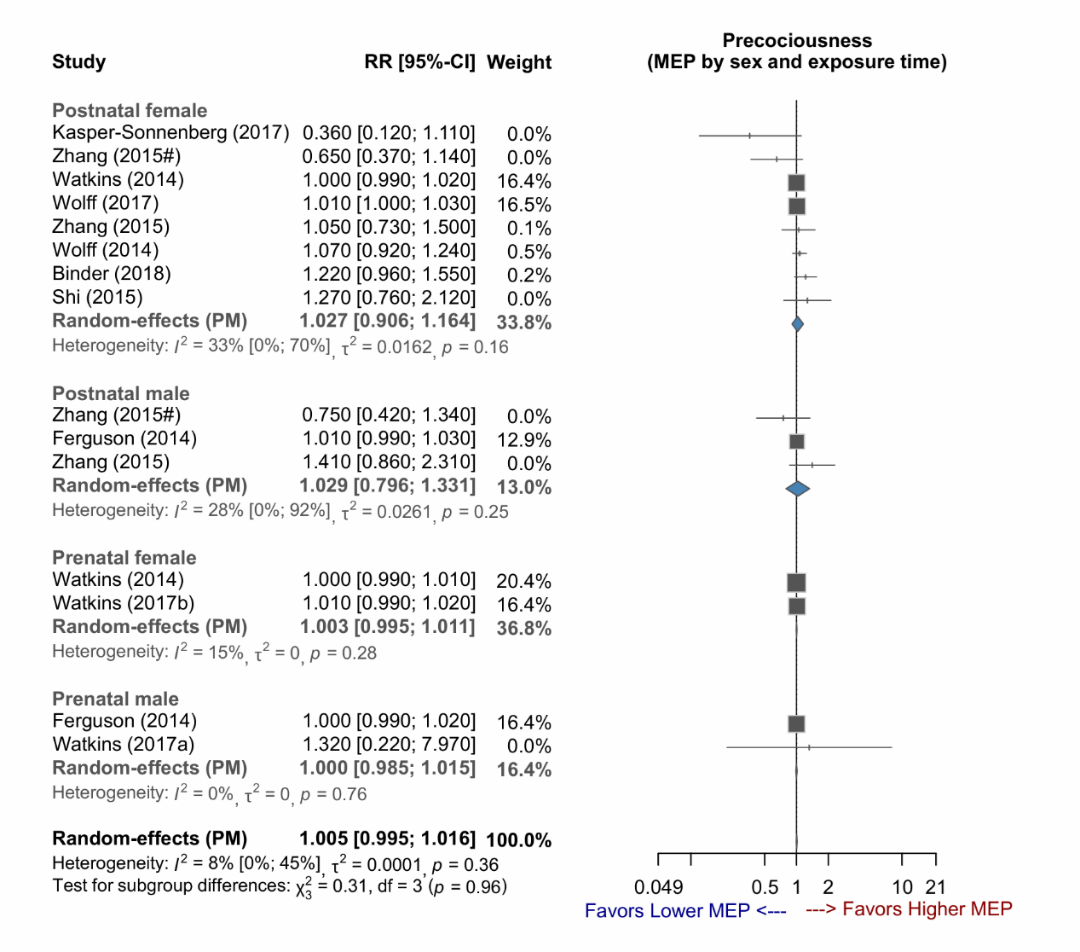

Supplementary Figure 1-25: Forest plot of studies demonstrating the RR of MiBP exposure for early puberty, stratified by sex.

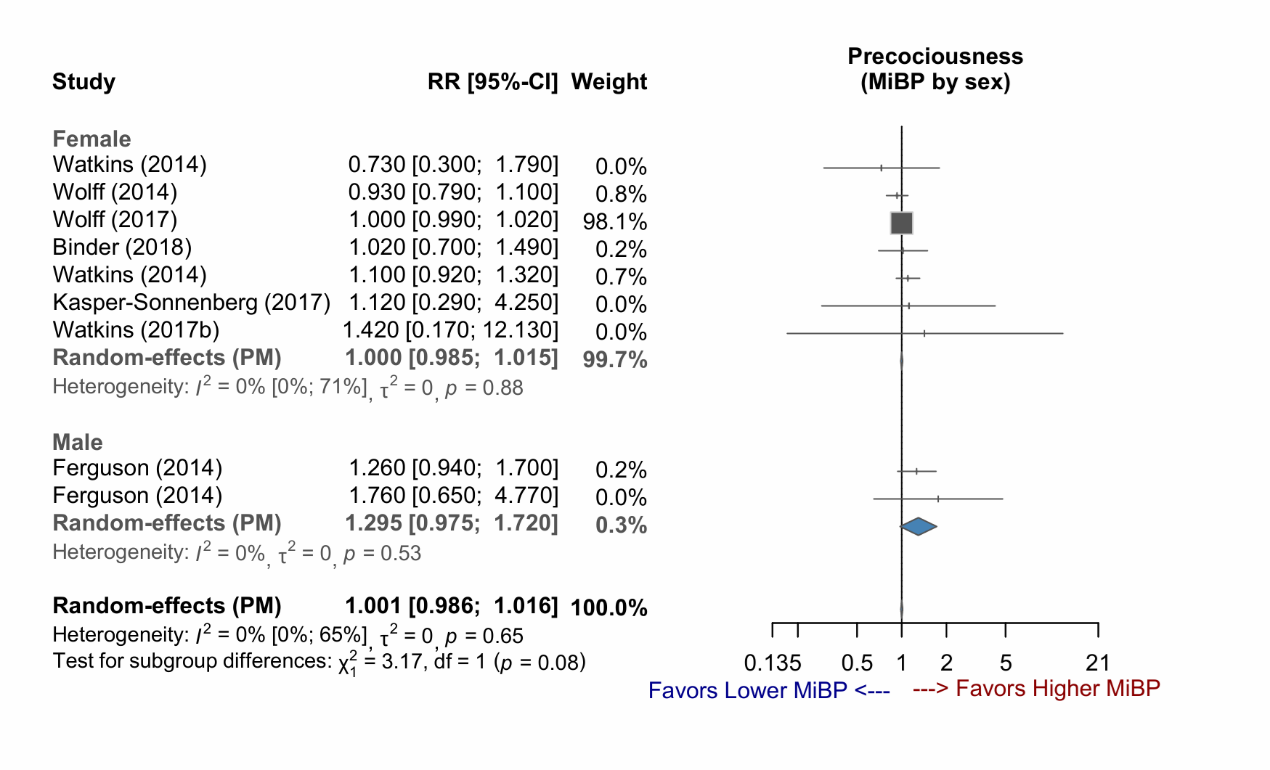

Supplementary Figure 1-26: Forest plot of studies demonstrating the RR of MiBP exposure for early puberty, stratified by exposure time.

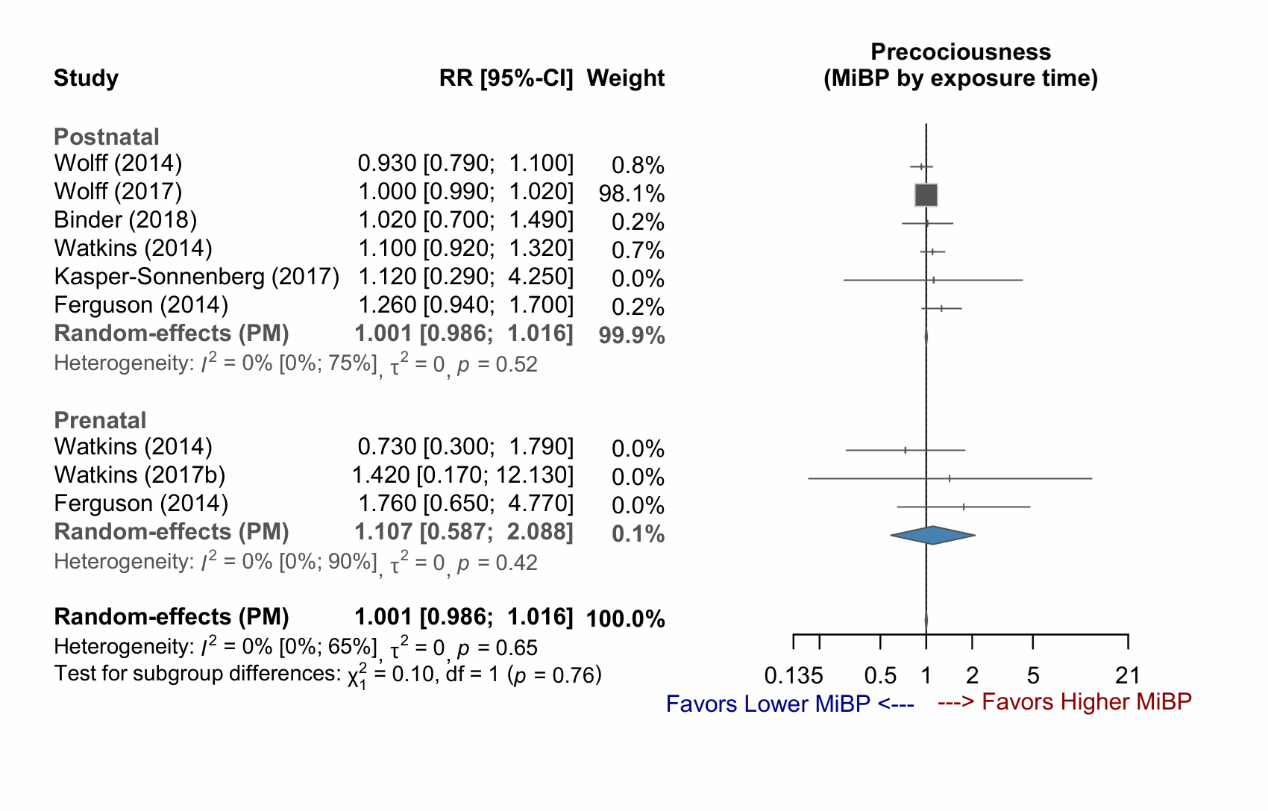

Supplementary Figure 1-27: Forest plot of studies demonstrating the RR of MiBP exposure for early puberty, stratified by sex and exposure time.

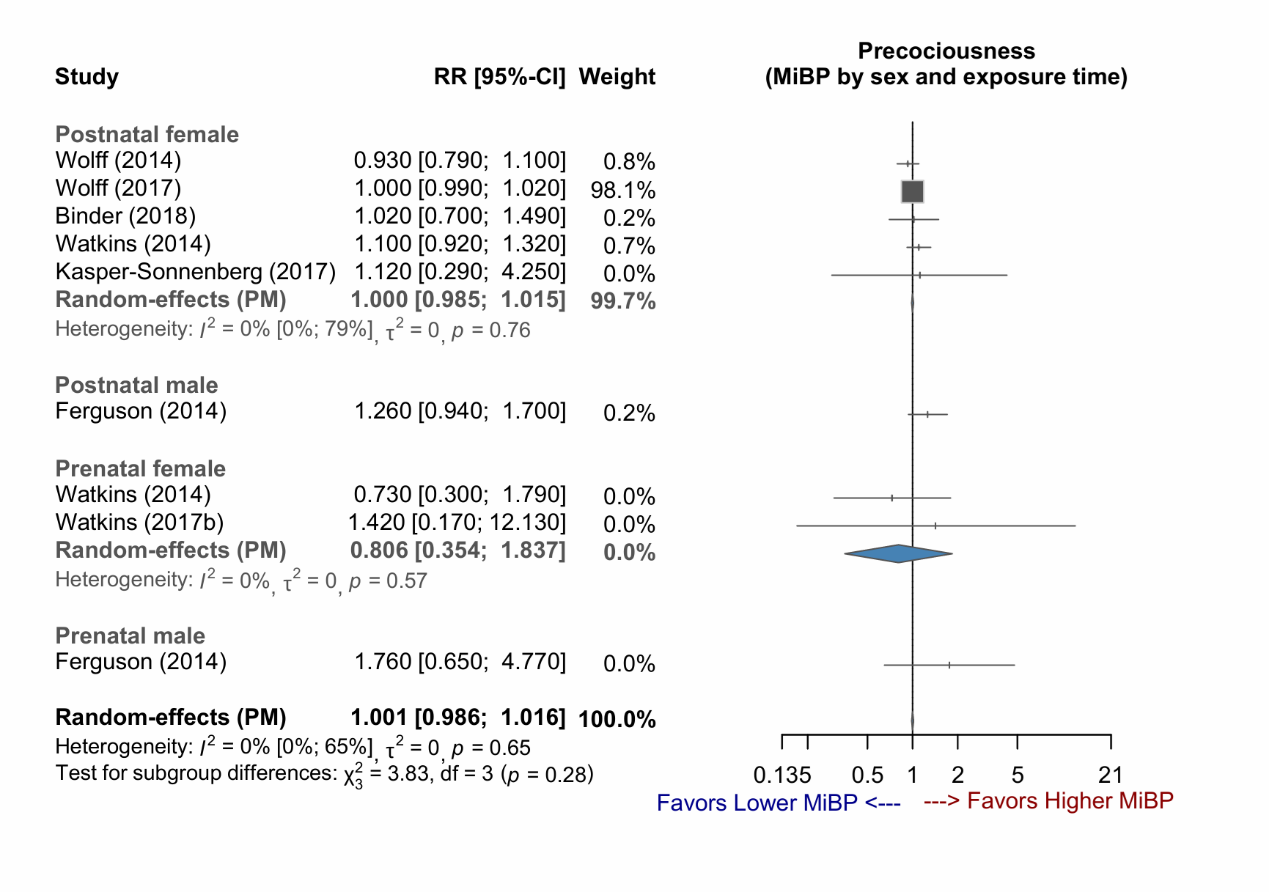

Supplementary Figure 1-28: Forest plot of studies demonstrating the RR of MMP exposure for early puberty, stratified by sex.

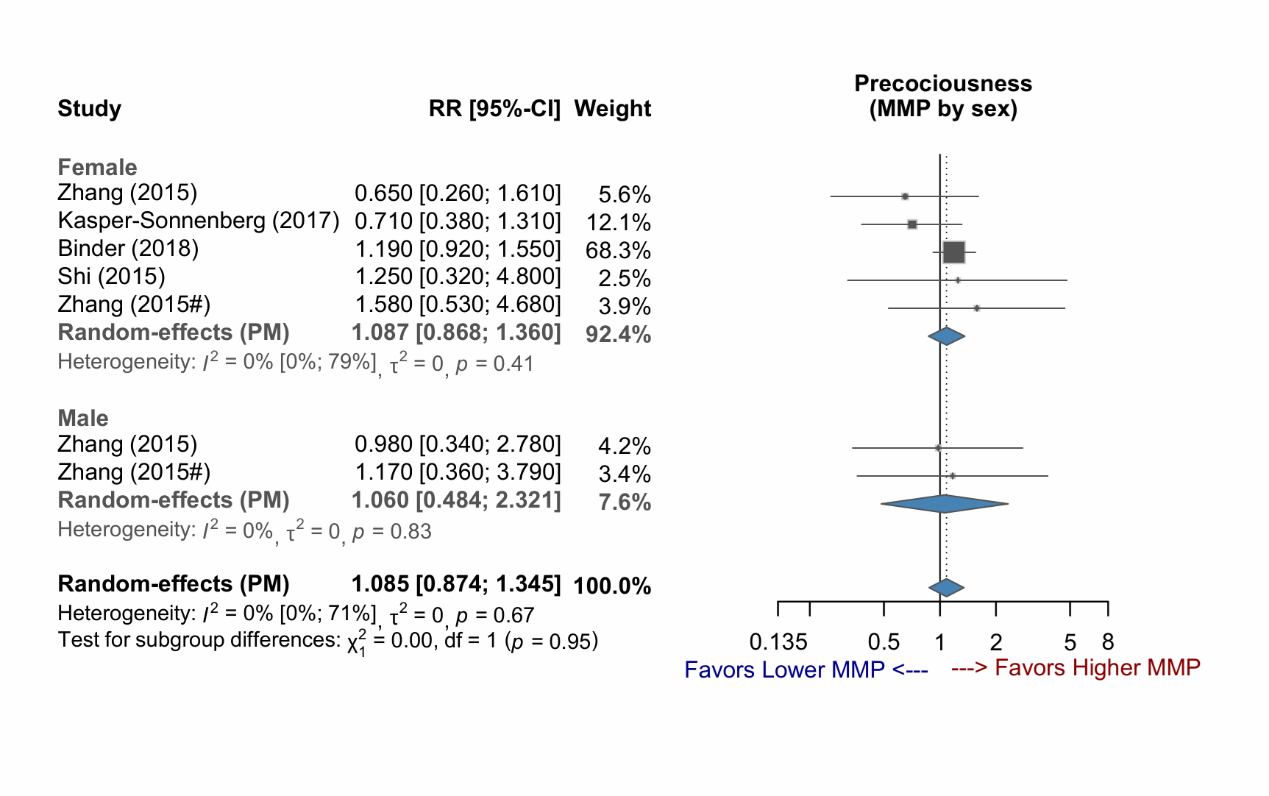

Supplementary Figure 1-29: Forest plot of studies demonstrating the RR of MMP exposure for early puberty.

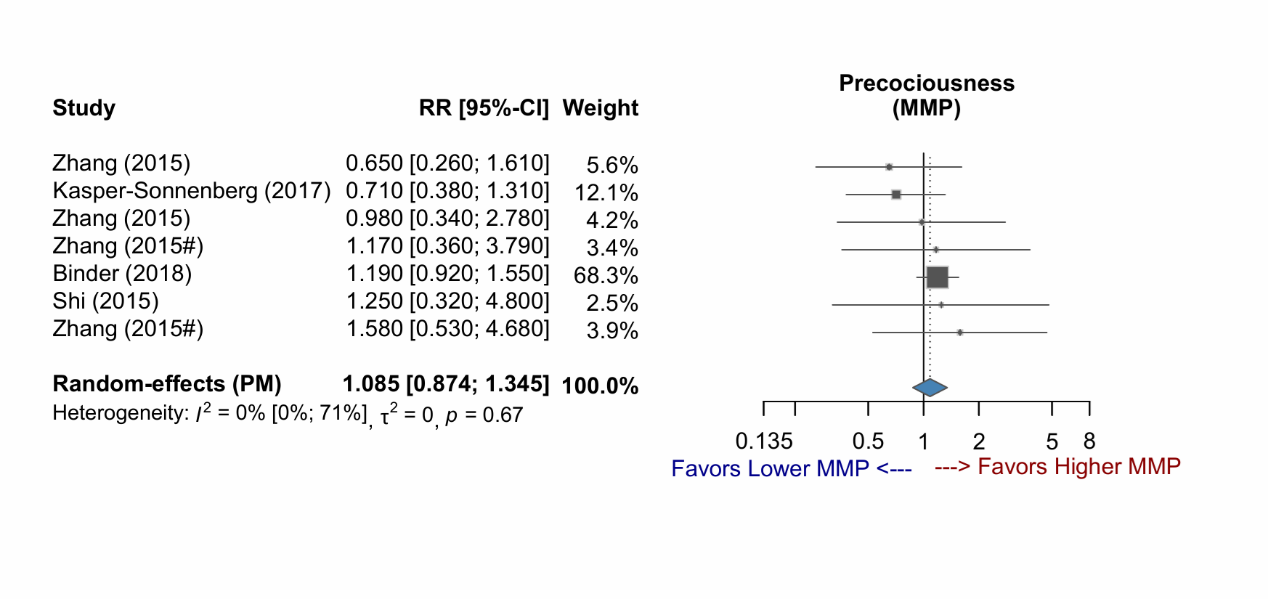

Supplementary Figure 1-30: Forest plot of studies demonstrating the RR of MMP exposure for early puberty, stratified by sex and exposure time.

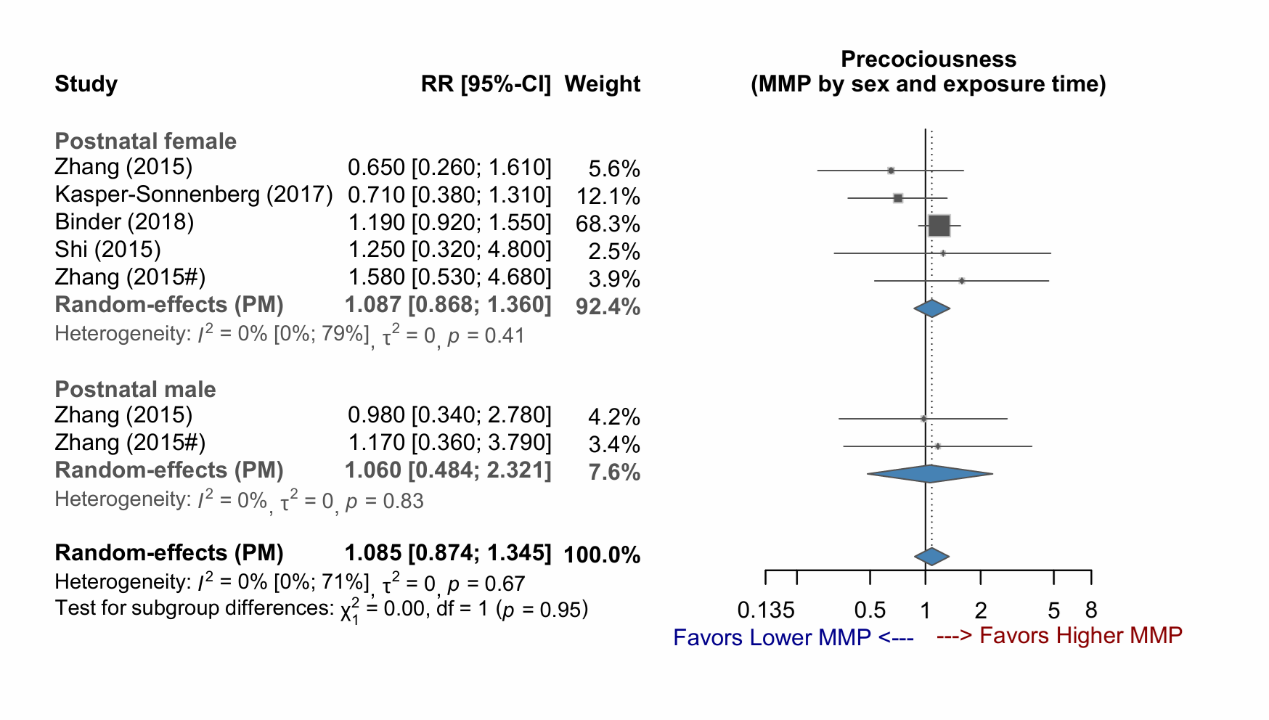

Supplementary Figure 1-31: Forest plot of studies demonstrating the RR of MnBP exposure for early puberty, stratified by sex.

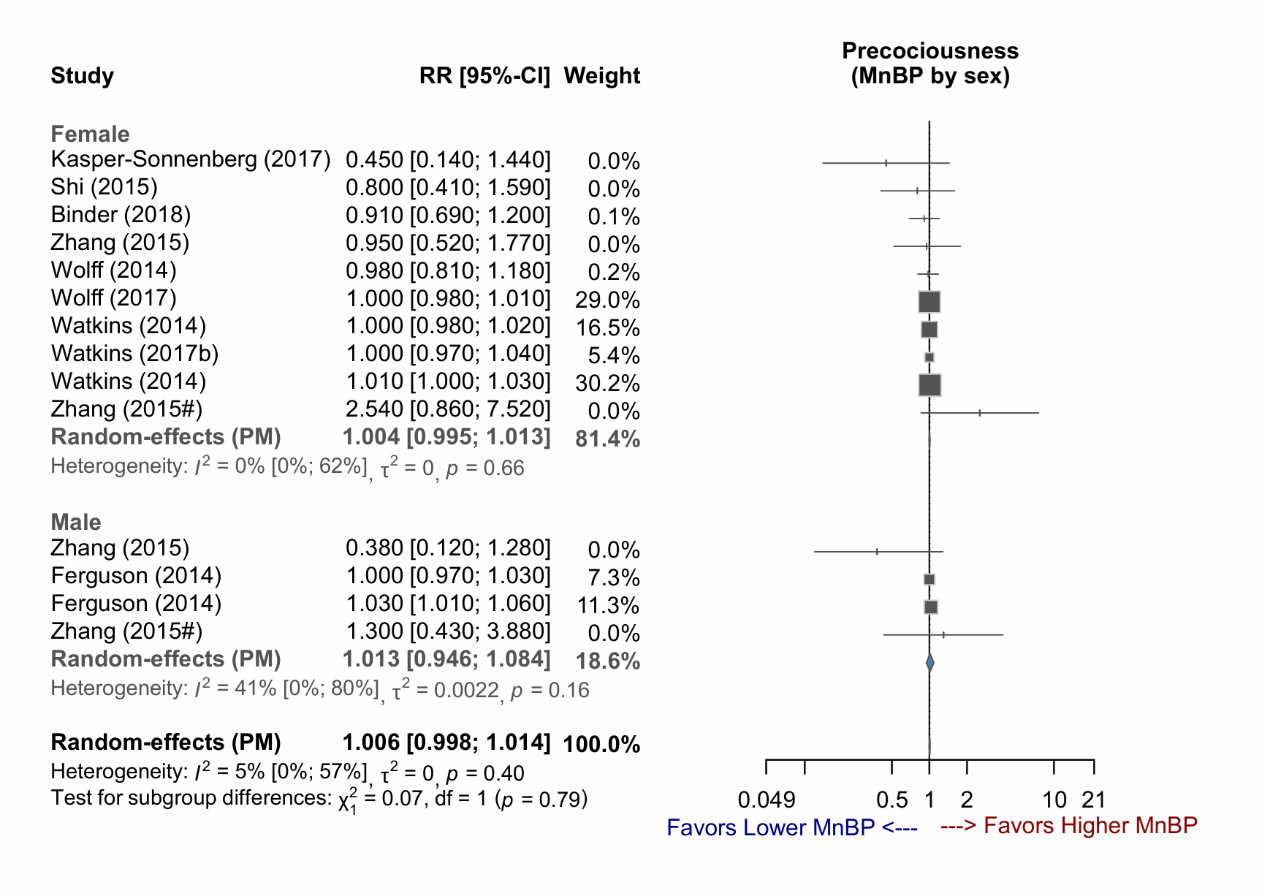

Supplementary Figure 1-32: Forest plot of studies demonstrating the RR of MnBP exposure for early puberty, stratified by exposure time.

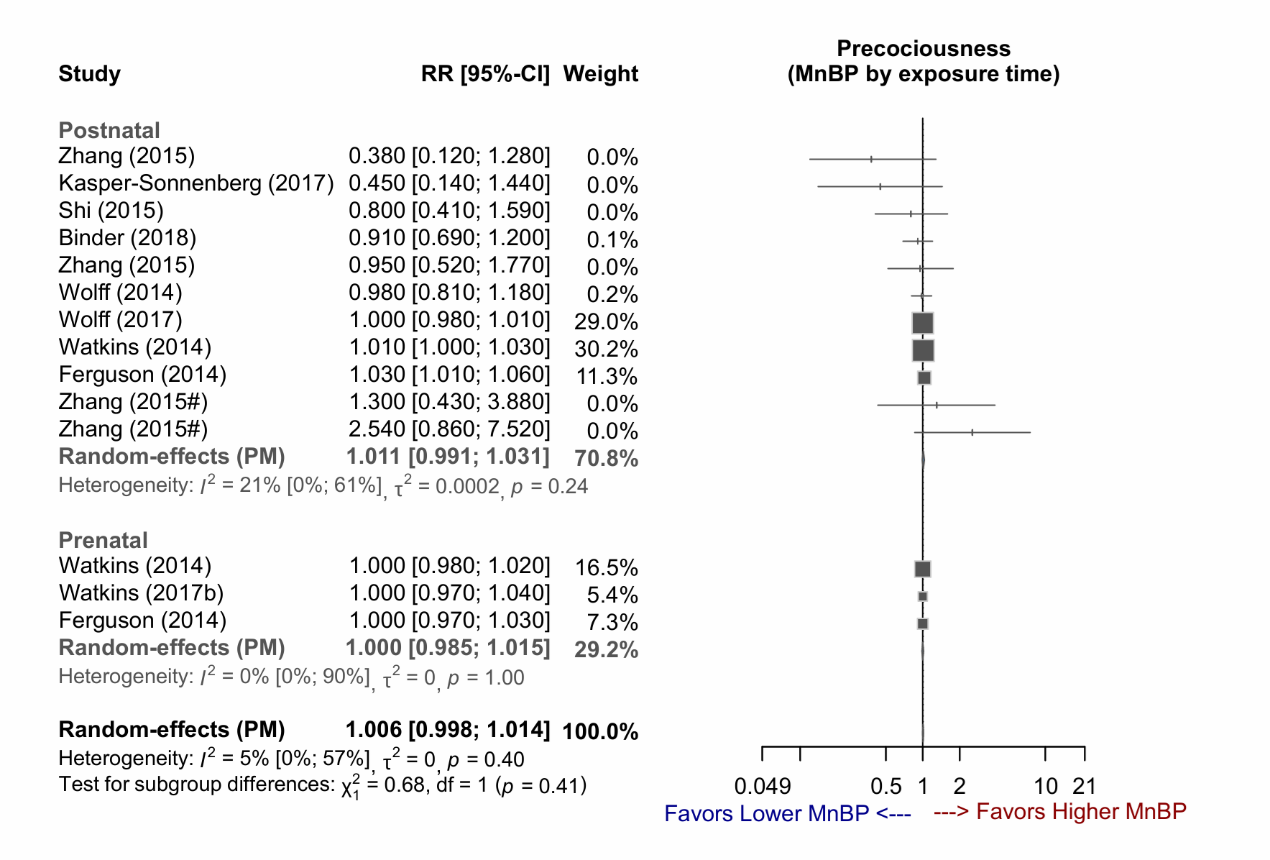

Supplementary Figure 1-33: Forest plot of studies demonstrating the RR of MnBP exposure for early puberty, stratified by sex and exposure time.

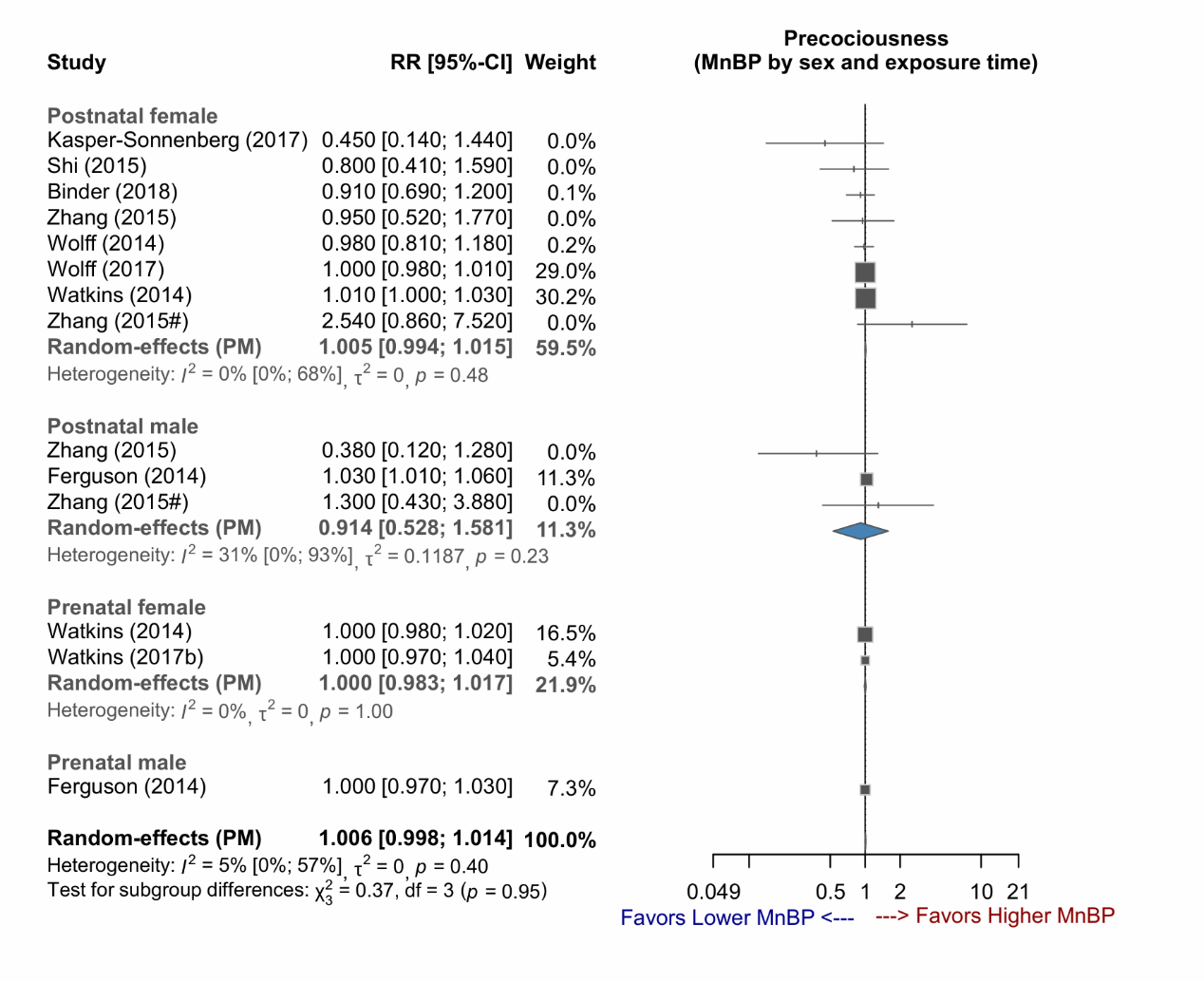

### **Supplementary Figure 2 (2-1~2-18) : Forest plots (adjusted phthalate level for urine-SG)**

Abbreviations:

urine-SG, urine specific gravity; 5OH-MEHP, mono(2-ethyl-5-hydroxyhexyl) phthalate; 5oxo-MEHP, mono(2-ethyl-5-oxohexyl)phthalate; MEHP, mono(2-ethylhexyl) phthalate; MEP, monoethyl phthalate; MMP, monomethyl phthalate; MnBP, mono-n-butyl phthalate

Supplementary Figure 2-1: Forest plot of studies demonstrating the RR of 5OH-MEHP exposure for early puberty, stratified by sex

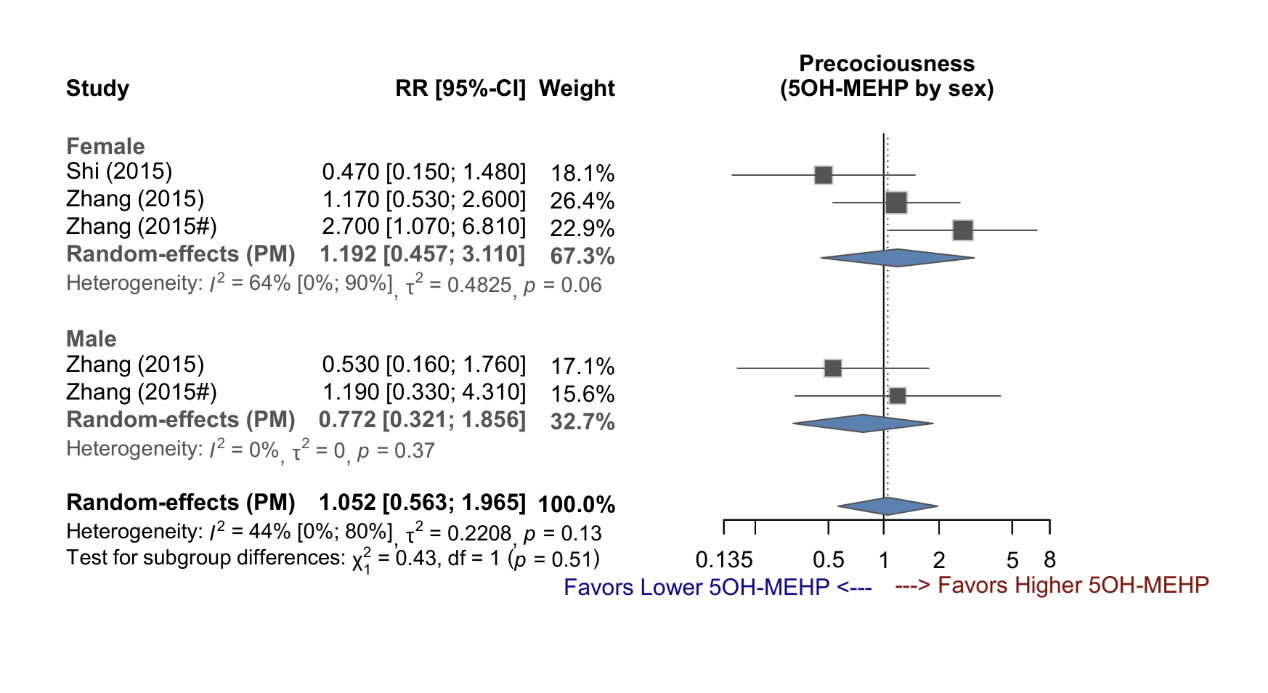

Supplementary Figure 2-2: Forest plot of studies demonstrating the RR of 5OH-MEHP exposure for early puberty.

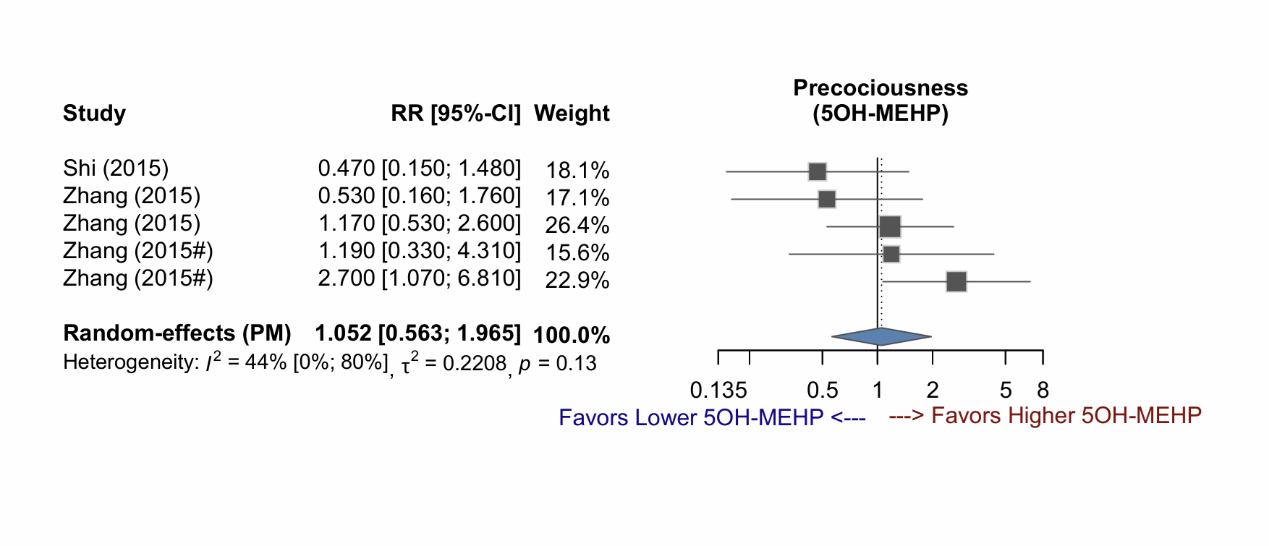

Supplementary Figure 2-3: Forest plot of studies demonstrating the RR of 5OH-MEHP exposure for early puberty, stratified by sex and exposure time.

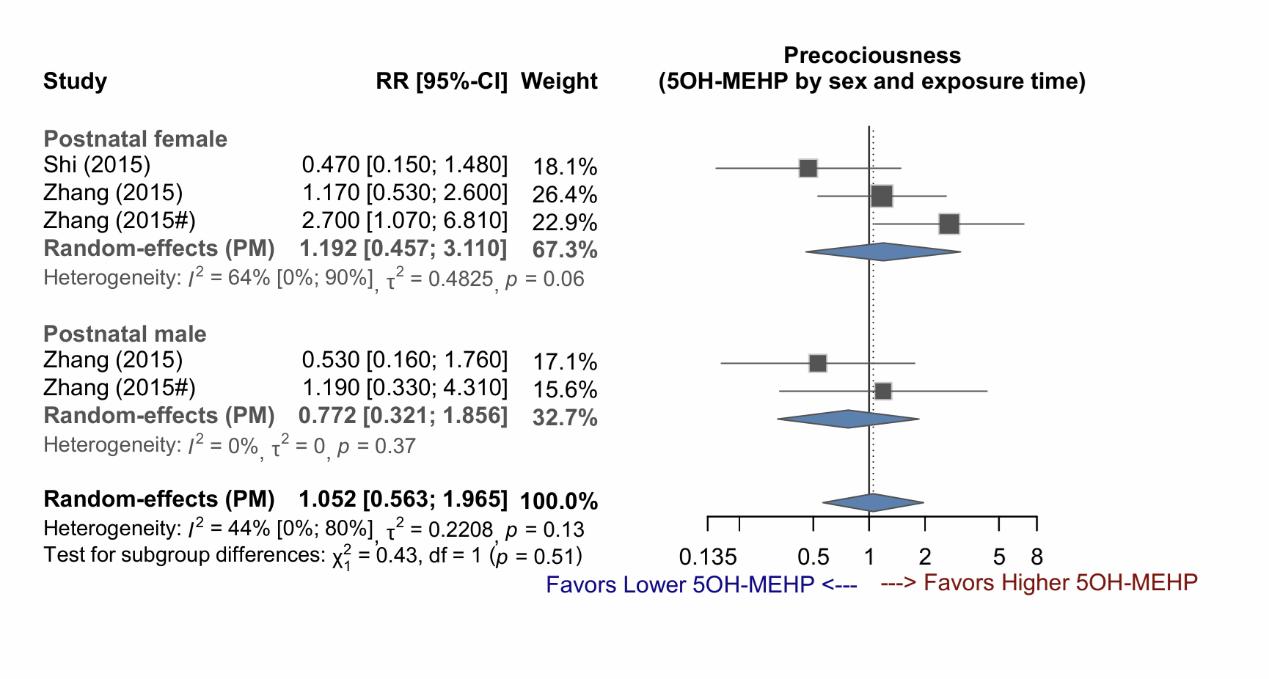

Supplementary Figure 2-4: Forest plot of studies demonstrating the RR of 5oxo-MEHP exposure for early puberty, stratified by sex.

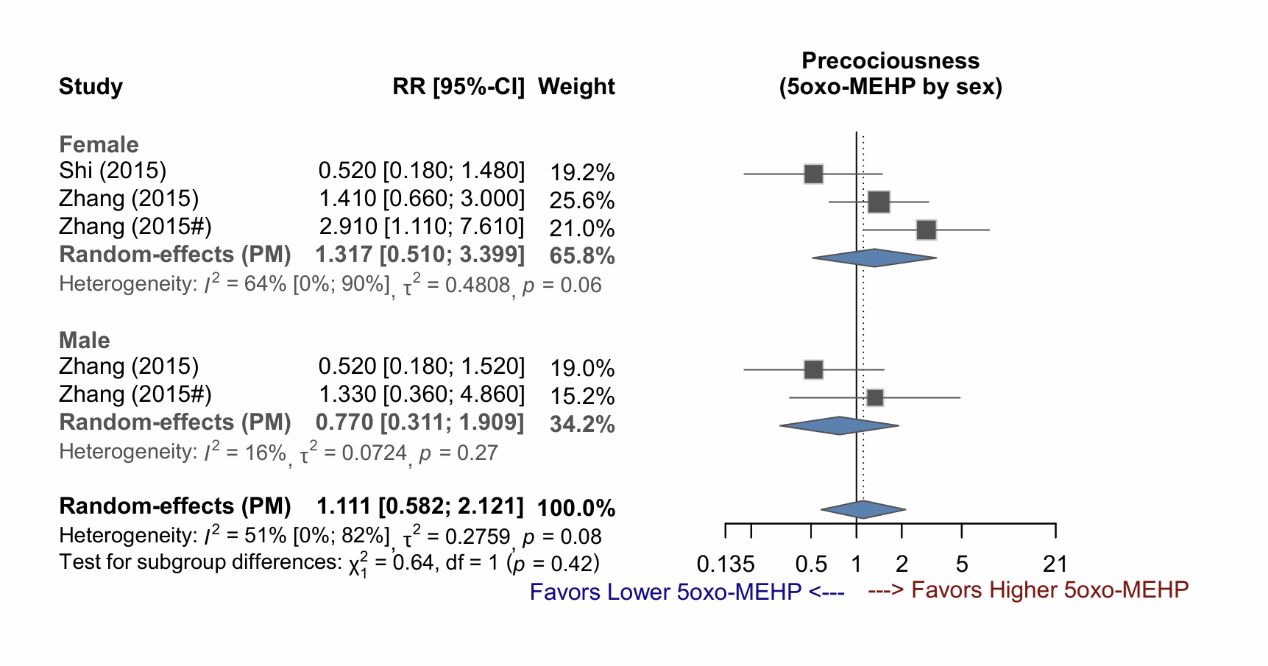

Supplementary Figure 2-5: Forest plot of studies demonstrating the RR of 5oxo-MEHP exposure for early puberty.

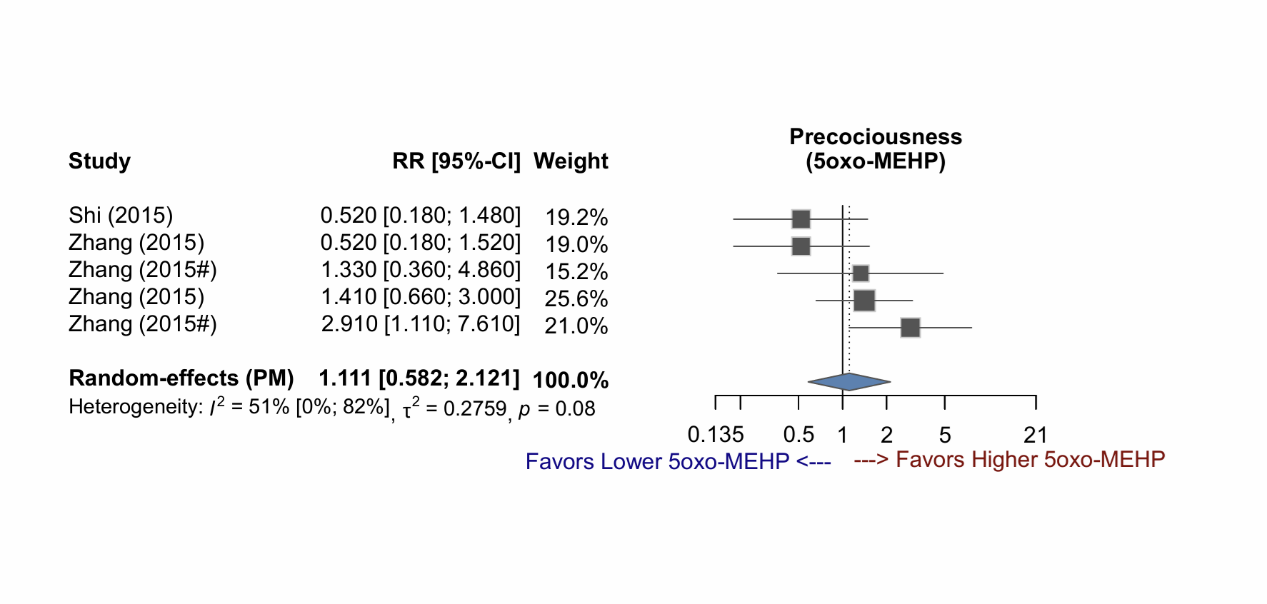

Supplementary Figure 2-6: Forest plot of studies demonstrating the RR of 5oxo-MEHP exposure for early puberty, stratified by sex and exposure time.

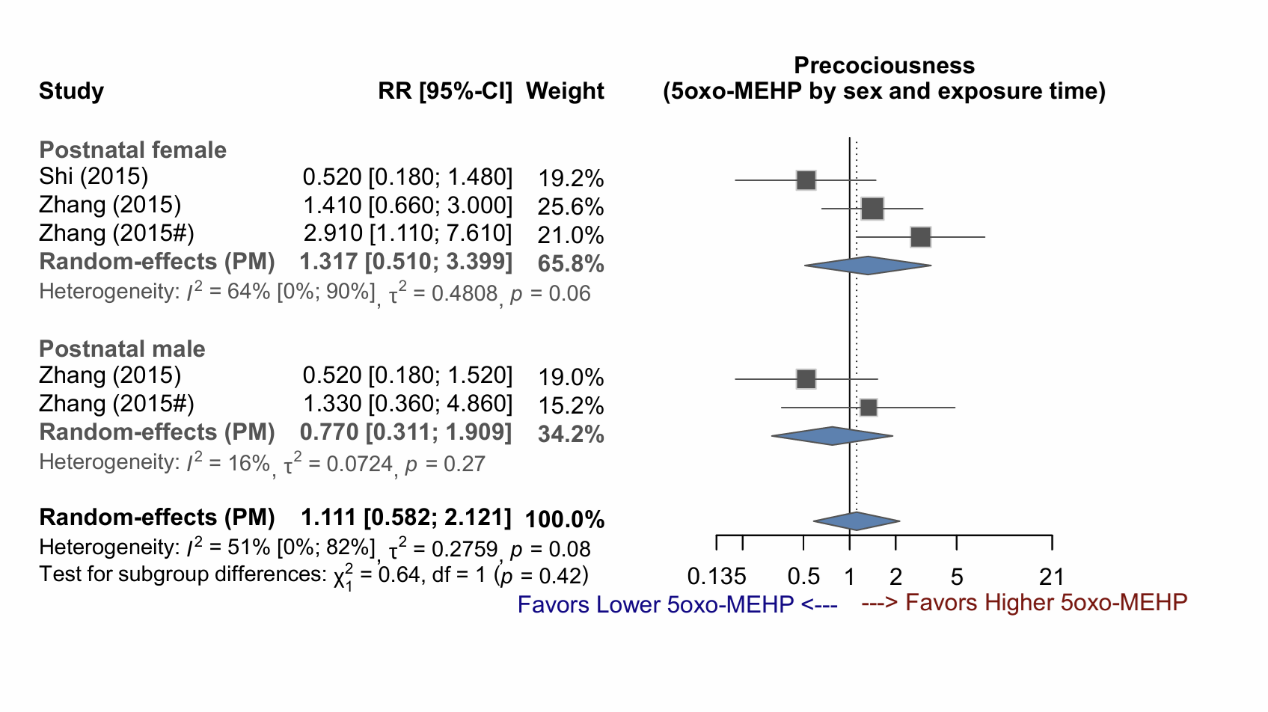

Supplementary Figure 2-7: Forest plot of studies demonstrating the RR of MEHP exposure for early puberty, stratified by sex.

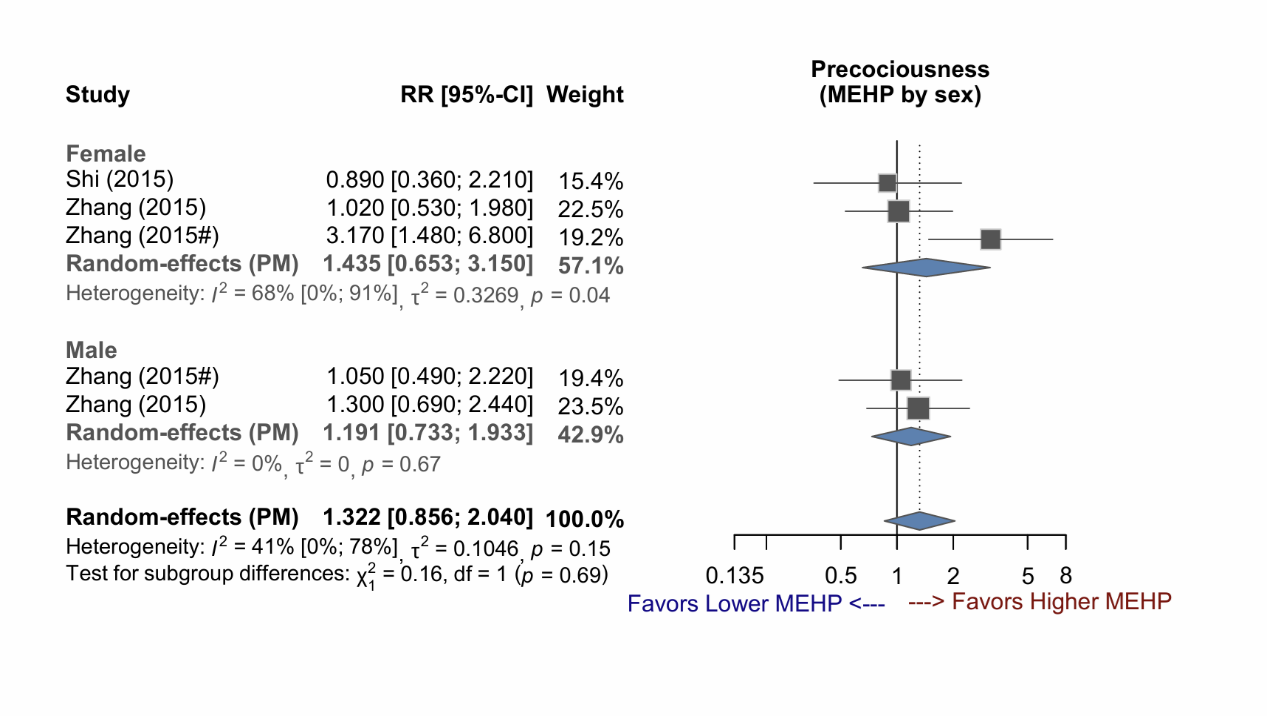

Supplementary Figure 2-8: Forest plot of studies demonstrating the RR of MEHP exposure for early puberty.

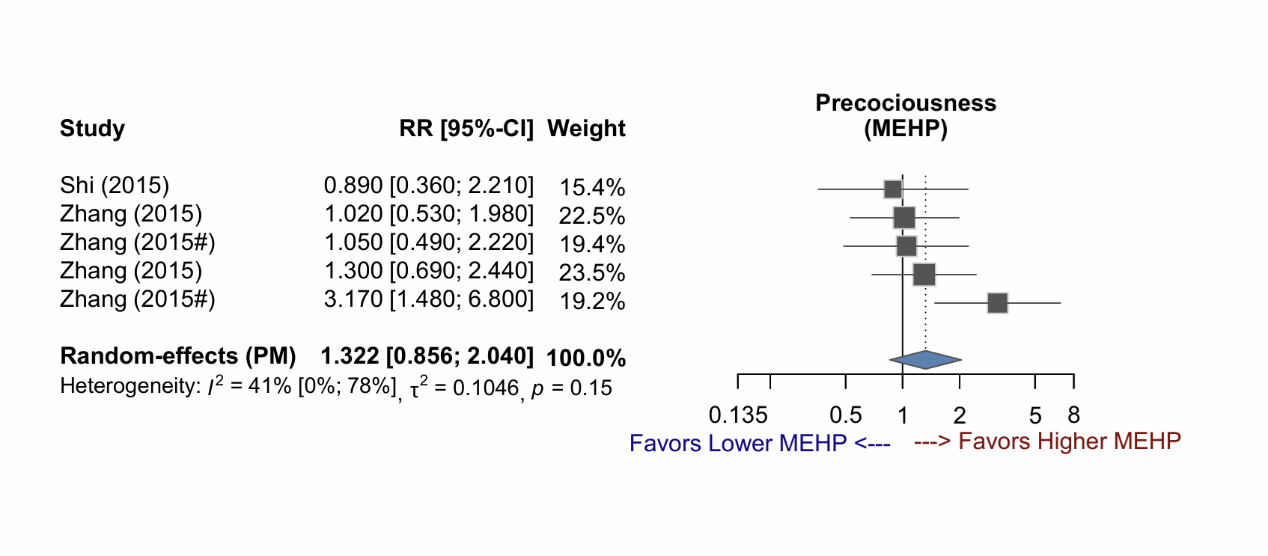

Supplementary Figure 2-9: Forest plot of studies demonstrating the RR of MEHP exposure for early puberty, stratified by sex and exposure time.

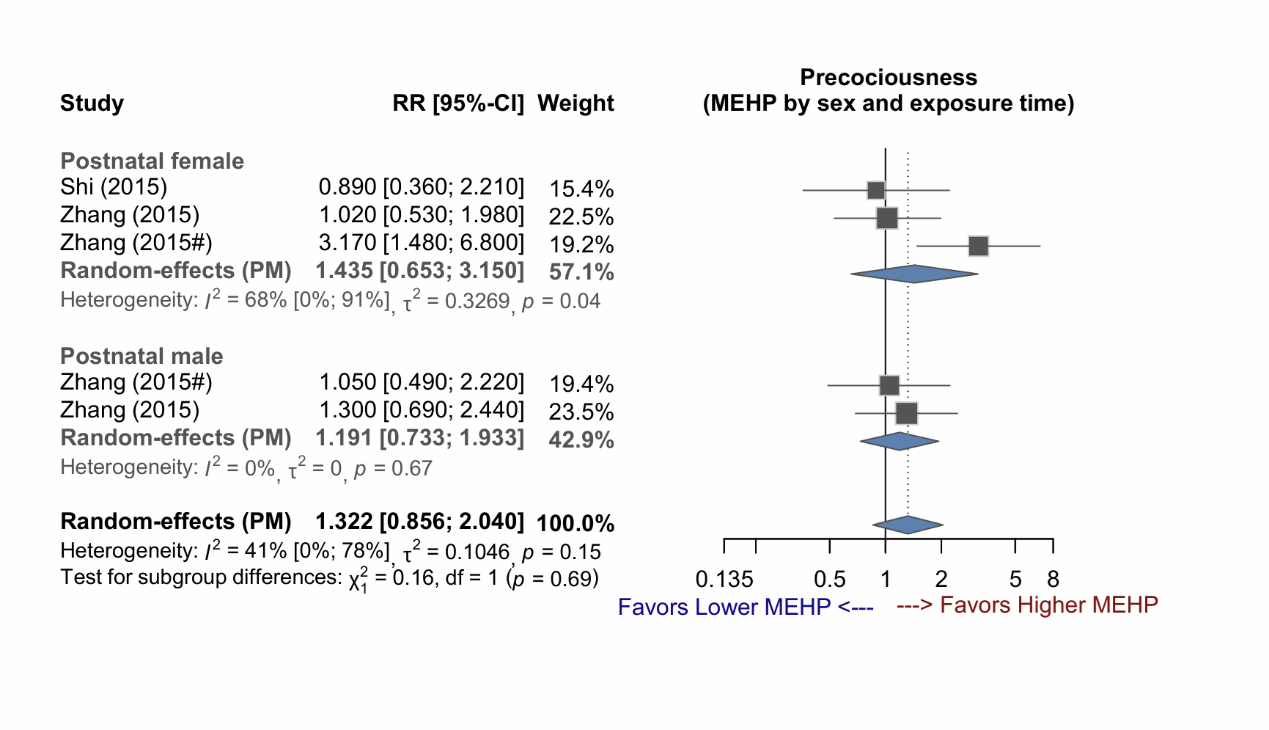

Supplementary Figure 2-10: Forest plot of studies demonstrating the RR of MEP exposure for early puberty, stratified by sex.

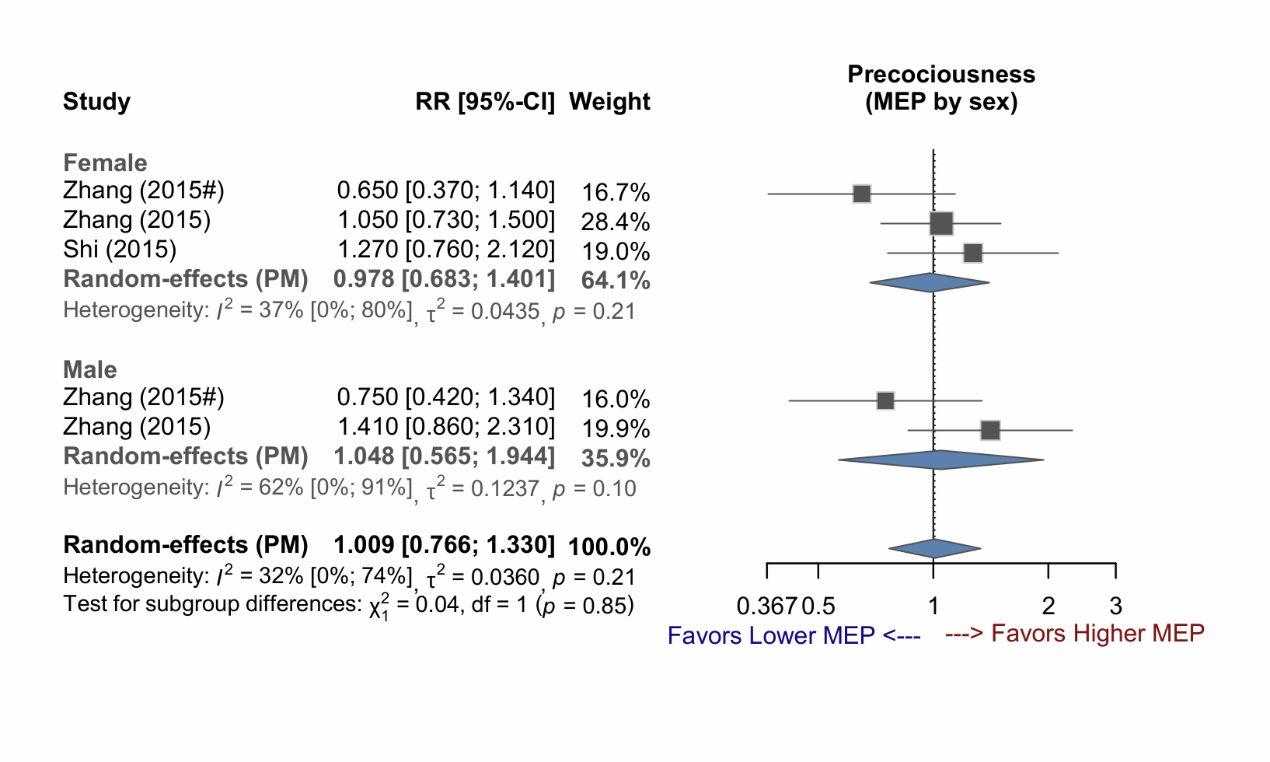

Supplementary Figure 2-11: Forest plot of studies demonstrating the RR of MEP exposure for early puberty.

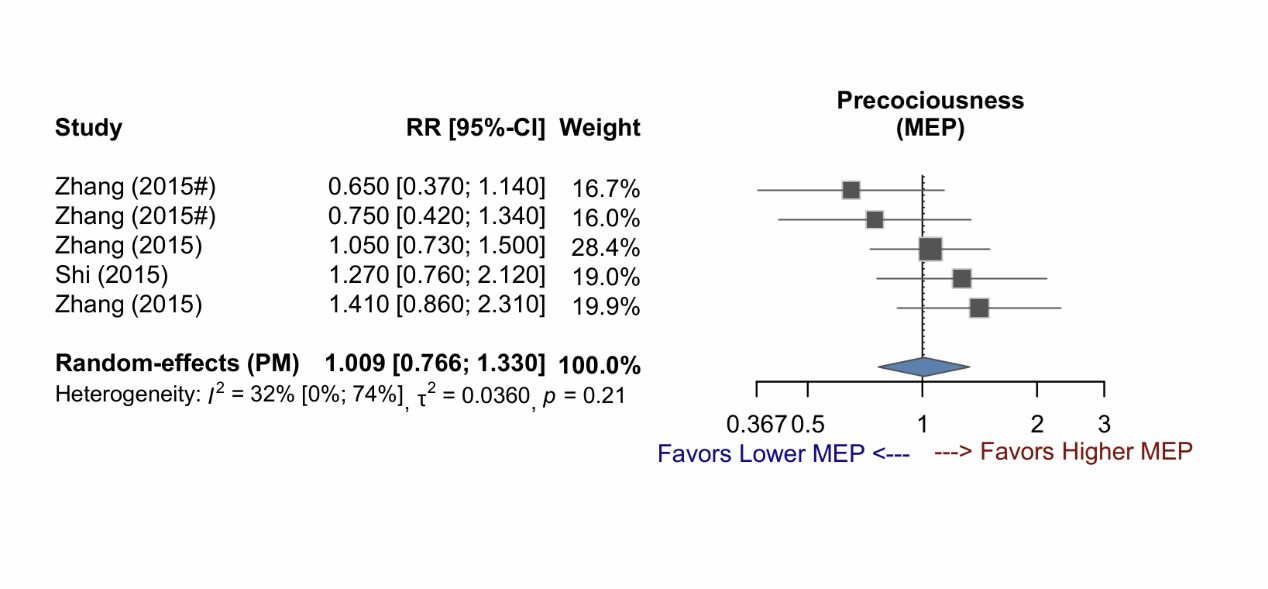

Supplementary Figure 2-12: Forest plot of studies demonstrating the RR of MEP exposure for early puberty, stratified by sex and exposure time.

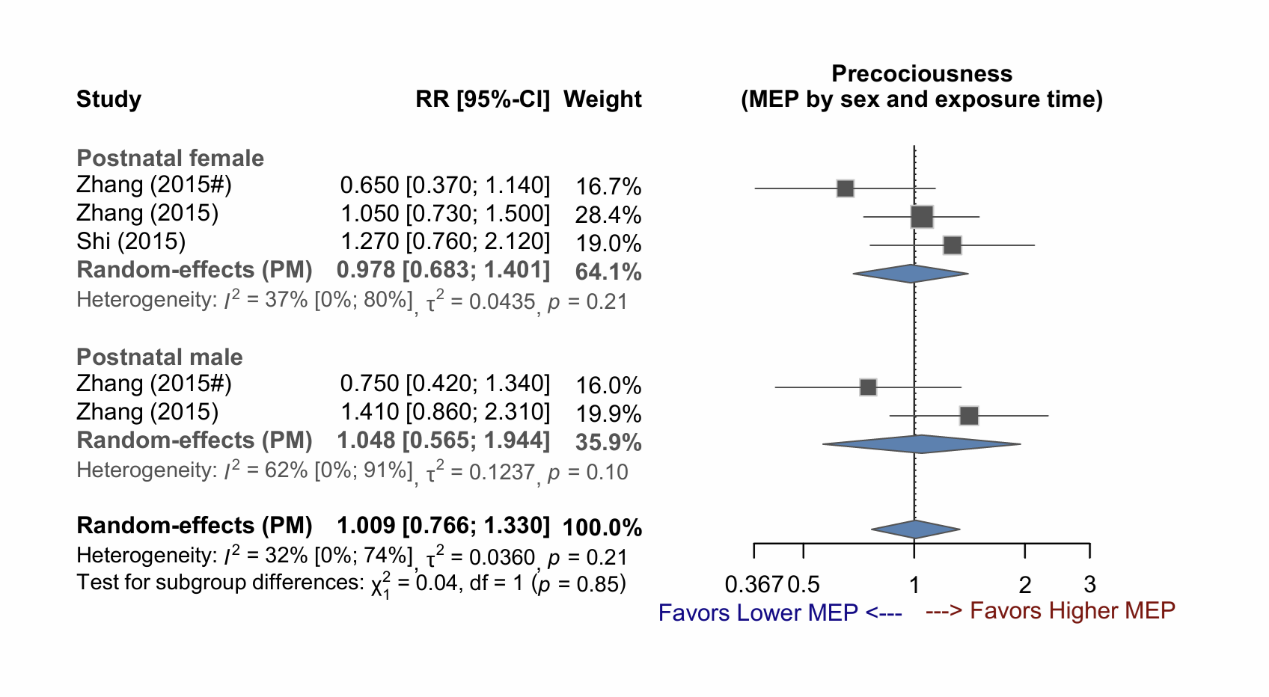

Supplementary Figure 2-13: Forest plot of studies demonstrating the RR of MMP exposure for early puberty, stratified by sex.

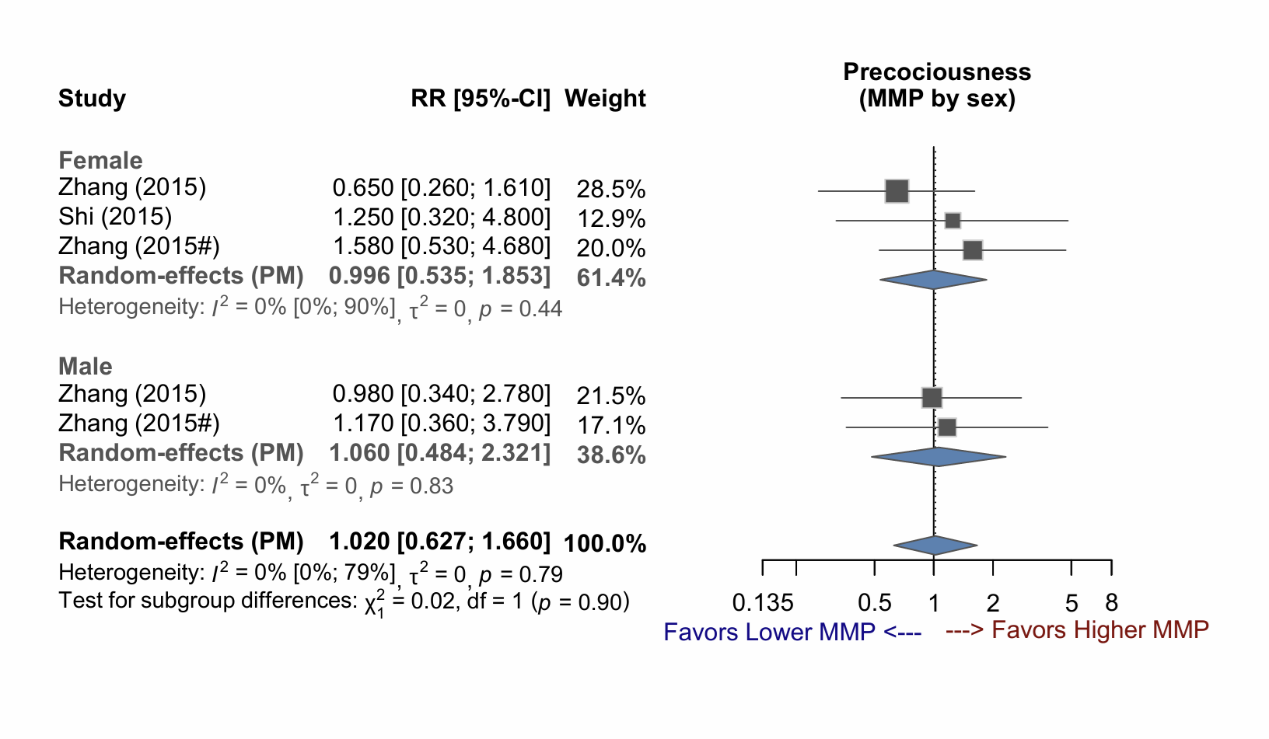

Supplementary Figure 2-14: Forest plot of studies demonstrating the RR of MMP exposure for early puberty.

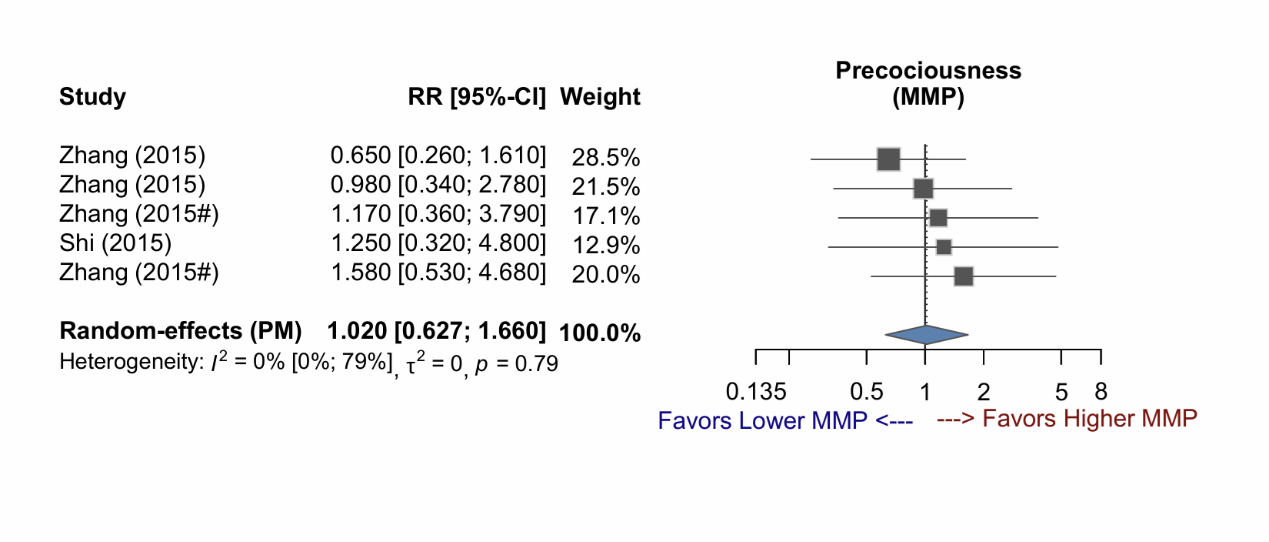

Supplementary Figure 2-15: Forest plot of studies demonstrating the RR of MMP exposure for early puberty, stratified by sex and exposure time.

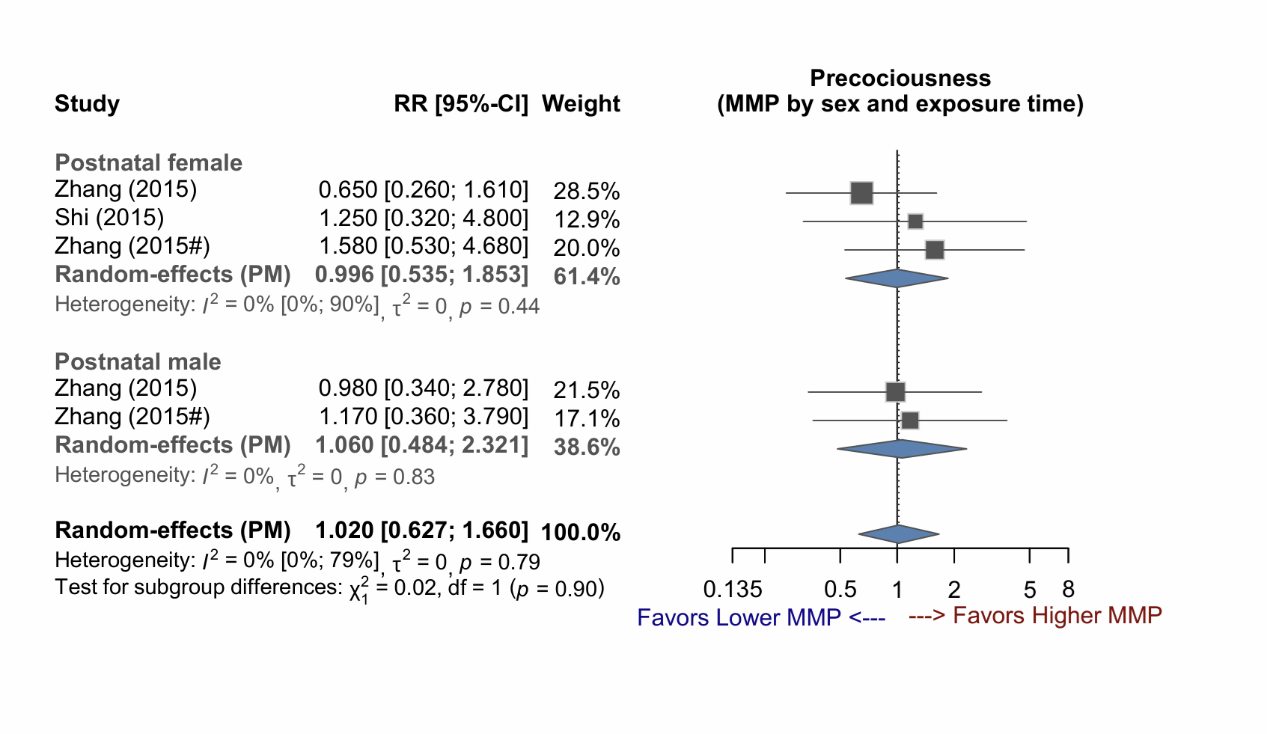

Supplementary Figure 2-16: Forest plot of studies demonstrating the RR of MnBP exposure for early puberty, stratified by sex.

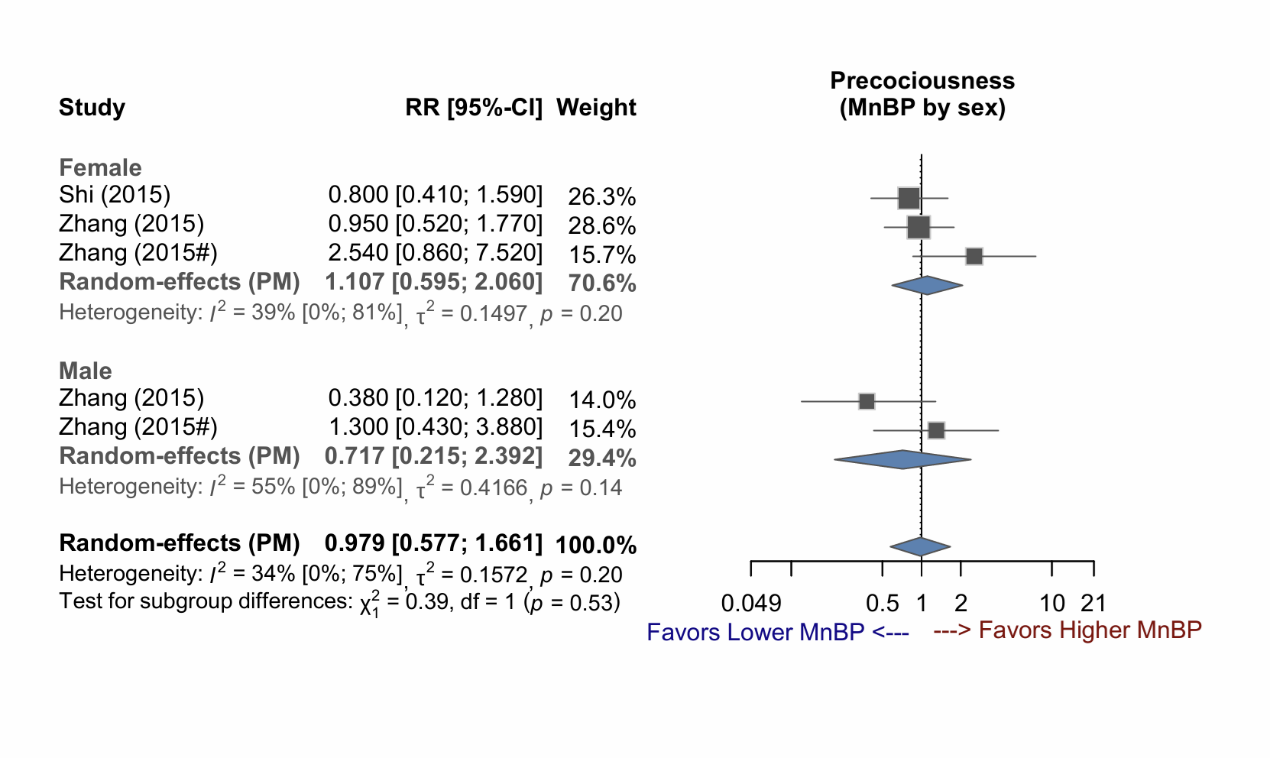

Supplementary Figure 2-17: Forest plot of studies demonstrating the RR of MnBP exposure for early puberty.

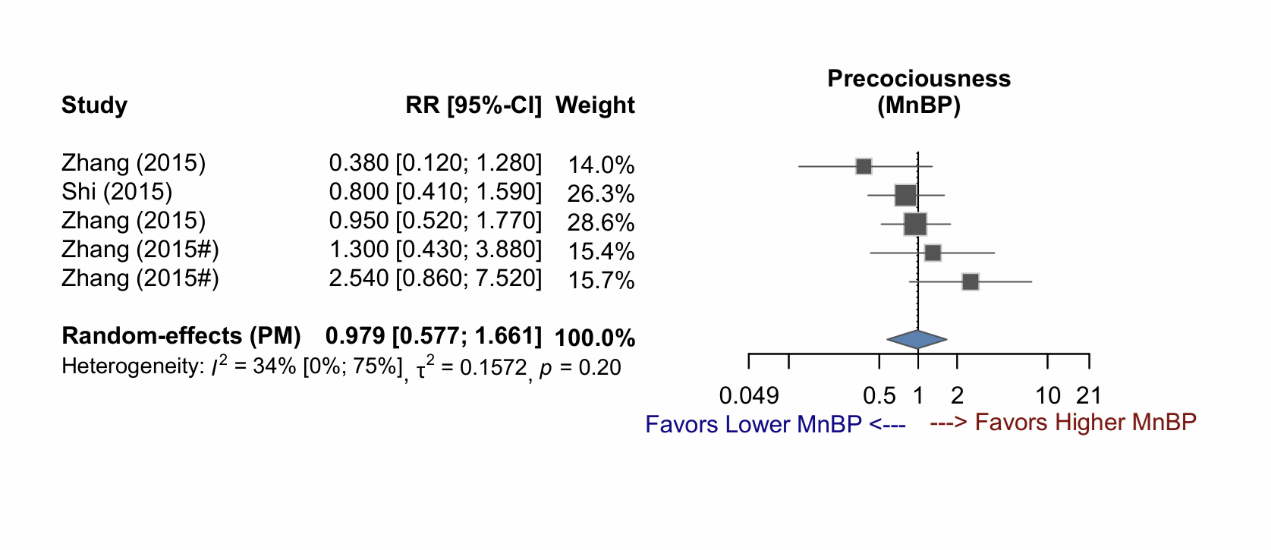

Supplementary Figure 2-18: Forest plot of studies demonstrating the RR of MnBP exposure for early puberty, stratified by sex and exposure time.

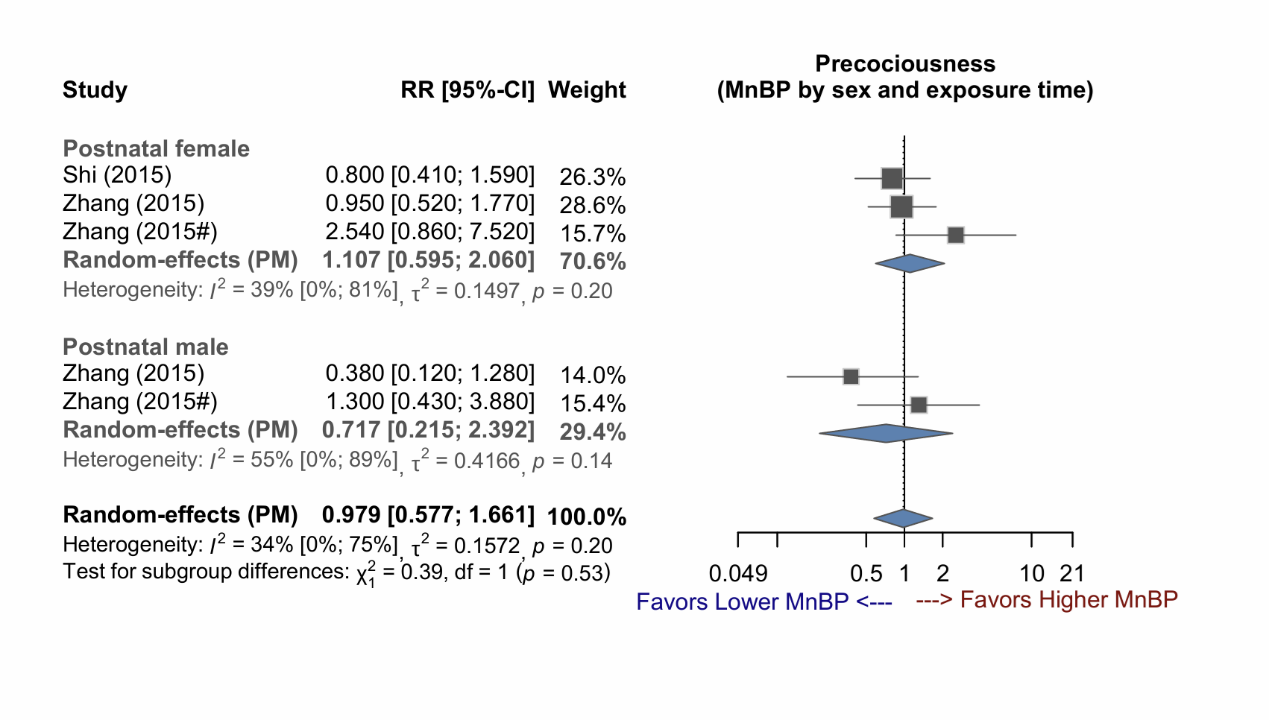

### **Supplementary Figure 3 (3-1~3-31) : Forest plots (unadjusted phthalate level for urine-SG)**

Abbreviations:

urine-SG, urine specific gravity; 2OH-MiBP, 2OH-mono-isobutyl phthalate; 3OH-MnBP, 3OH-mono-n-butyl phthalate; 5cx-MEPP, mono(2-ethyl-5-carboxypentyl)phthalate; 5OH-MEHP, mono(2-ethyl-5-hydroxyhexyl) phthalate; 5oxo-MEHP, mono(2-ethyl-5-oxohexyl)phthalate; cx-MiOP, monocarboxy isooctyl phthalate; MBzP, monobenzyl phthalate; MCPP, mono-(3-carboxypropyl) phthalate; MEHP, mono(2-ethylhexyl) phthalate; MEP, monoethyl phthalate; MiBP, mono-isobutyl phthalate; MMP, monomethyl phthalate; MnBP, mono-n-butyl phthalate

Supplementary Figure 3-1: Forest plot of studies demonstrating the RR of 2OH-MiBP exposure for early puberty.

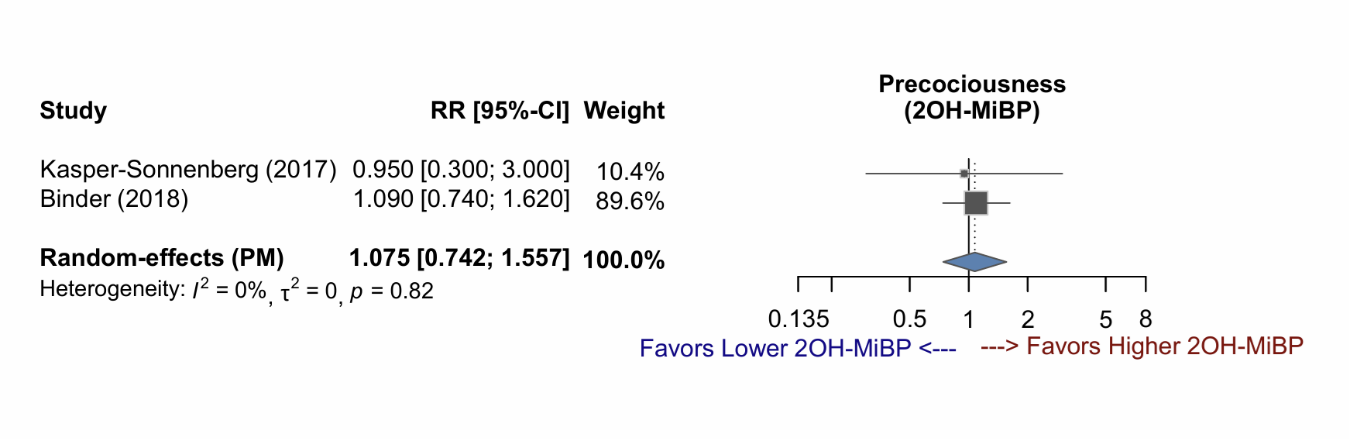

Supplementary Figure 3-2: Forest plot of studies demonstrating the RR of 3OH-MnBP exposure for early puberty.

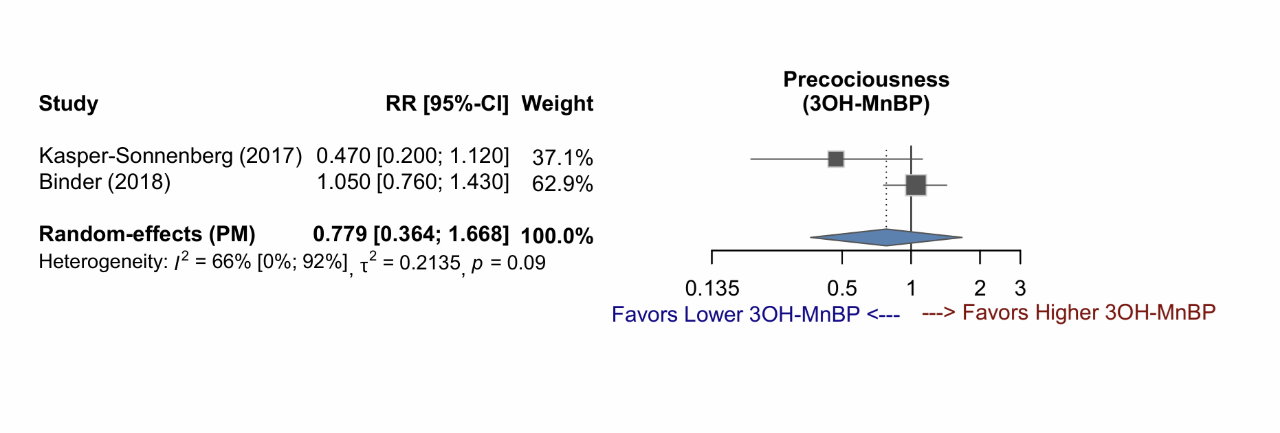

Supplementary Figure 3-3: Forest plot of studies demonstrating the RR of 5cx-MEPP exposure for early puberty, stratified by sex.

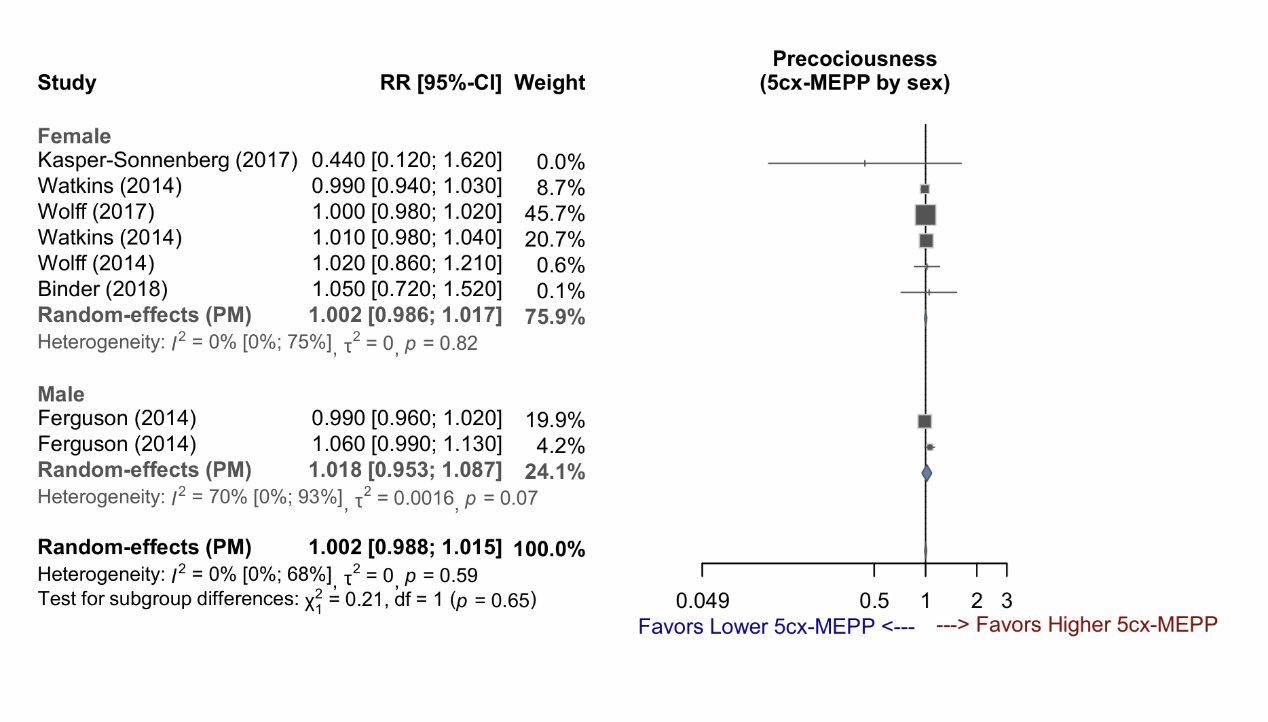

Supplementary Figure 3-4: Forest plot of studies demonstrating the RR of 5cx-MEPP exposure for early puberty, stratified by exposure time.

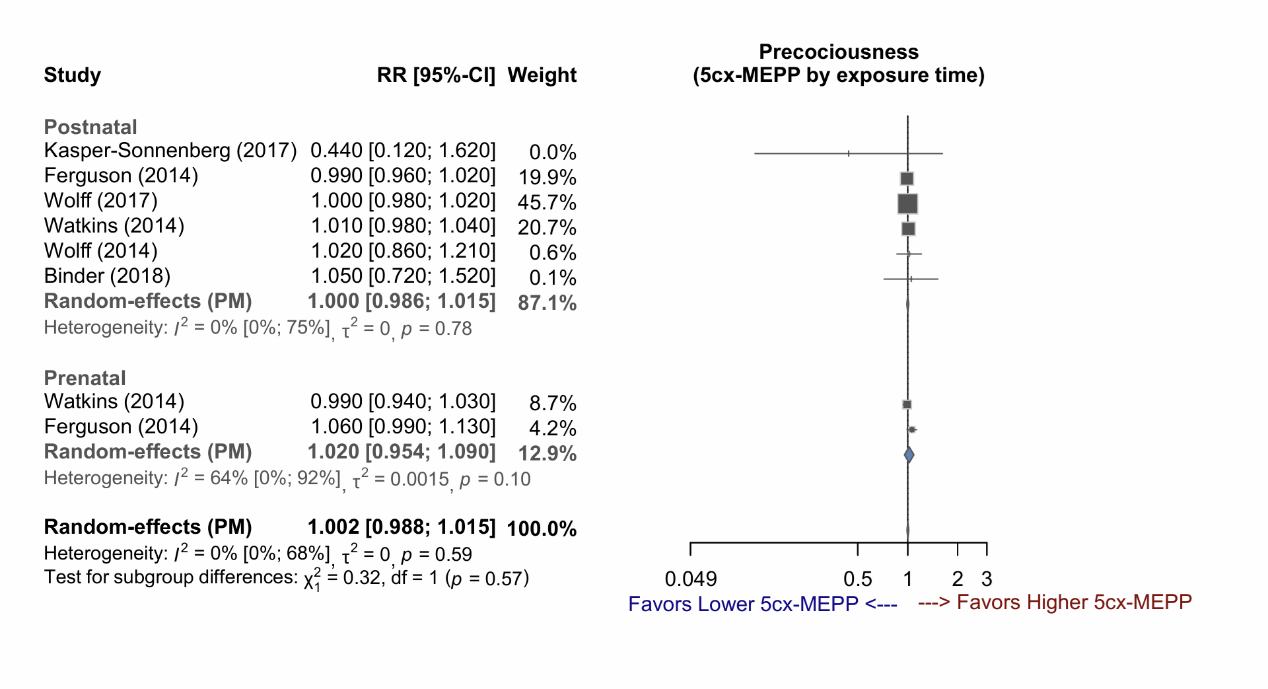

Supplementary Figure 3-5: Forest plot of studies demonstrating the RR of 5cx-MEPP exposure for early puberty, stratified by sex and exposure time.

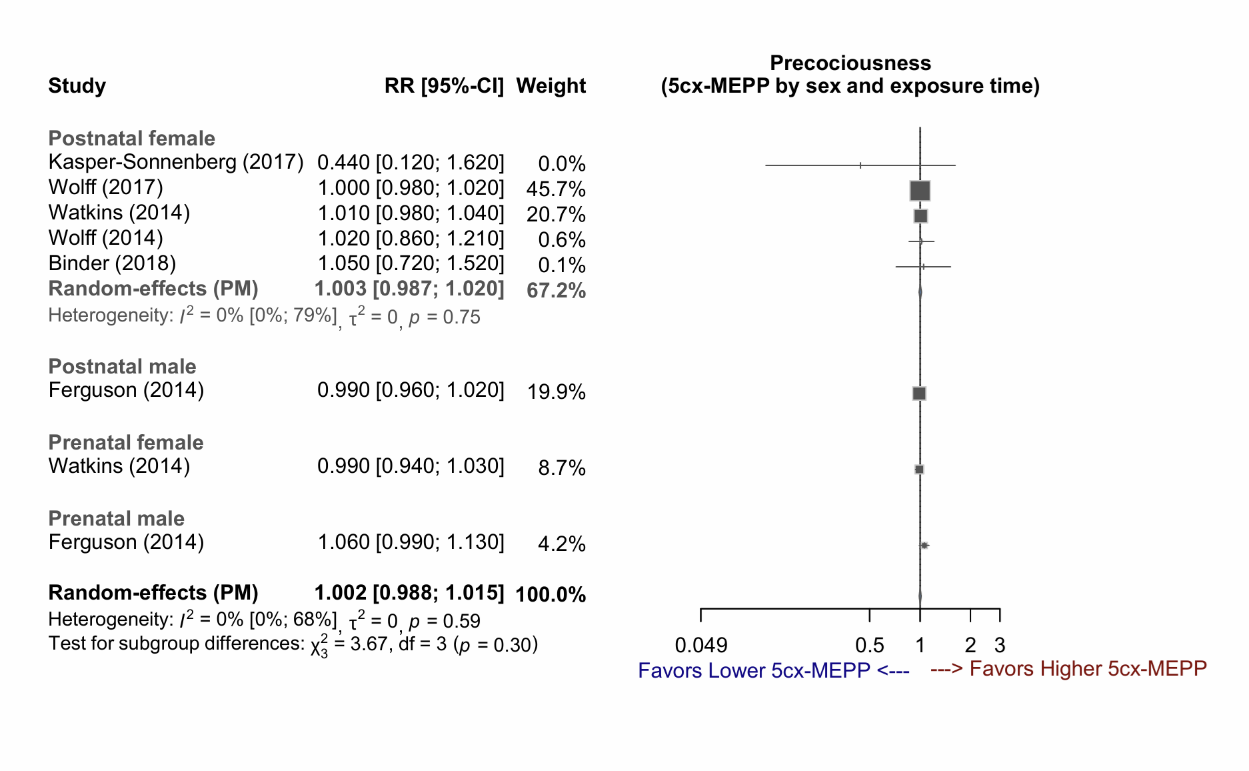

Supplementary Figure 3-6: Forest plot of studies demonstrating the RR of 5OH-MEHP exposure for early puberty, stratified by sex.

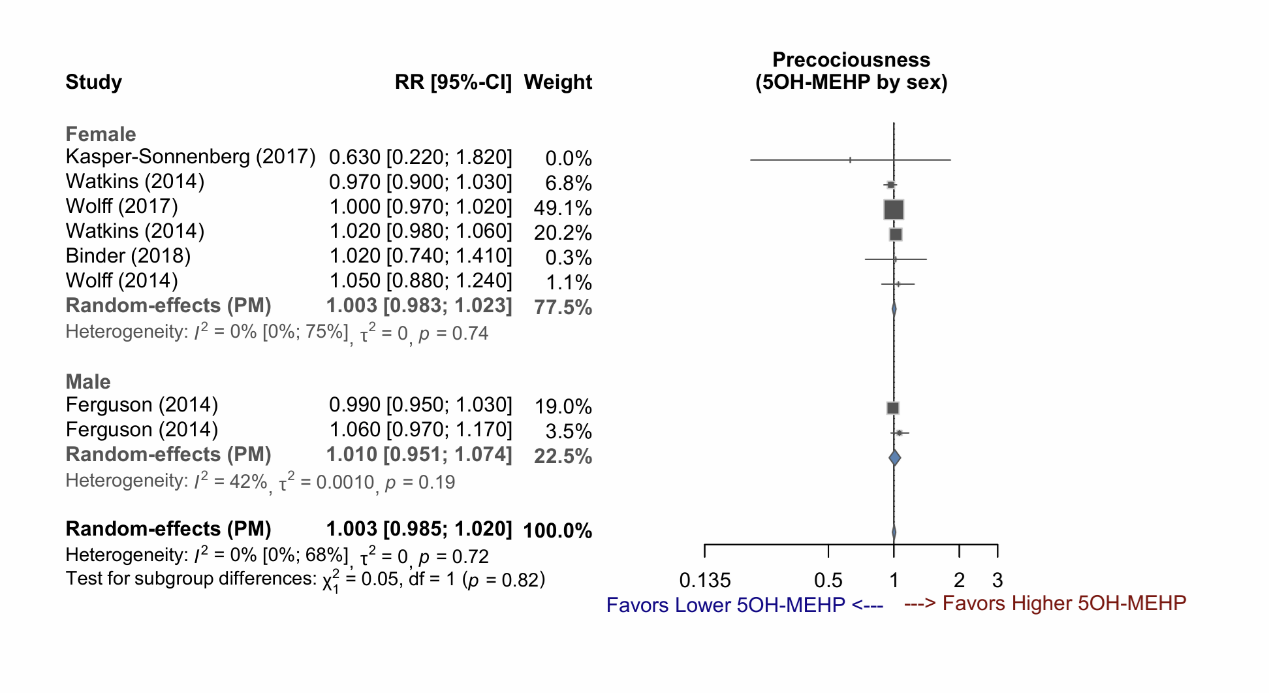

Supplementary Figure 3-7: Forest plot of studies demonstrating the RR of 5OH-MEHP exposure for early puberty, stratified by exposure time.

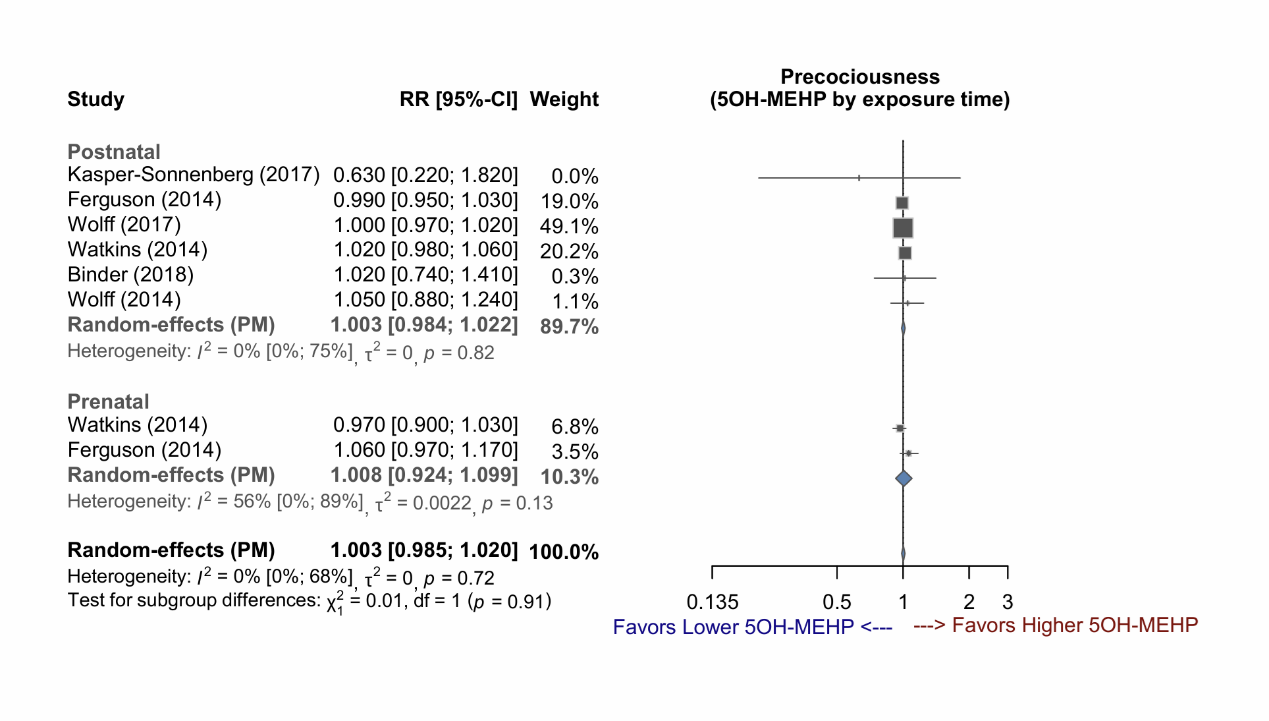

Supplementary Figure 3-8: Forest plot of studies demonstrating the RR of 5OH-MEHP exposure for early puberty, stratified by sex and exposure time.

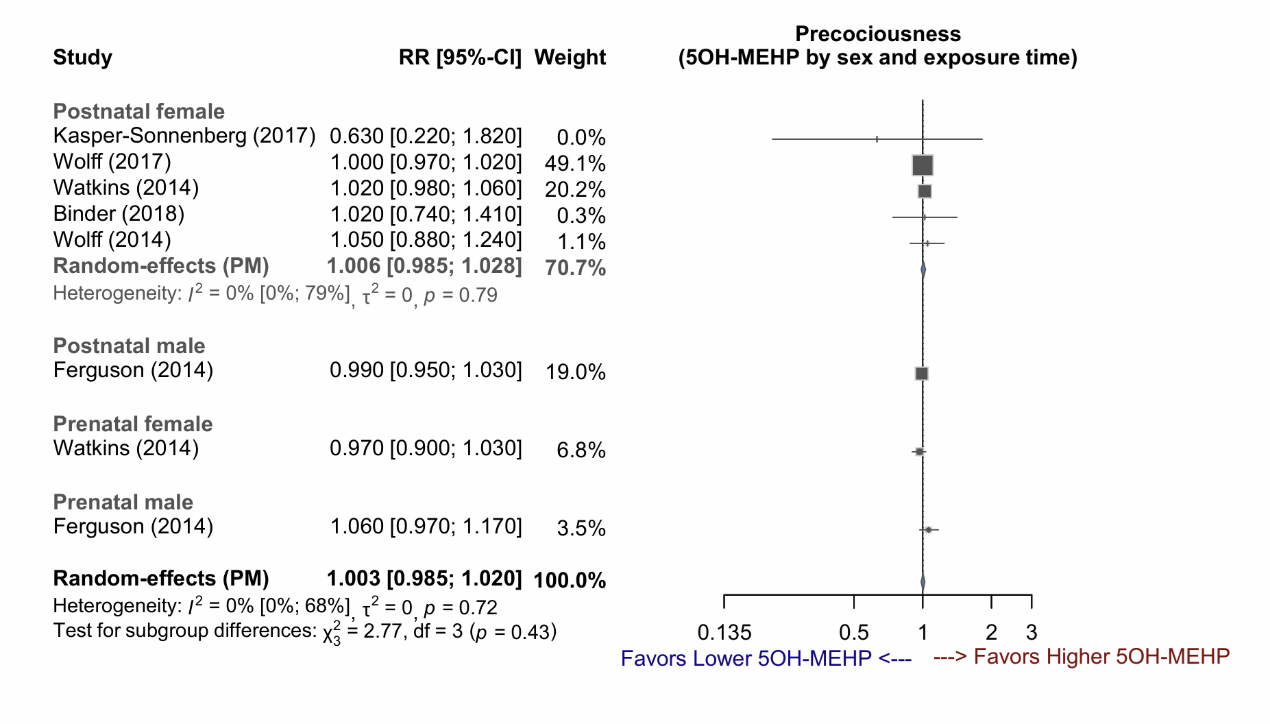

Supplementary Figure 3-9: Forest plot of studies demonstrating the RR of 5oxo-MEHP exposure for early puberty, stratified by sex.

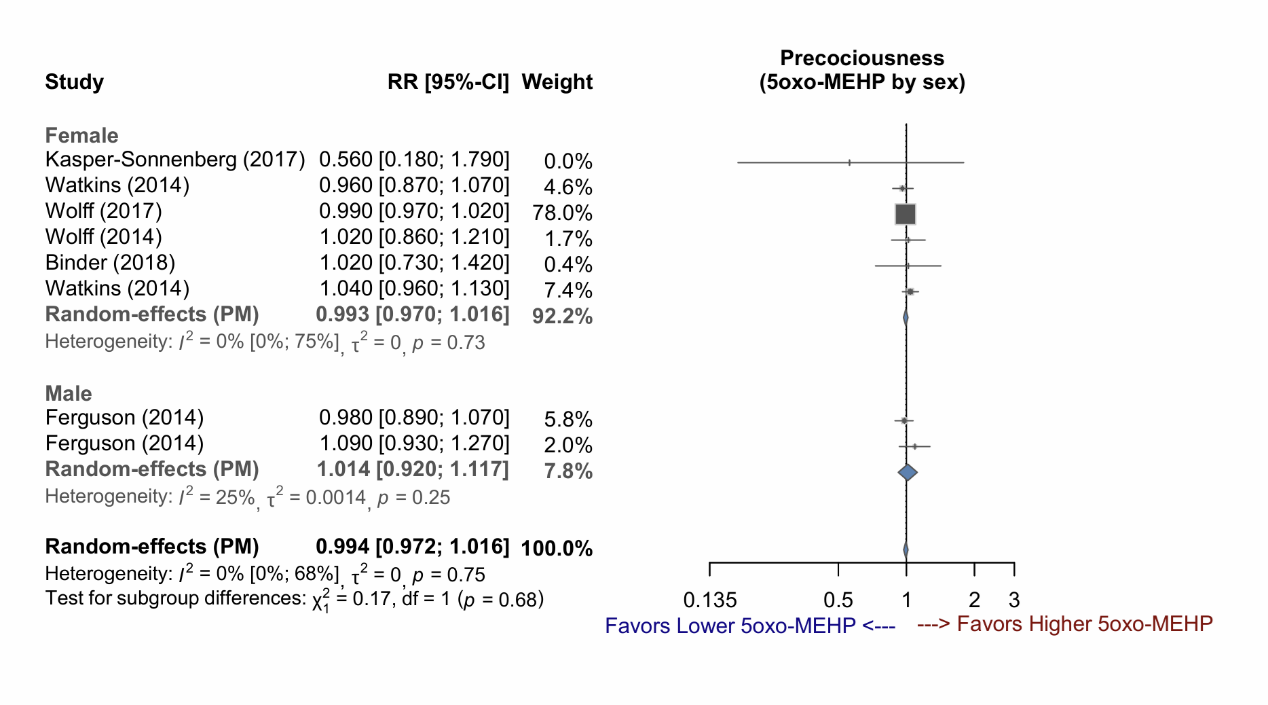

Supplementary Figure 3-10: Forest plot of studies demonstrating the RR of 5oxo-MEHP exposure for early puberty, stratified by exposure time.

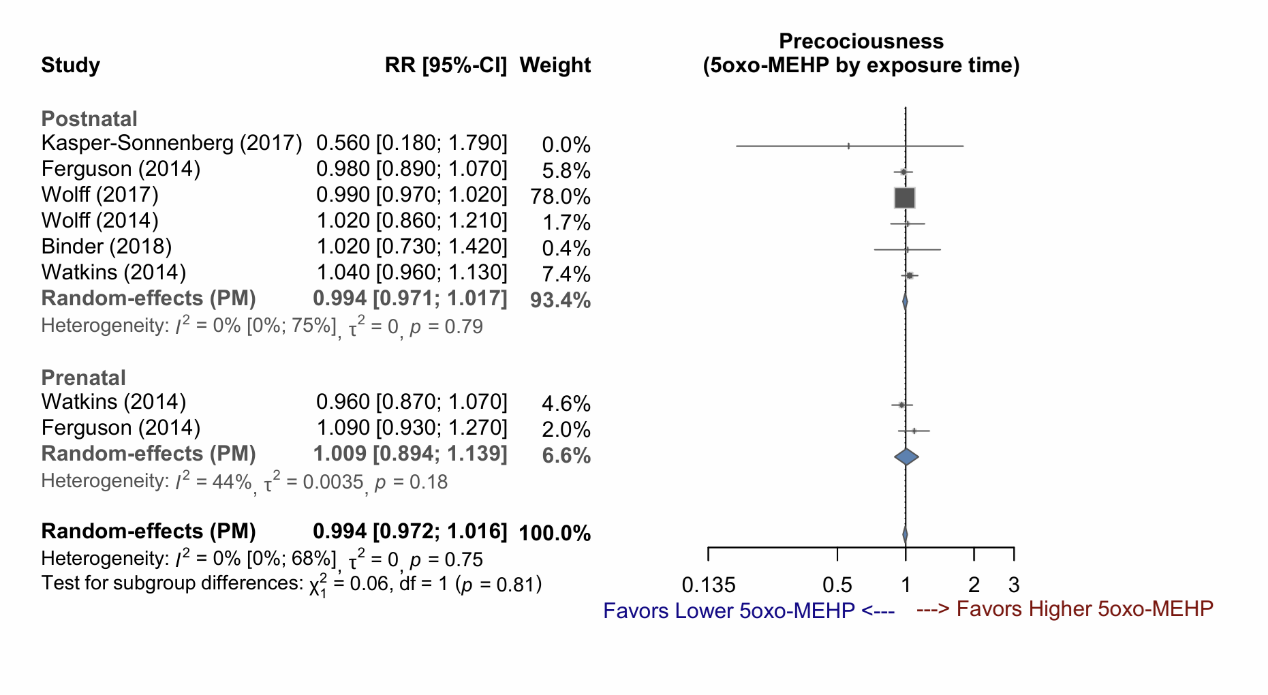

Supplementary Figure 3-11: Forest plot of studies demonstrating the RR of 5oxo-MEHP exposure for early puberty, stratified by sex and exposure time.

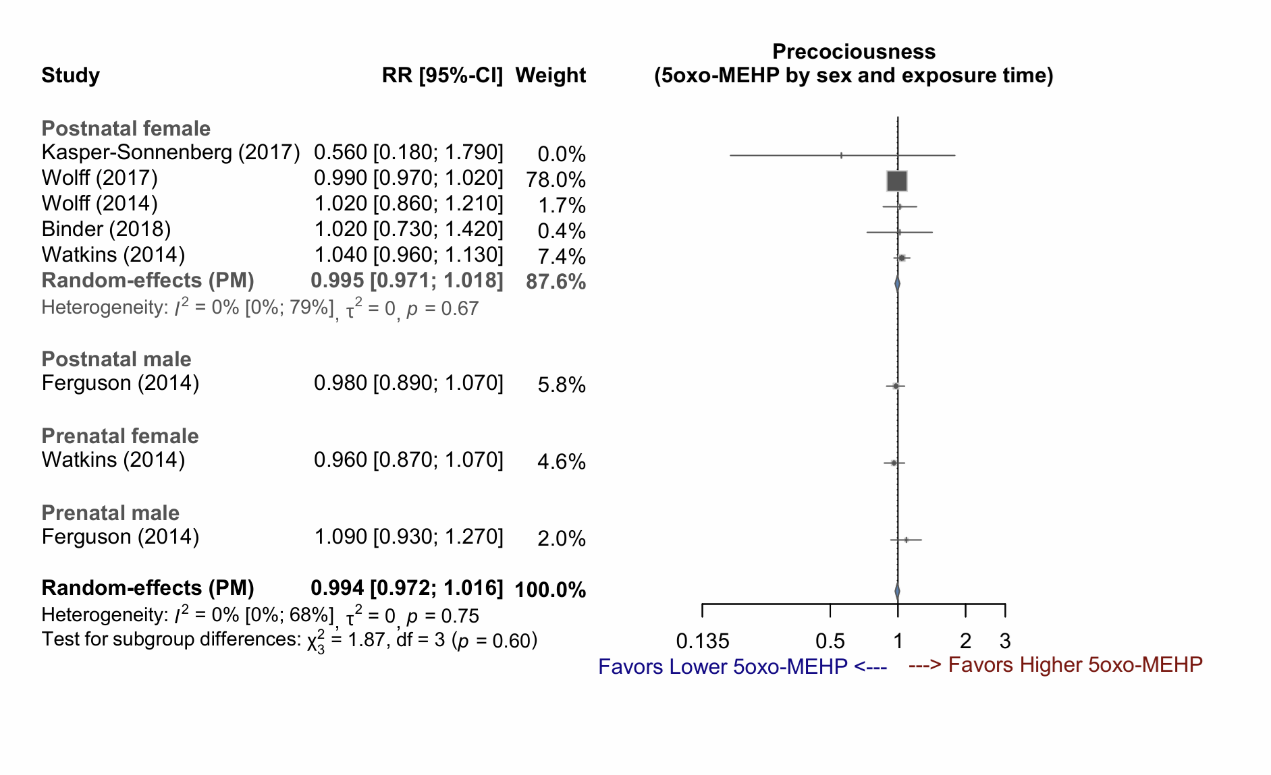

Supplementary Figure 3-12: Forest plot of studies demonstrating the RR of cx-MiOP exposure for early puberty.

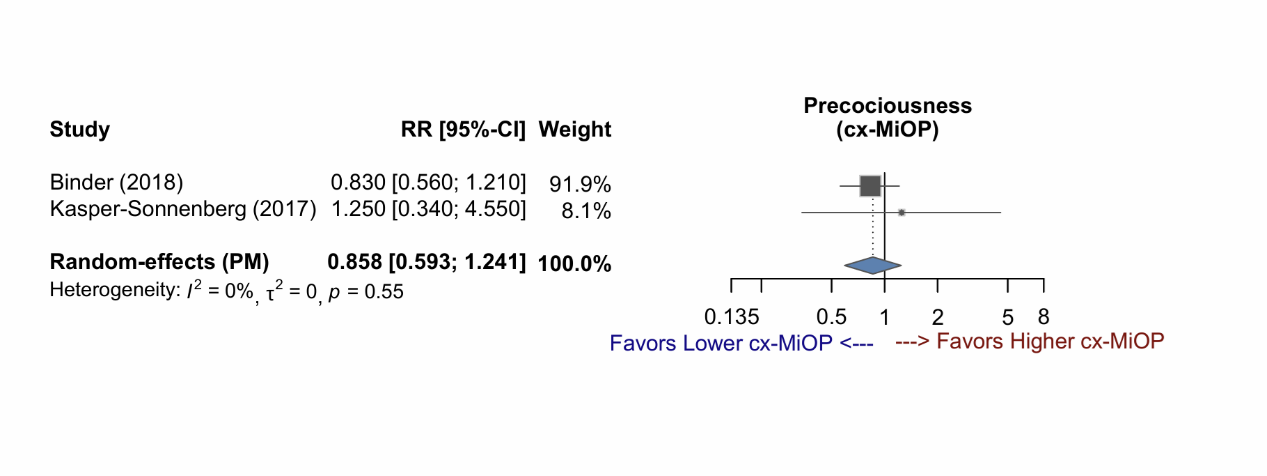

Supplementary Figure 3-13: Forest plot of studies demonstrating the RR of MBzP exposure for early puberty, stratified by sex.

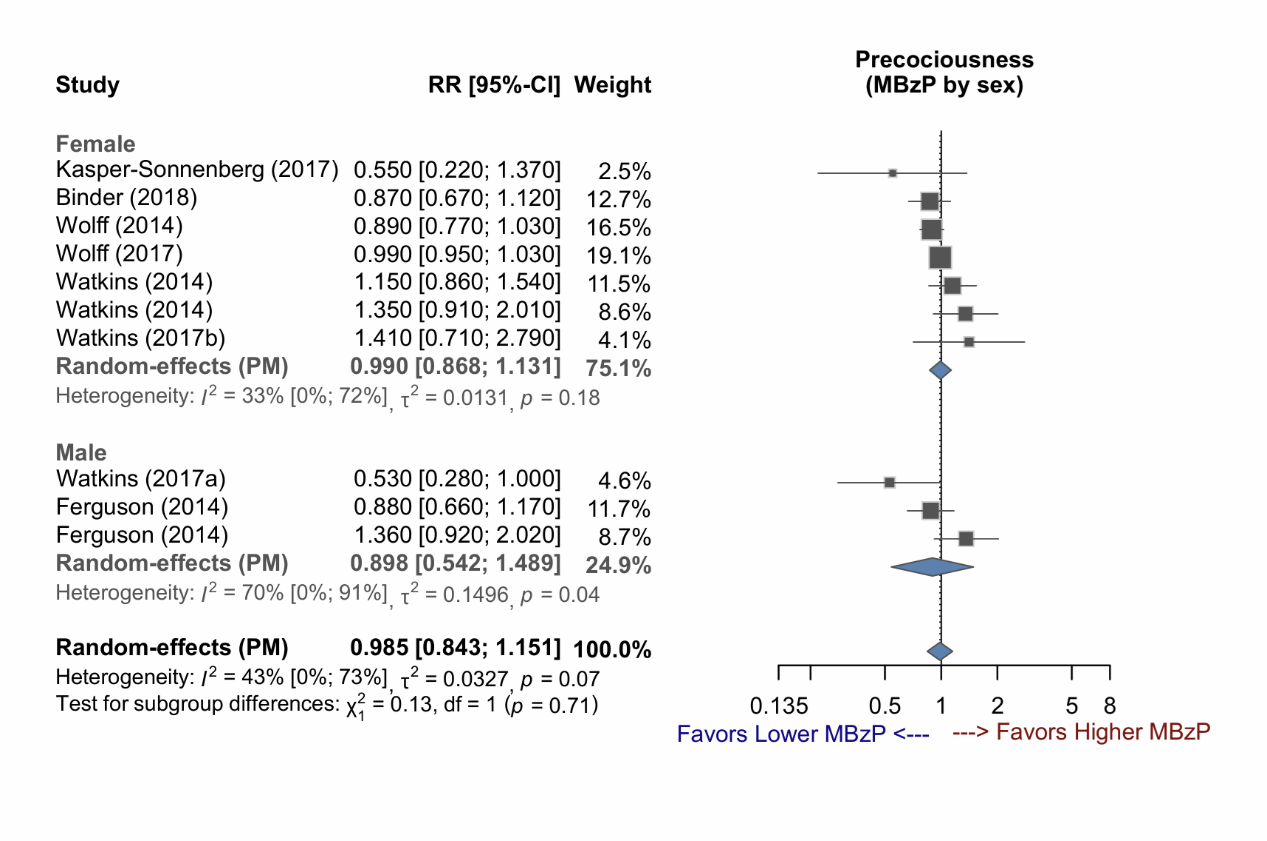

Supplementary Figure 3-14: Forest plot of studies demonstrating the RR of MBzP exposure for early puberty, stratified by exposure time.

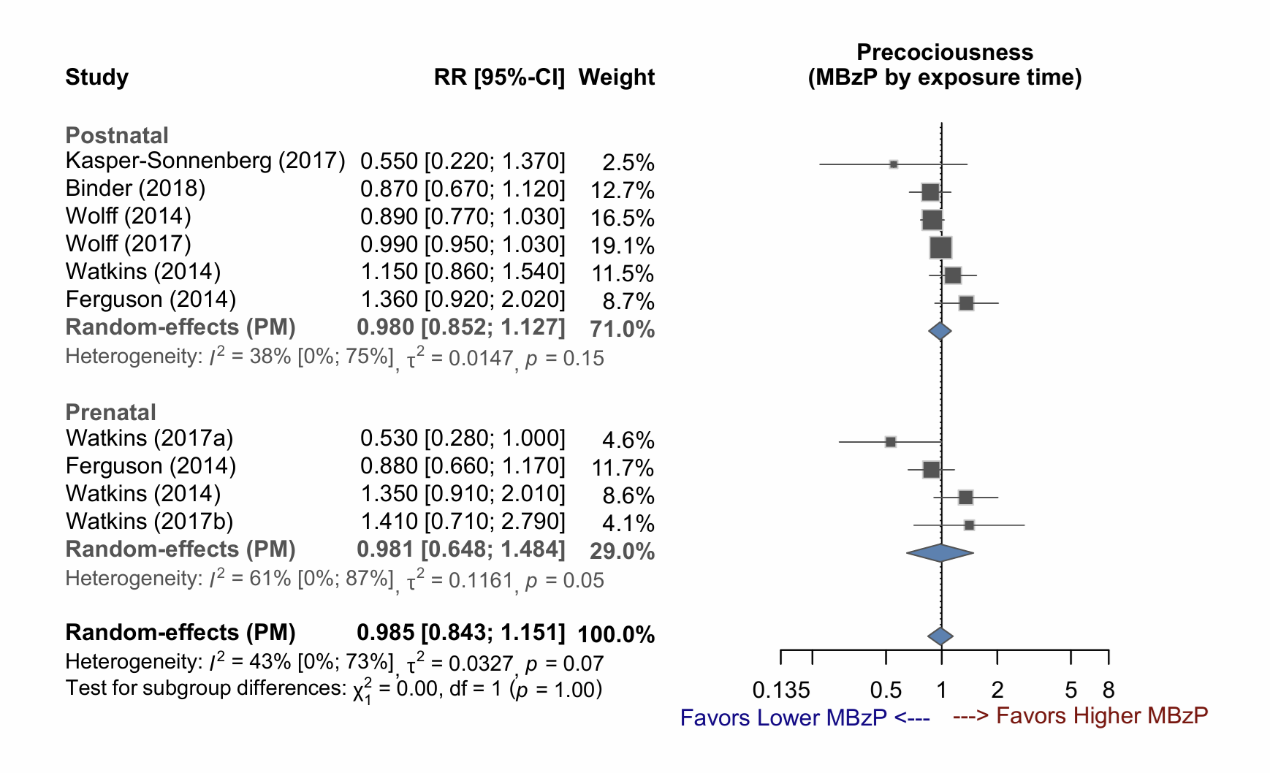

Supplementary Figure 3-15: Forest plot of studies demonstrating the RR of MBzP exposure for early puberty, stratified by sex and exposure time.

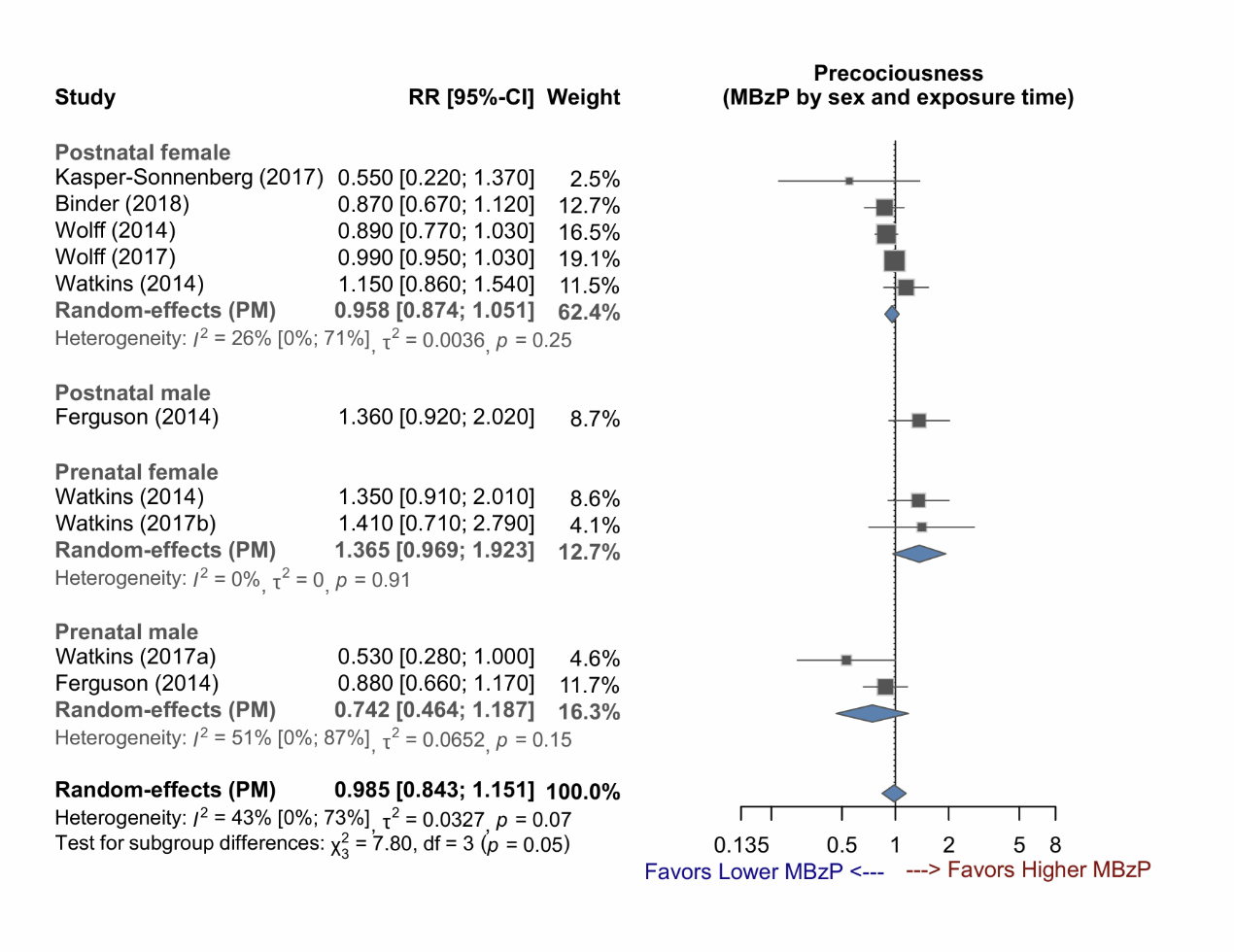

Supplementary Figure 3-16: Forest plot of studies demonstrating the RR of MCPP exposure for early puberty, stratified by sex.

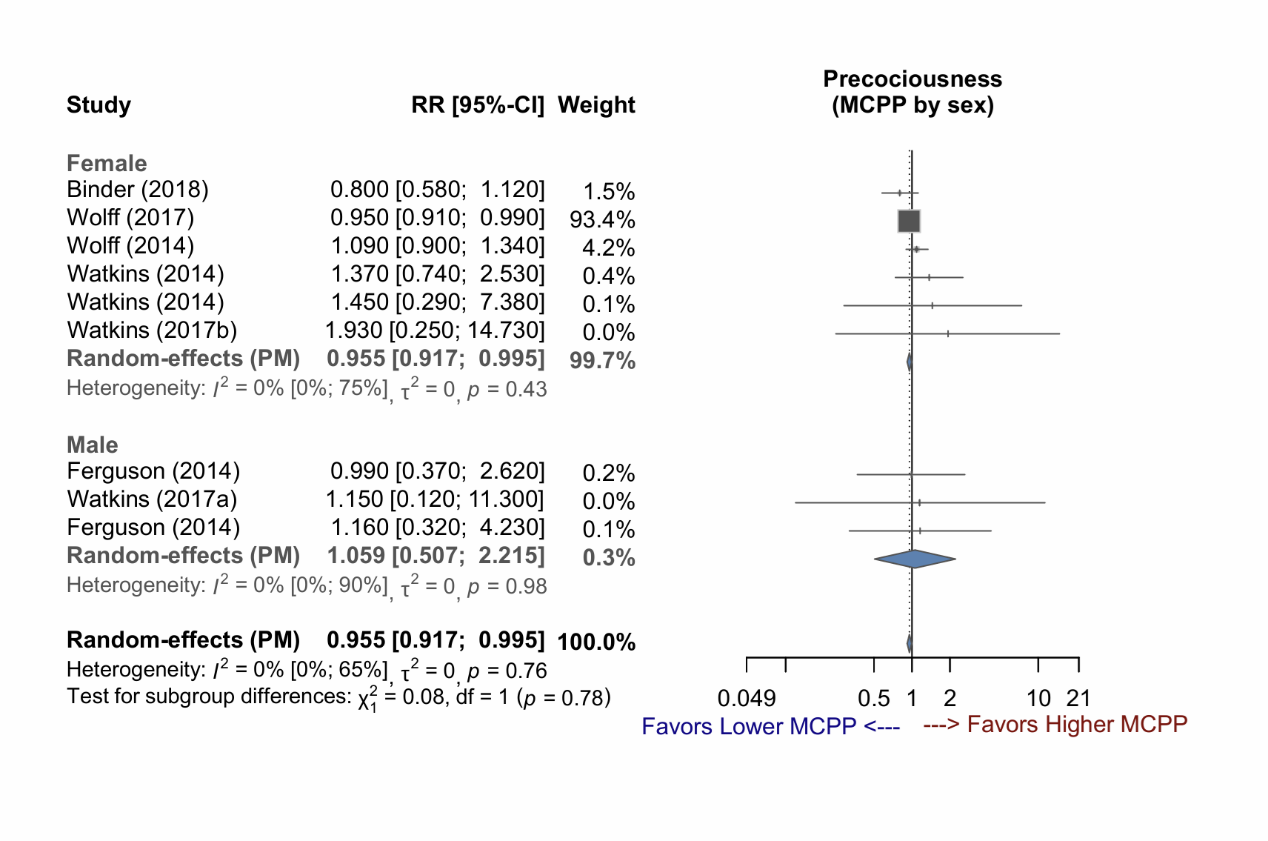

Supplementary Figure 3-17: Forest plot of studies demonstrating the RR of MCPP exposure for early puberty, stratified by exposure time.

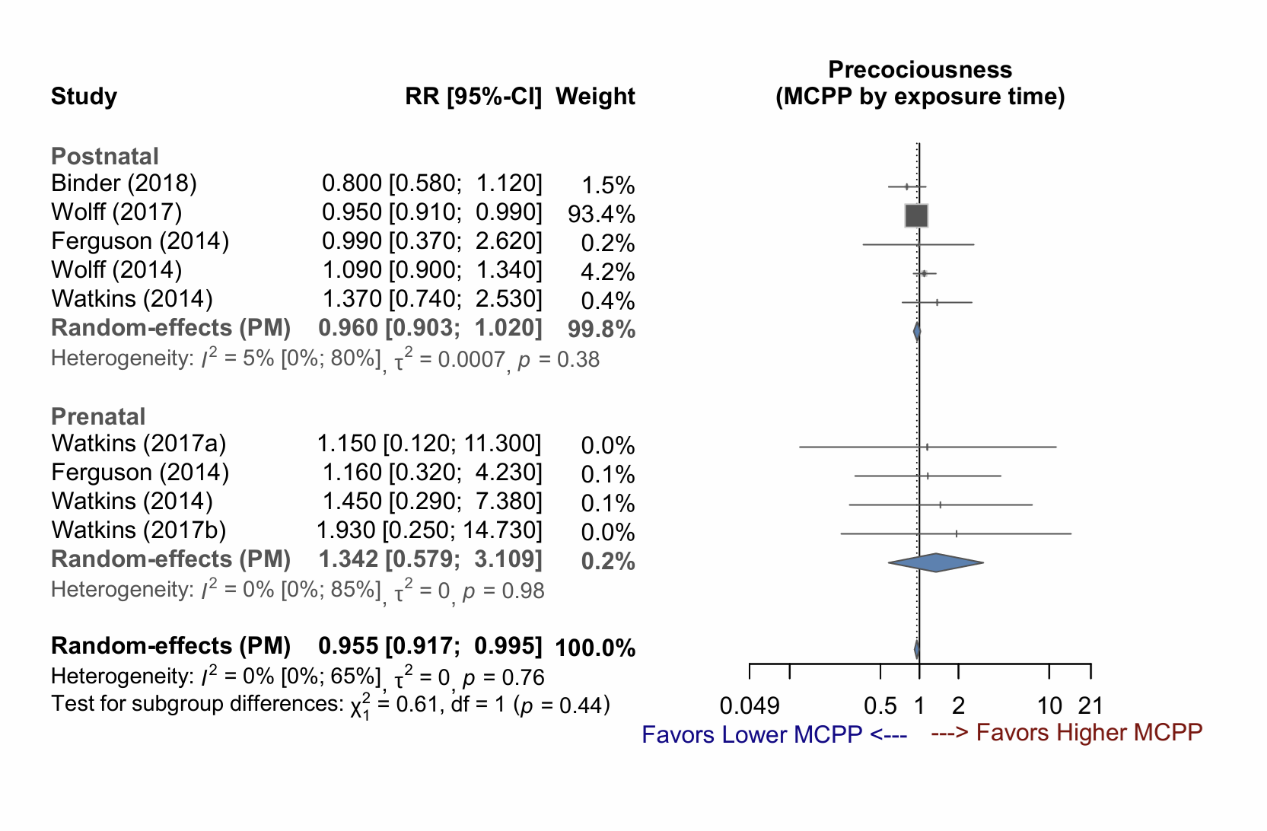

Supplementary Figure 3-18: Forest plot of studies demonstrating the RR of MCPP exposure for early puberty, stratified by sex and exposure time.

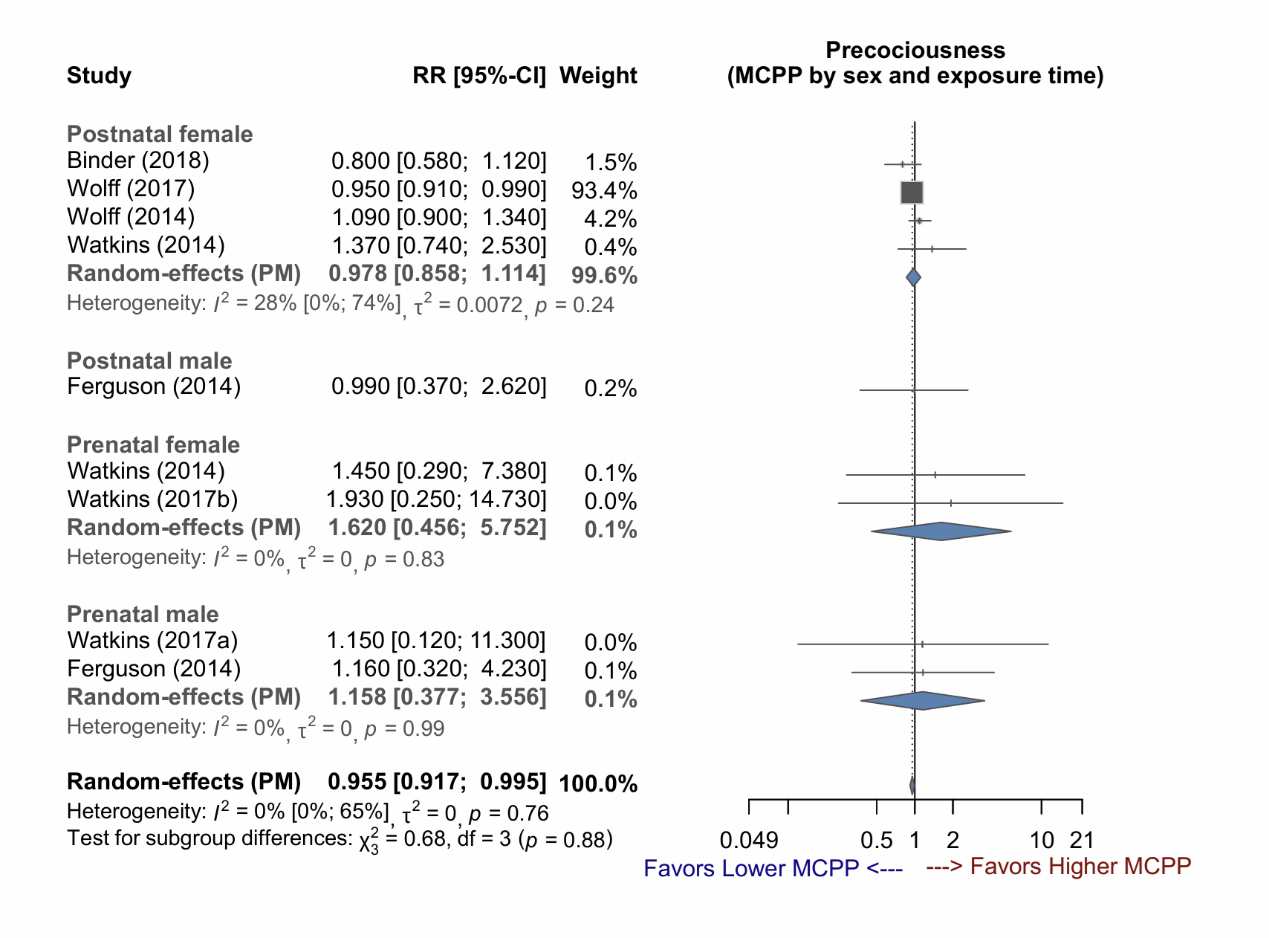

Supplementary Figure 3-19: Forest plot of studies demonstrating the RR of MEHP exposure for early puberty, stratified by sex.

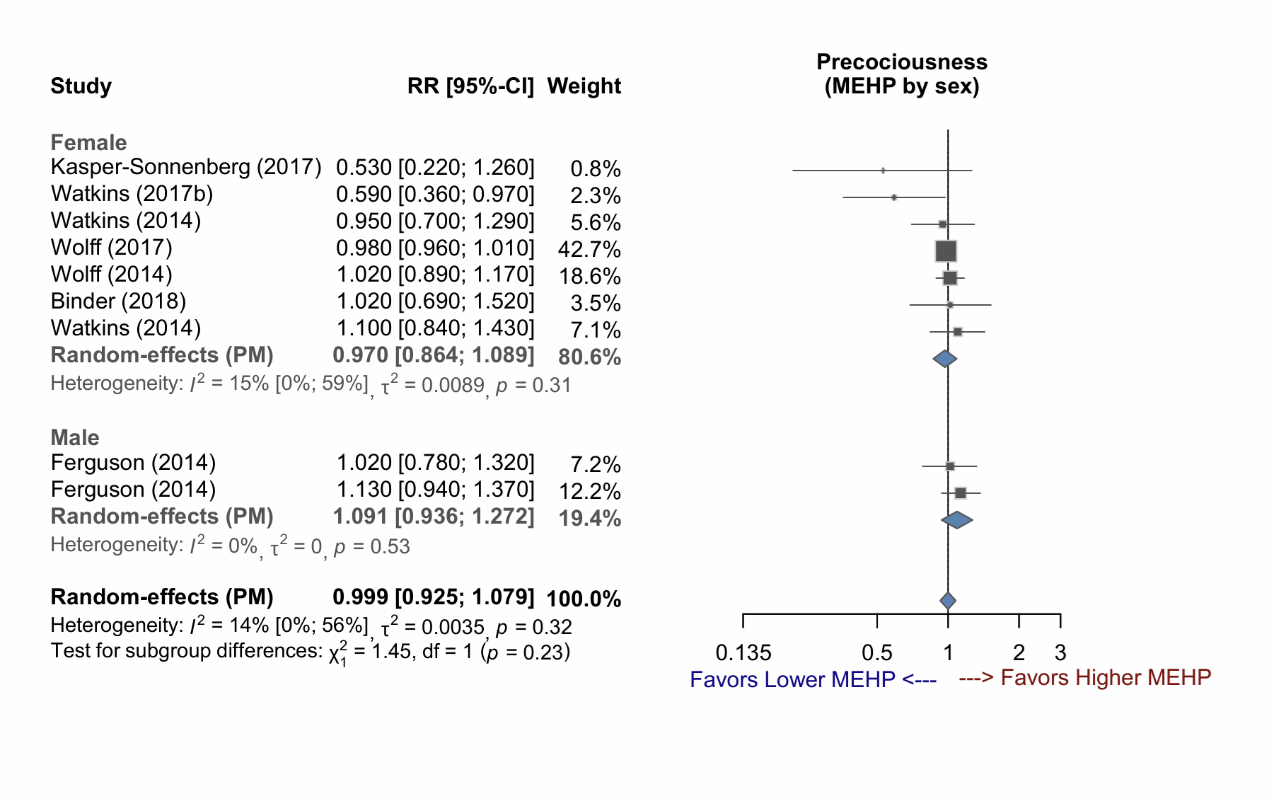

Supplementary Figure 3-20: Forest plot of studies demonstrating the RR of MEHP exposure for early puberty, stratified by exposure time.

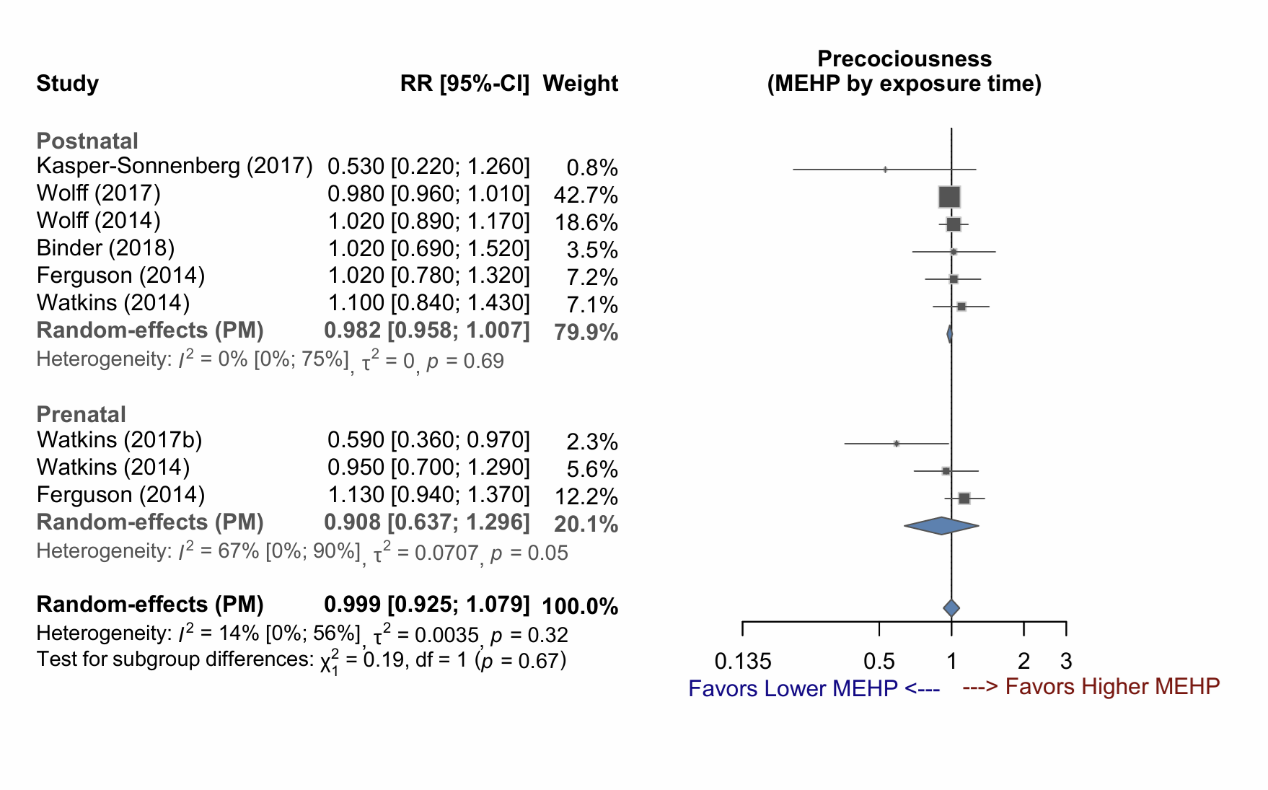

Supplementary Figure 3-21: Forest plot of studies demonstrating the RR of MEHP exposure for early puberty, stratified by sex and exposure time.

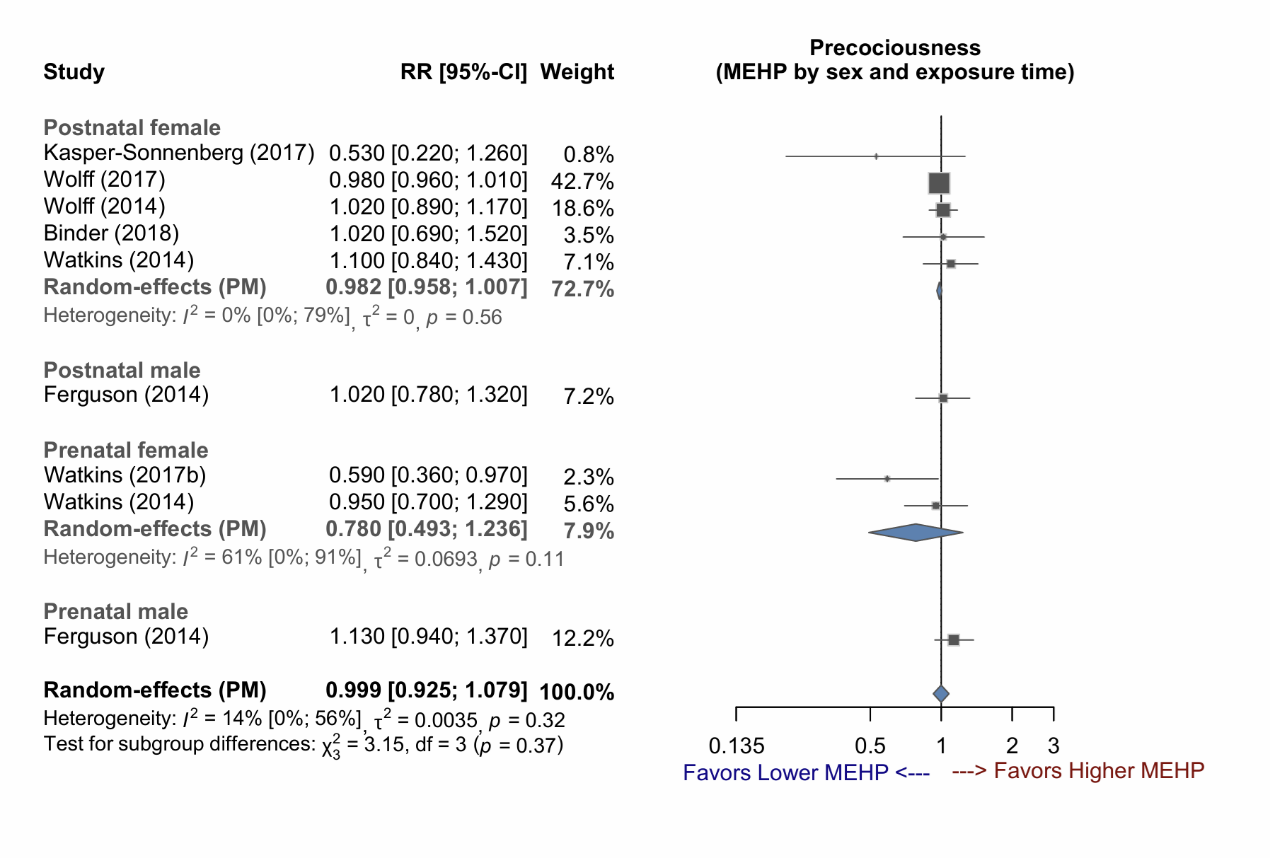

Supplementary Figure 3-22: Forest plot of studies demonstrating the RR of MEP exposure for early puberty, stratified by sex.

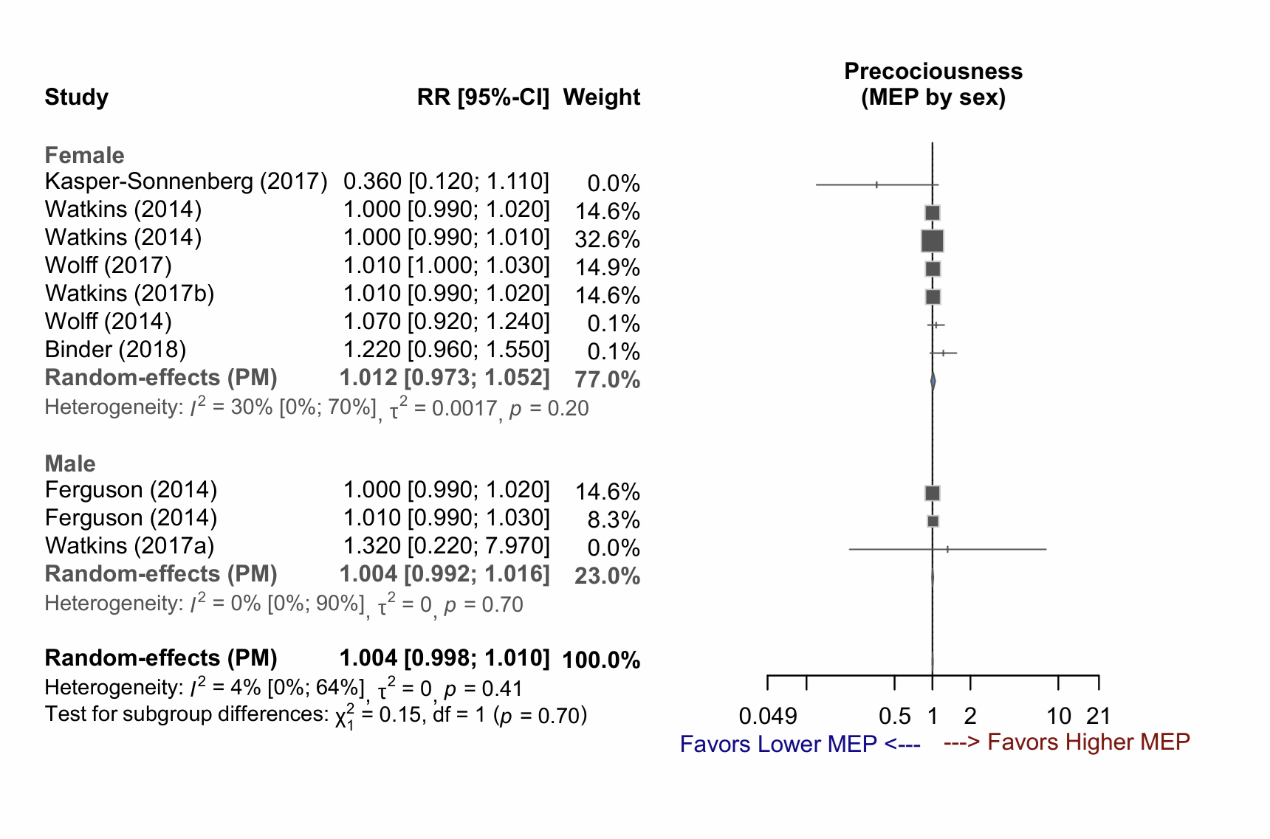

Supplementary Figure 3-23: Forest plot of studies demonstrating the RR of MEP exposure for early puberty, stratified by exposure time.

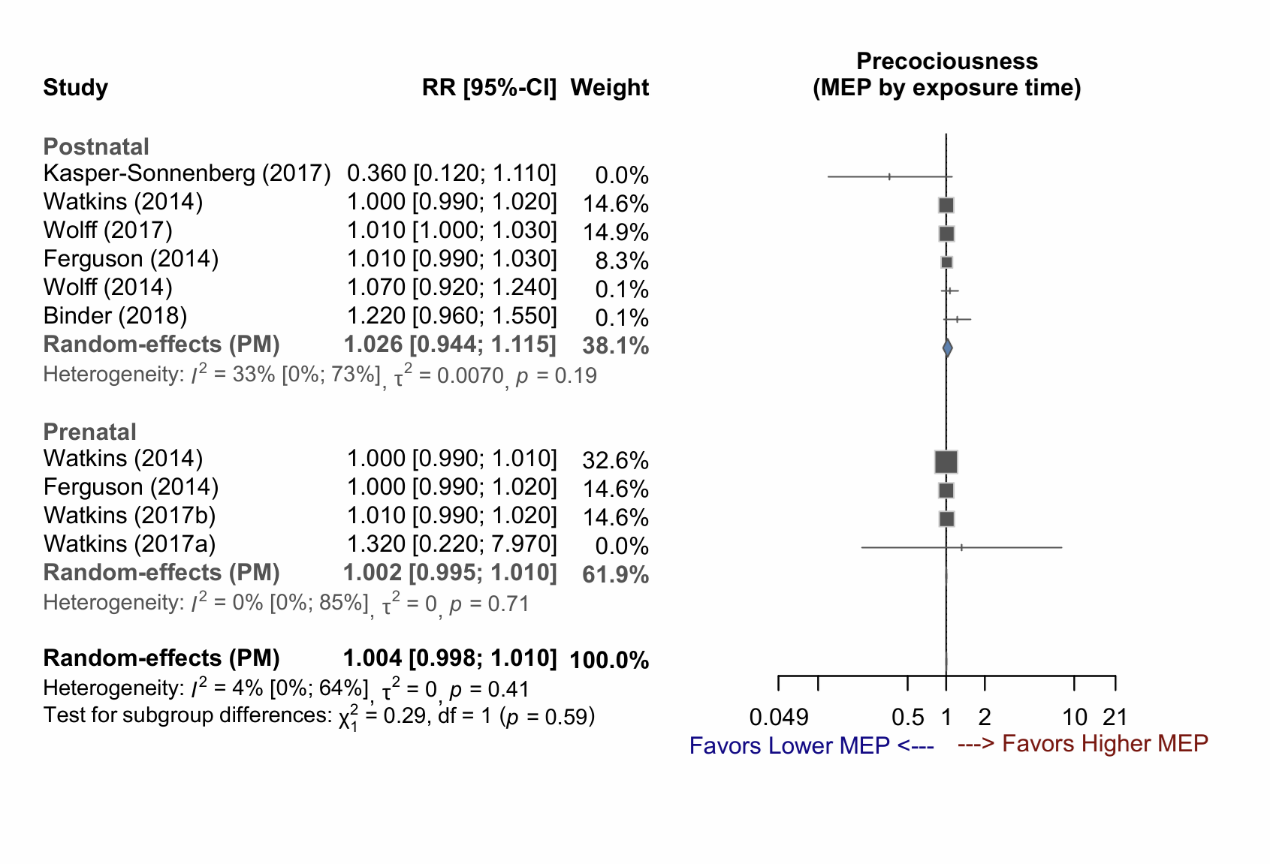

Supplementary Figure 3-24: Forest plot of studies demonstrating the RR of MEP exposure for early puberty, stratified by sex and exposure time.

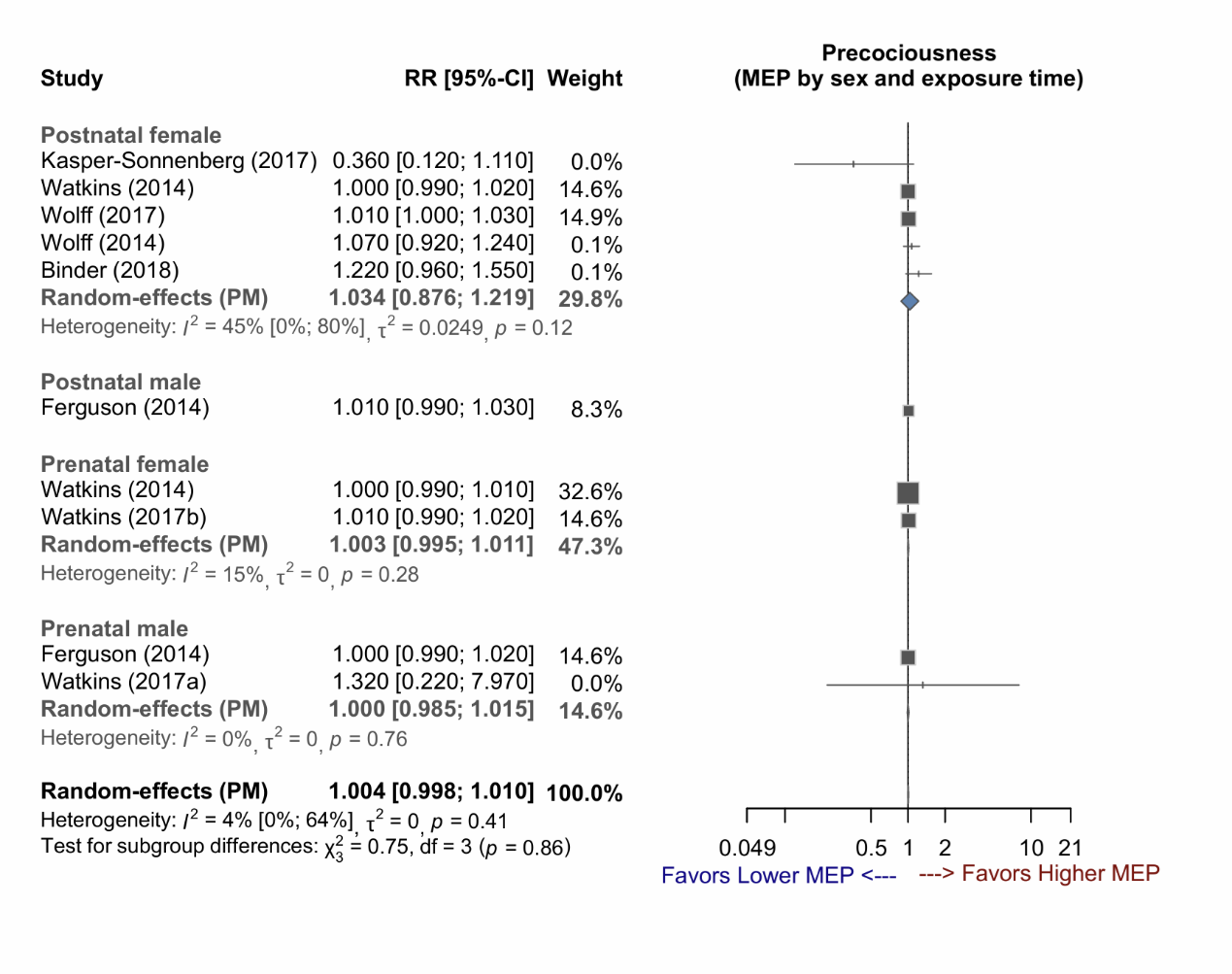

Supplementary Figure 3-25: Forest plot of studies demonstrating the RR of MiBP exposure for early puberty, stratified by sex.

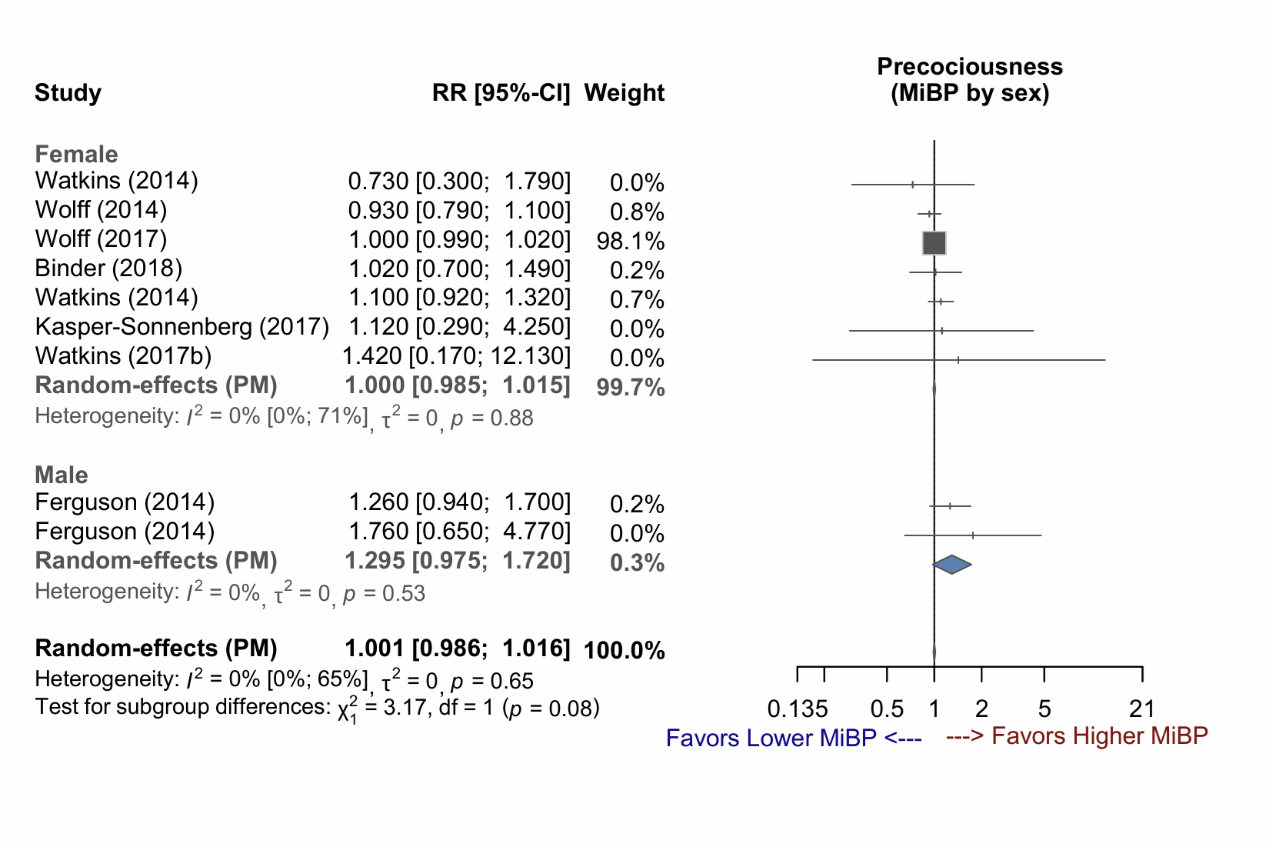

Supplementary Figure 3-26: Forest plot of studies demonstrating the RR of MiBP exposure for early puberty, stratified by exposure time.

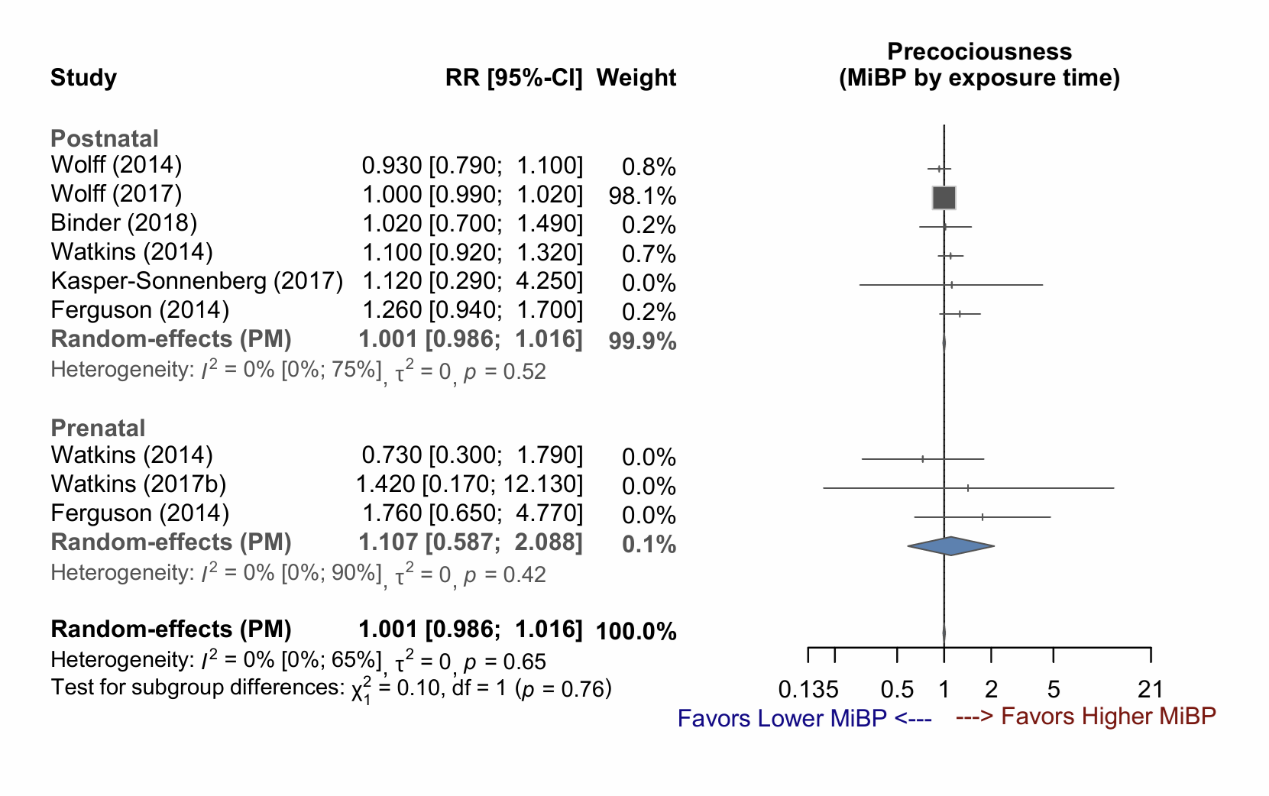

Supplementary Figure 3-27: Forest plot of studies demonstrating the RR of MiBP exposure for early puberty, stratified by sex and exposure time.

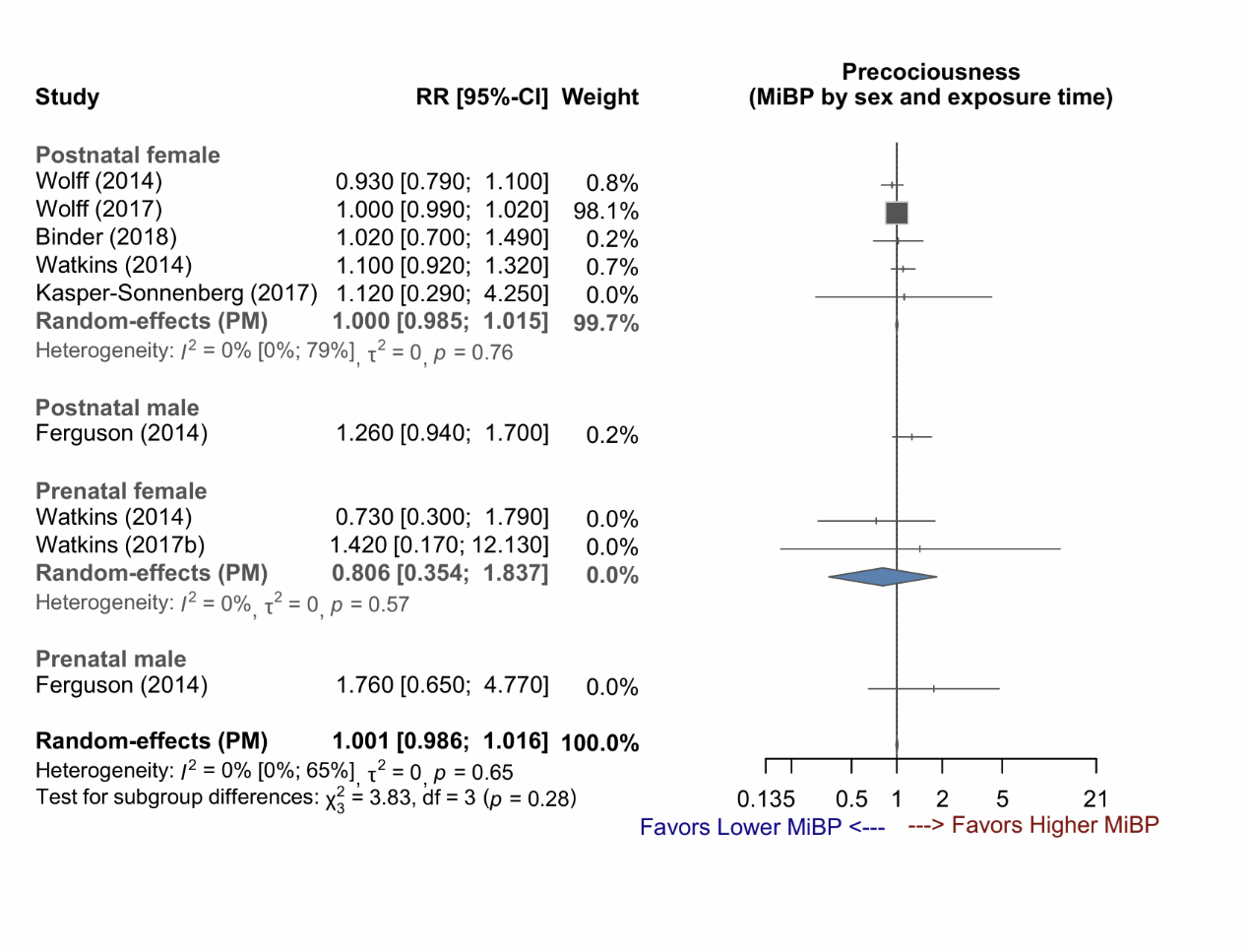

Supplementary Figure 3-28: Forest plot of studies demonstrating the RR of MMP exposure for early puberty.

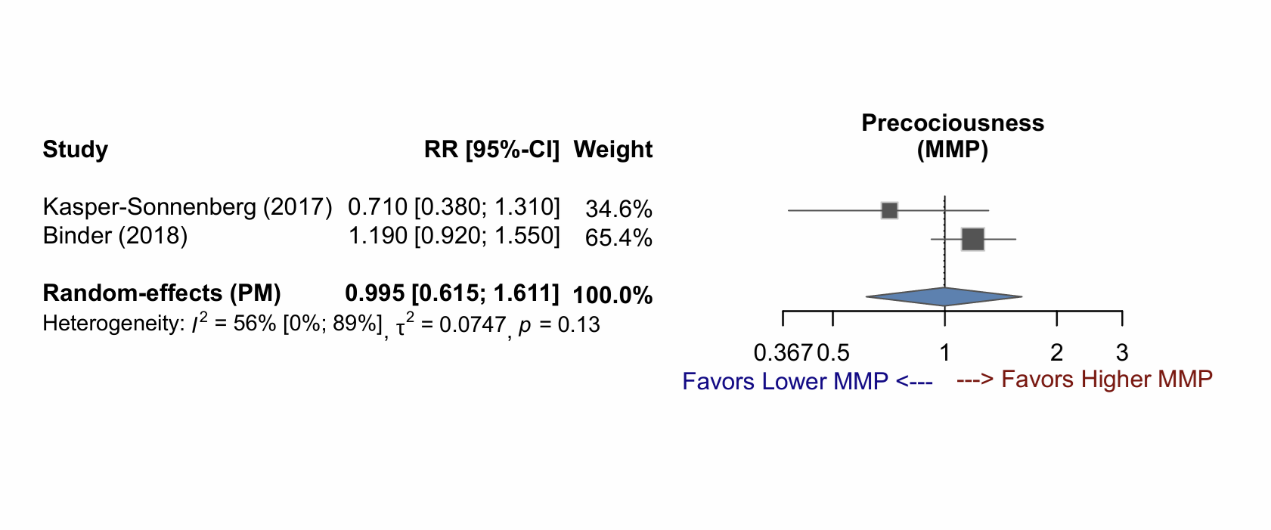

Supplementary Figure 3-29: Forest plot of studies demonstrating the RR of MnBP exposure for early puberty, stratified by sex.

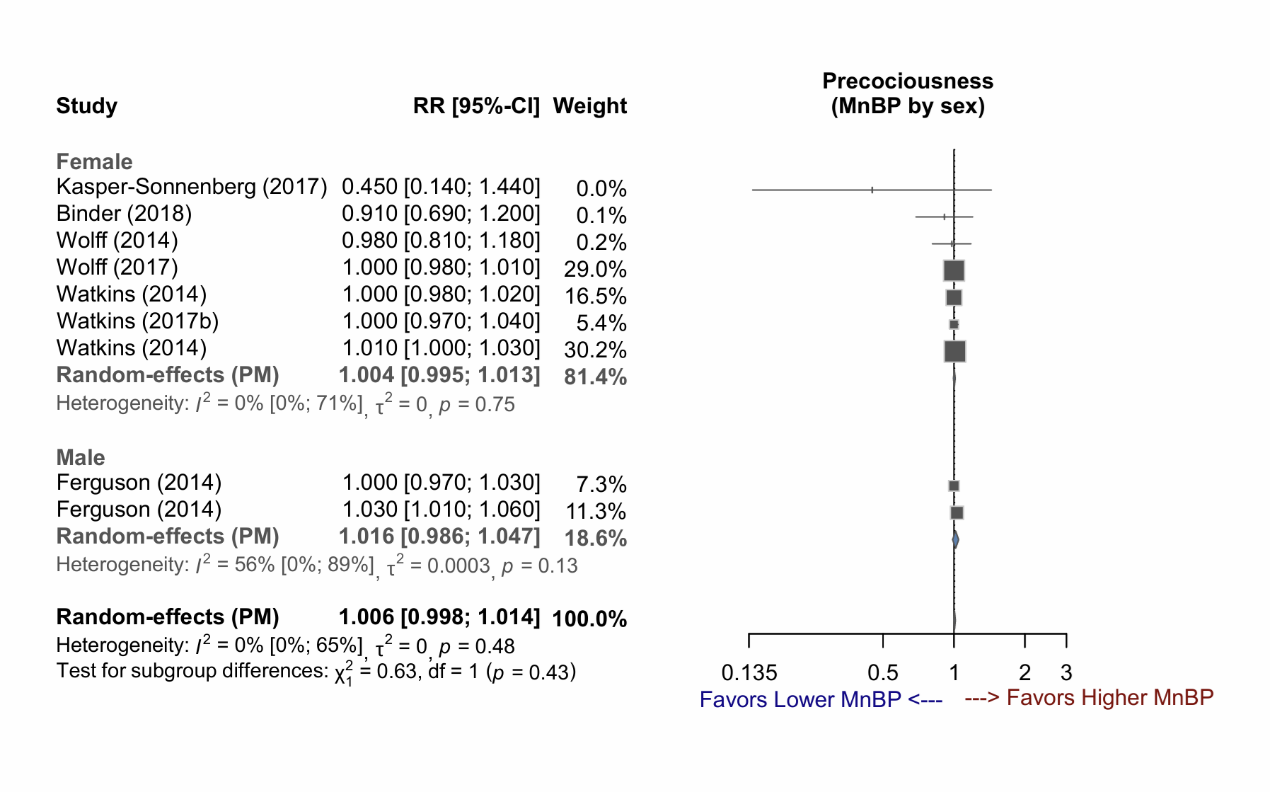

Supplementary Figure 3-30: Forest plot of studies demonstrating the RR of MnBP exposure for early puberty, stratified by exposure time.

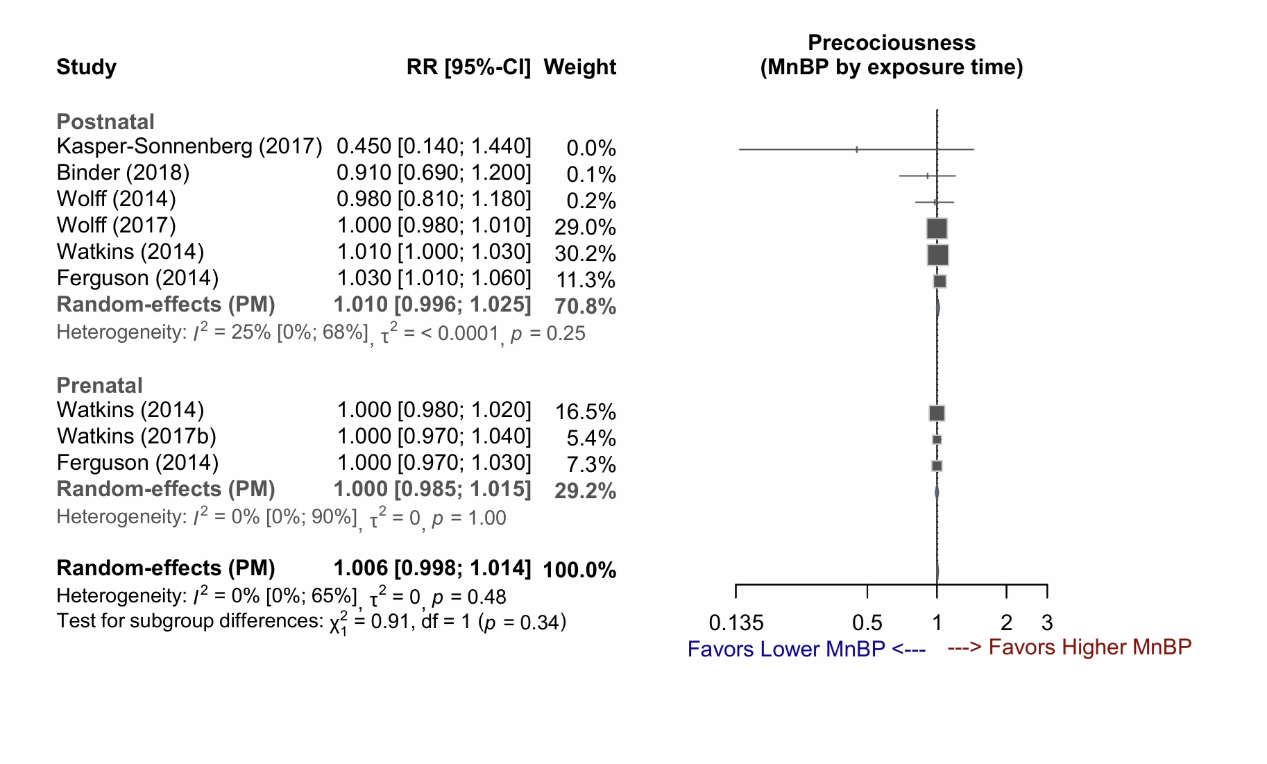

Supplementary Figure 3-31: Forest plot of studies demonstrating the RR of MnBP exposure for early puberty, stratified by sex and exposure time.

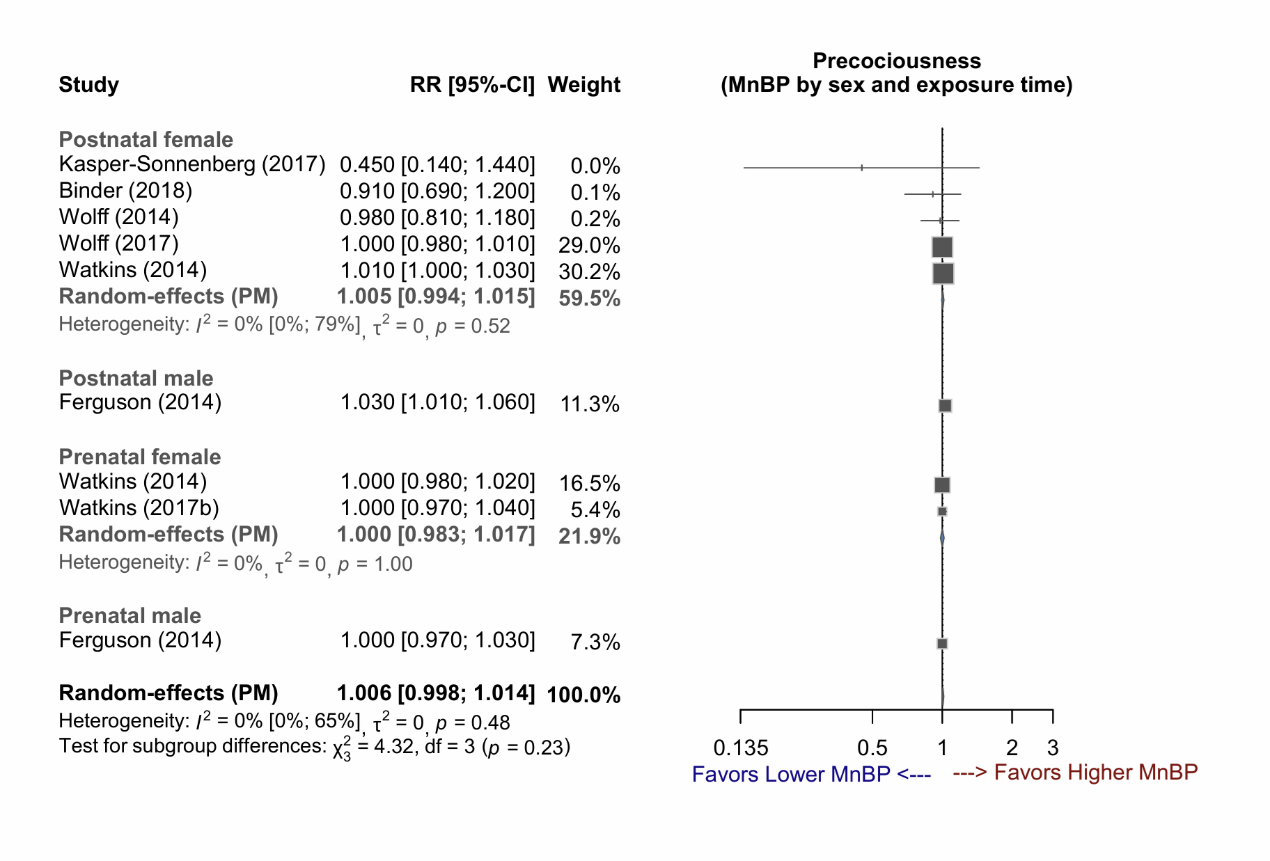

### **Supplementary Figure 4 (4-1~4-8) : Funnel plots of studies**

Abbreviations:

urine-SG, urine specific gravity; 5OH-MEHP, mono(2-ethyl-5-hydroxyhexyl) phthalate; 5oxo-MEHP, mono(2-ethyl-5-oxohexyl)phthalate; cx-MiOP, monocarboxy isooctyl phthalate; MBzP, monobenzyl phthalate; MEHP, mono(2-ethylhexyl) phthalate; MEP, monoethyl phthalate; MnBP, mono-n-butyl phthalate

Supplementary Figure 4-1: Funnel plot of studies demonstrating the publication bias of 5OH-MEHP exposure (combined) on the risk of early puberty.

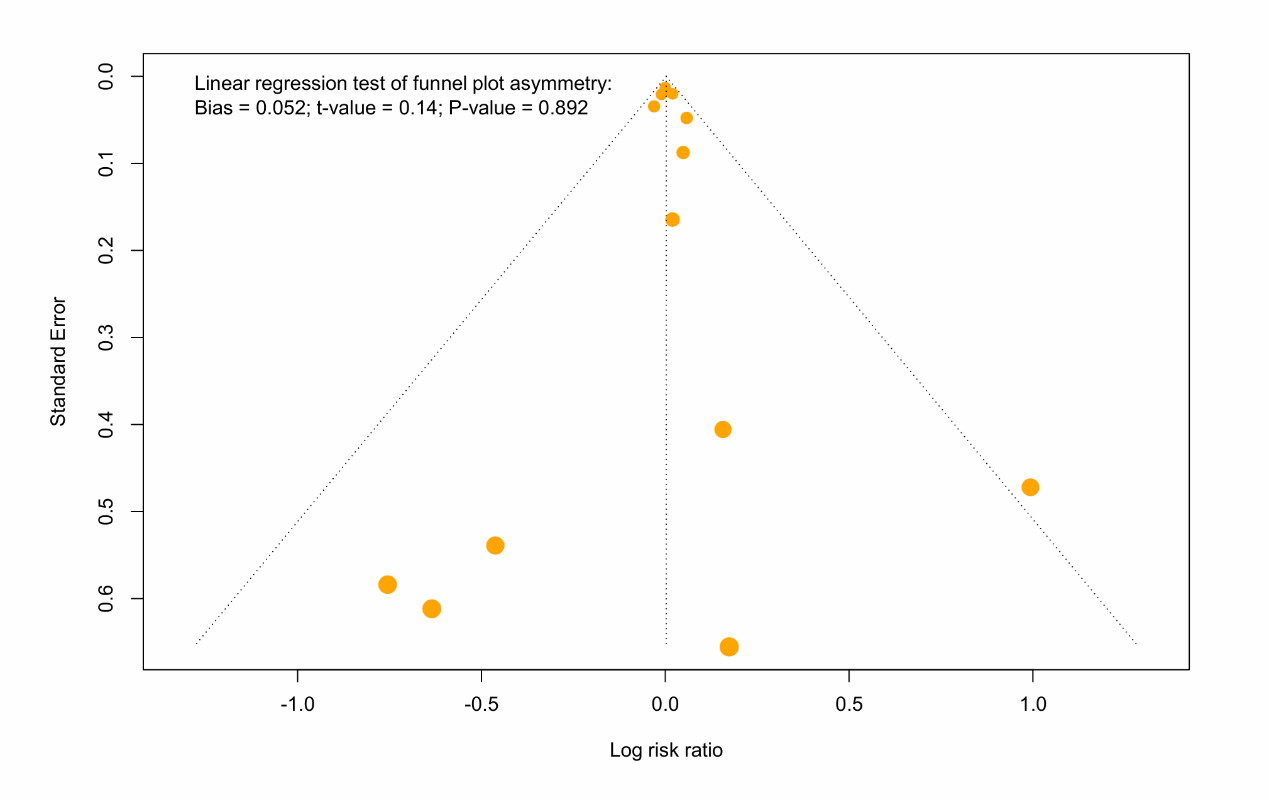

Supplementary Figure 4-2: Forest plot of 5oxo-MEHP exposure on the risk of early puberty (combined).

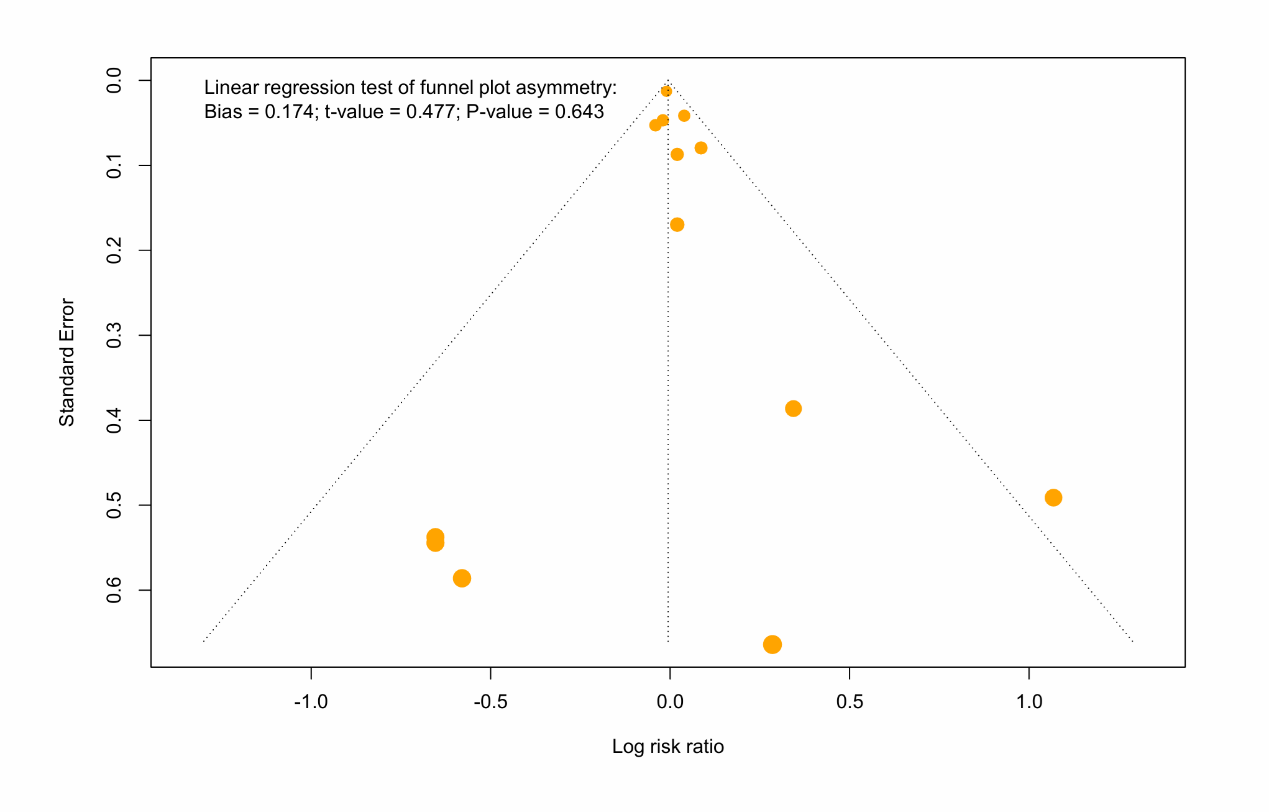

Supplementary Figure 4-3: Funnel plot of MBzP exposure on the risk of early puberty (combined).

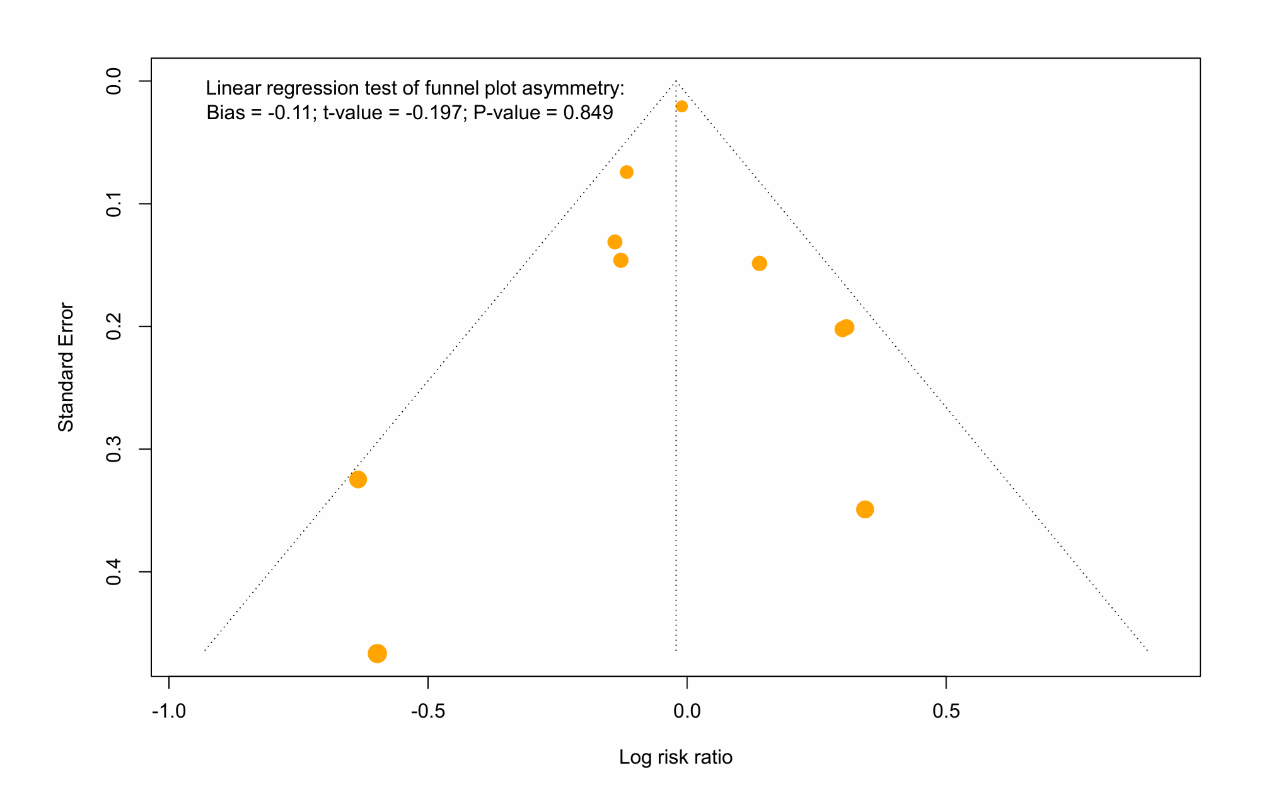

Supplementary Figure 4-4: Funnel plot of MEHP exposure on the risk of early puberty (combined).

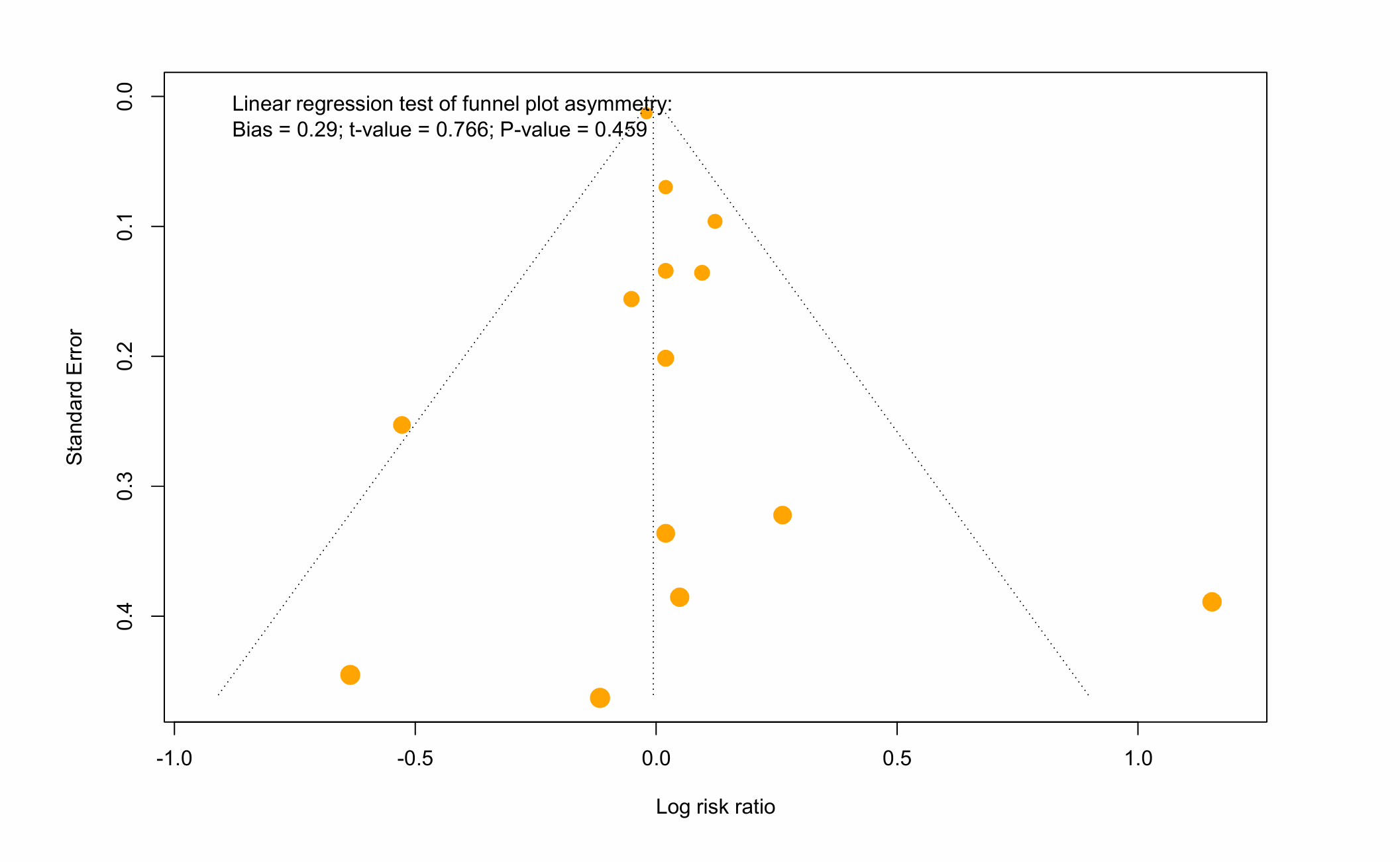

Supplementary Figure 4-5: Funnel plot of studies of MEP exposure on the risk of early puberty (combined).

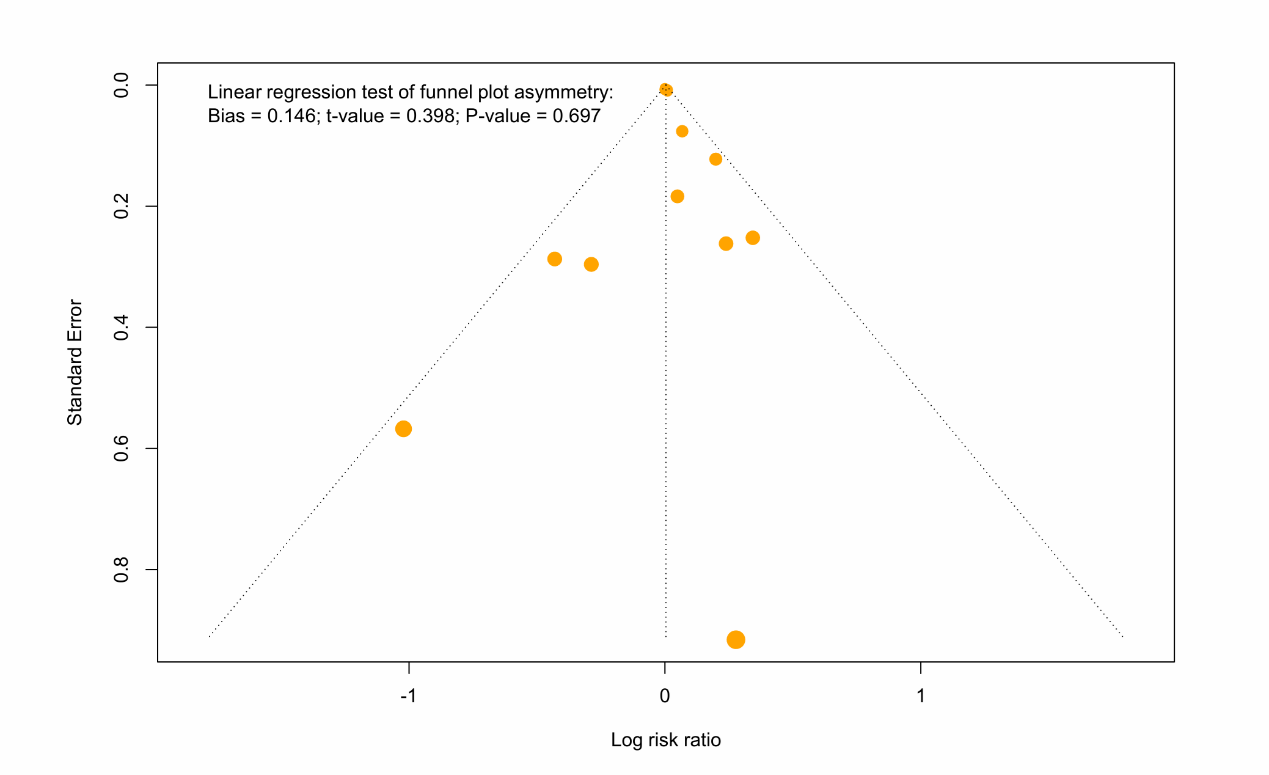

Supplementary Figure 4-6: Funnel plot of studies demonstrating the RR of MnBP exposure for early puberty (combined).

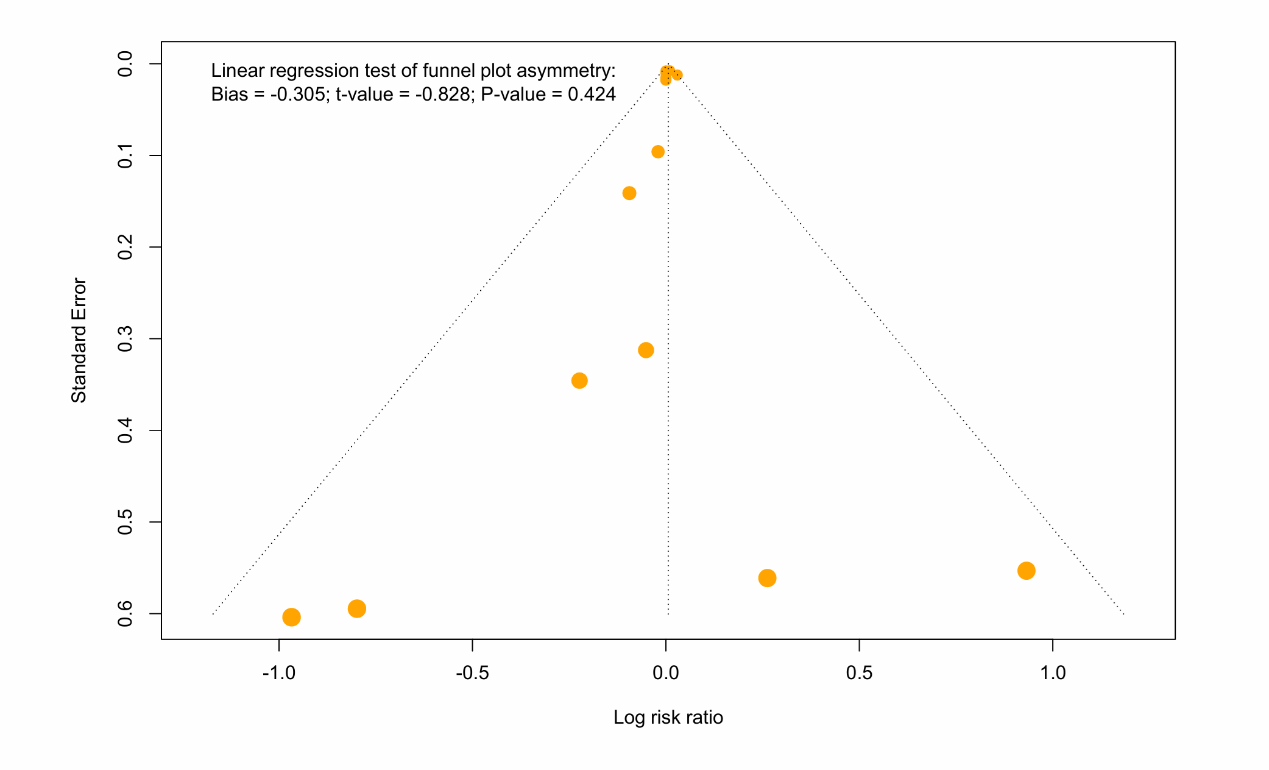

Supplementary Figure 4-7: Funnel plot of studies demonstrating the publication bias of MBzP exposure (unadjusted) for early puberty.

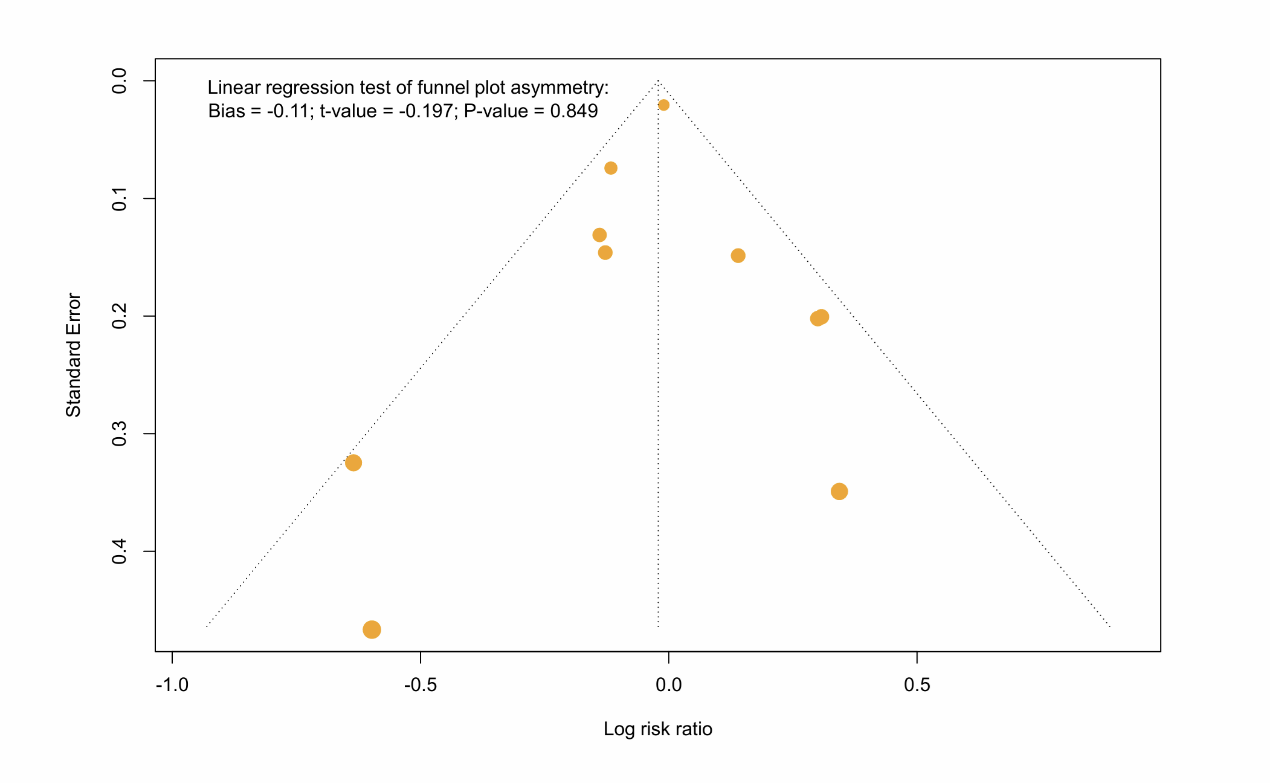

Supplementary Figure 4-8: Funnel plot of studies demonstrating the publication bias of MEP exposure (unadjusted) on the risk of early puberty.

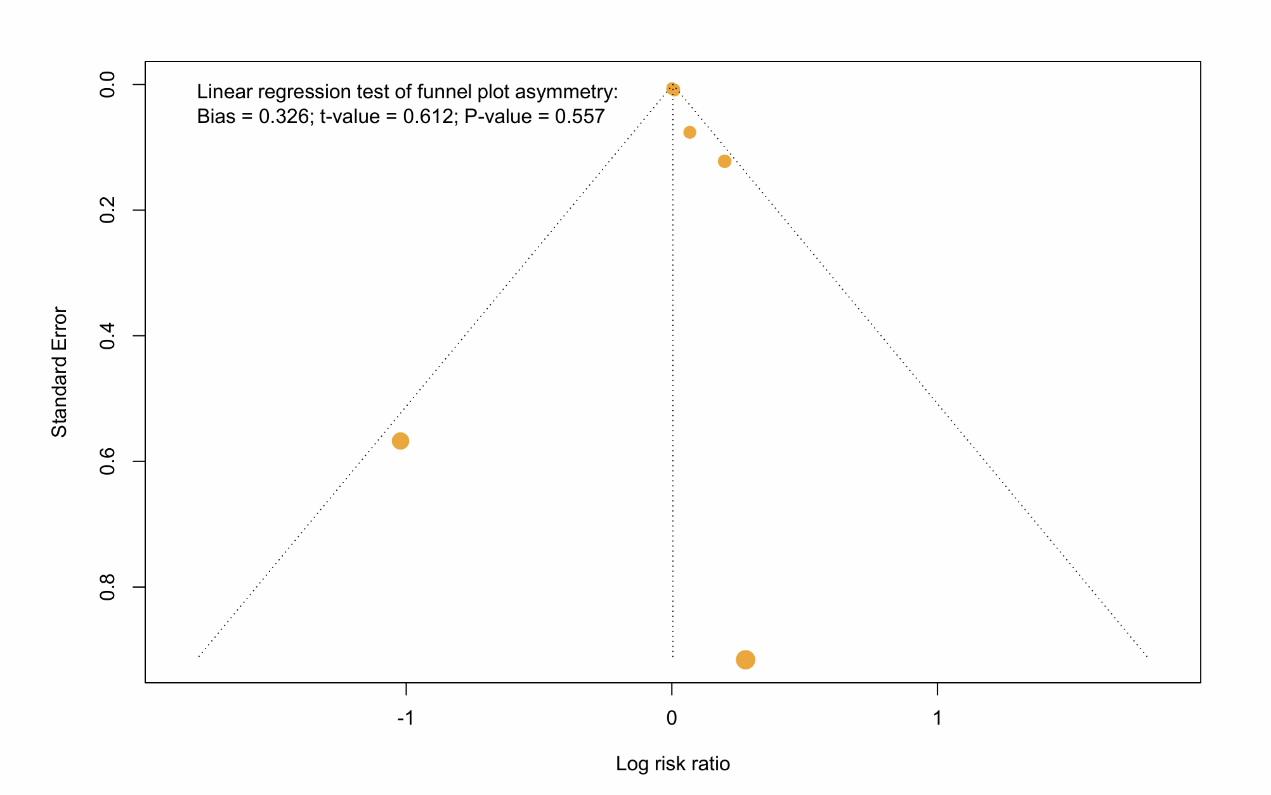

### **Supplementary Figure 5 (5-1~5-10) : Forest plots (outcome of thelarche)**

Abbreviations:

5cx-MEPP, mono(2-ethyl-5-carboxypentyl)phthalate; 5OH-MEHP, mono(2-ethyl-5-hydroxyhexyl) phthalate; 5oxo-MEHP, mono(2-ethyl-5-oxohexyl)phthalate; MBzP, monobenzyl phthalate; MCPP, mono-(3-carboxypropyl) phthalate; MEHP, mono(2-ethylhexyl) phthalate; MEP, monoethyl phthalate; MiBP, mono-isobutyl phthalate; MMP, monomethyl phthalate; MnBP, mono-n-butyl phthalate

Supplementary Figure 5-1: Forest plot of studies demonstrating the RR of 5cx-MEPP exposure for early puberty (thelarche), stratified by exposure time.

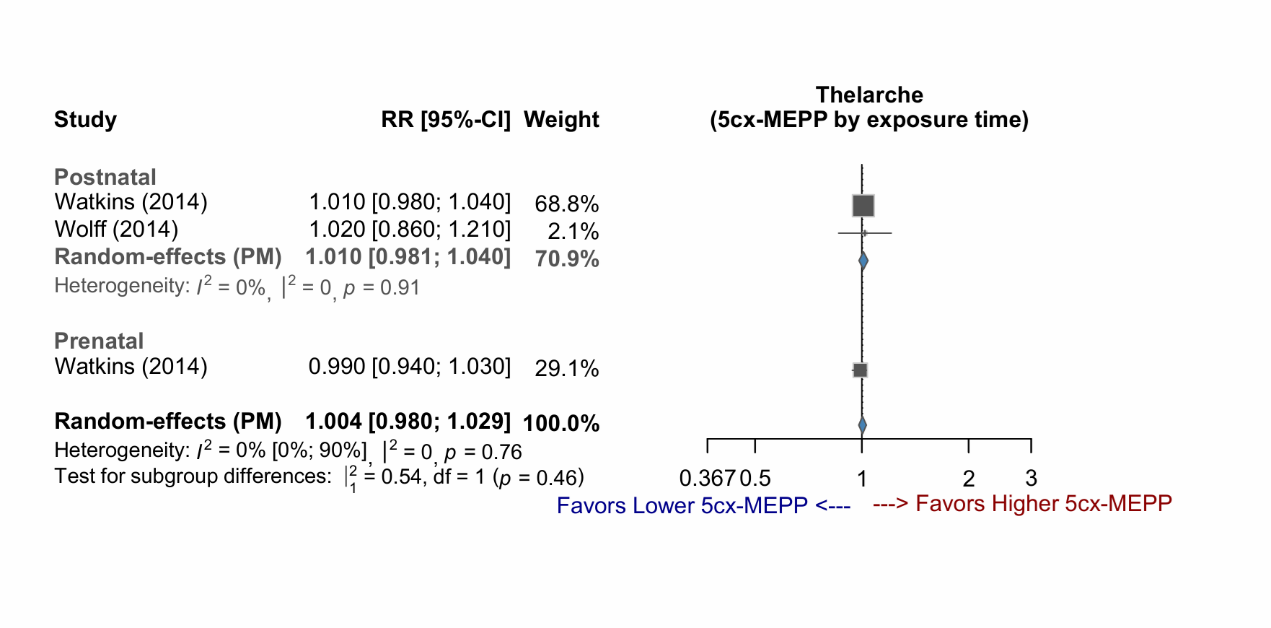

Supplementary Figure 5-2: Forest plot of studies demonstrating the RR of 5OH-MEHP exposure for early puberty (thelarche), stratified by exposure time.

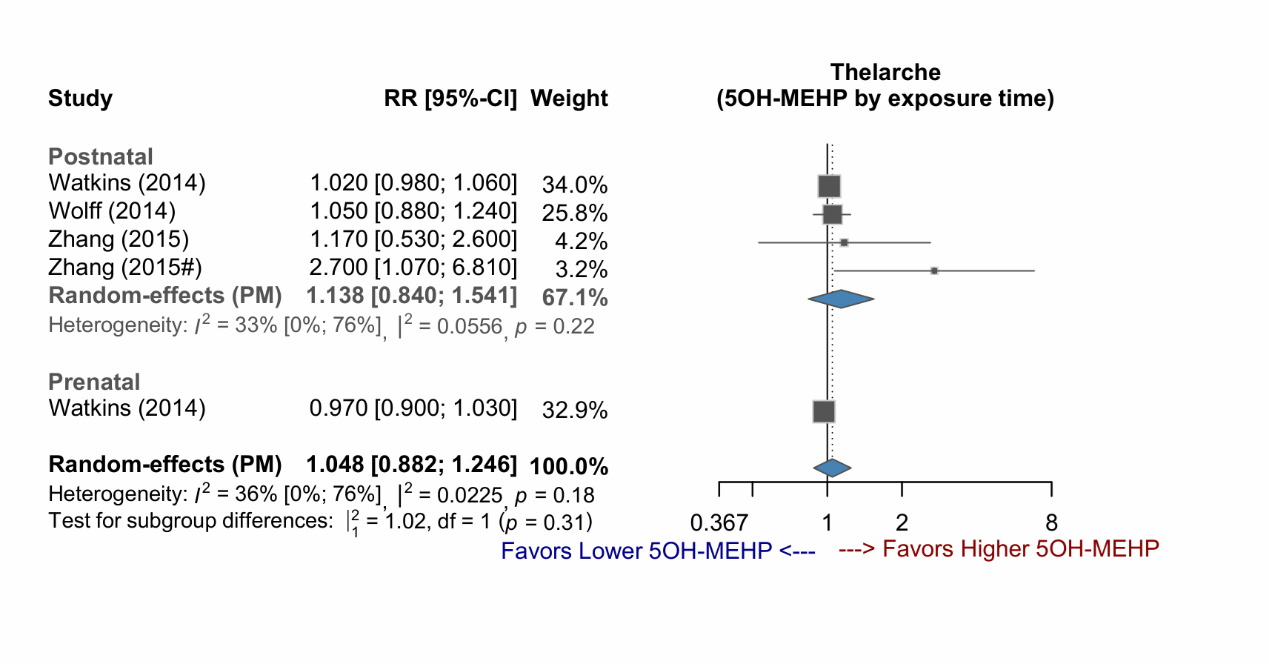

Supplementary Figure 5-3: Forest plot of studies demonstrating the RR of 5oxo-MEHP exposure for early puberty (thelarche), stratified by exposure time.

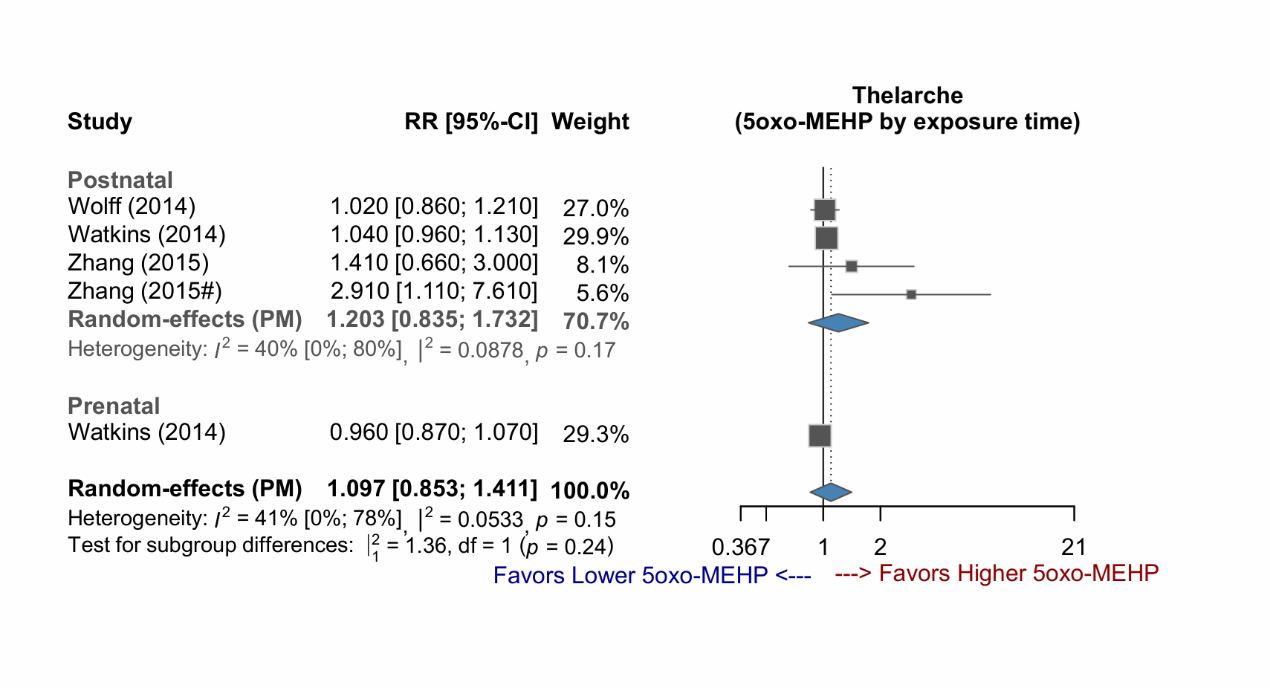

Supplementary Figure 5-4: Forest plot of studies demonstrating the RR of MBzP exposure for early puberty (thelarche), stratified by exposure time.

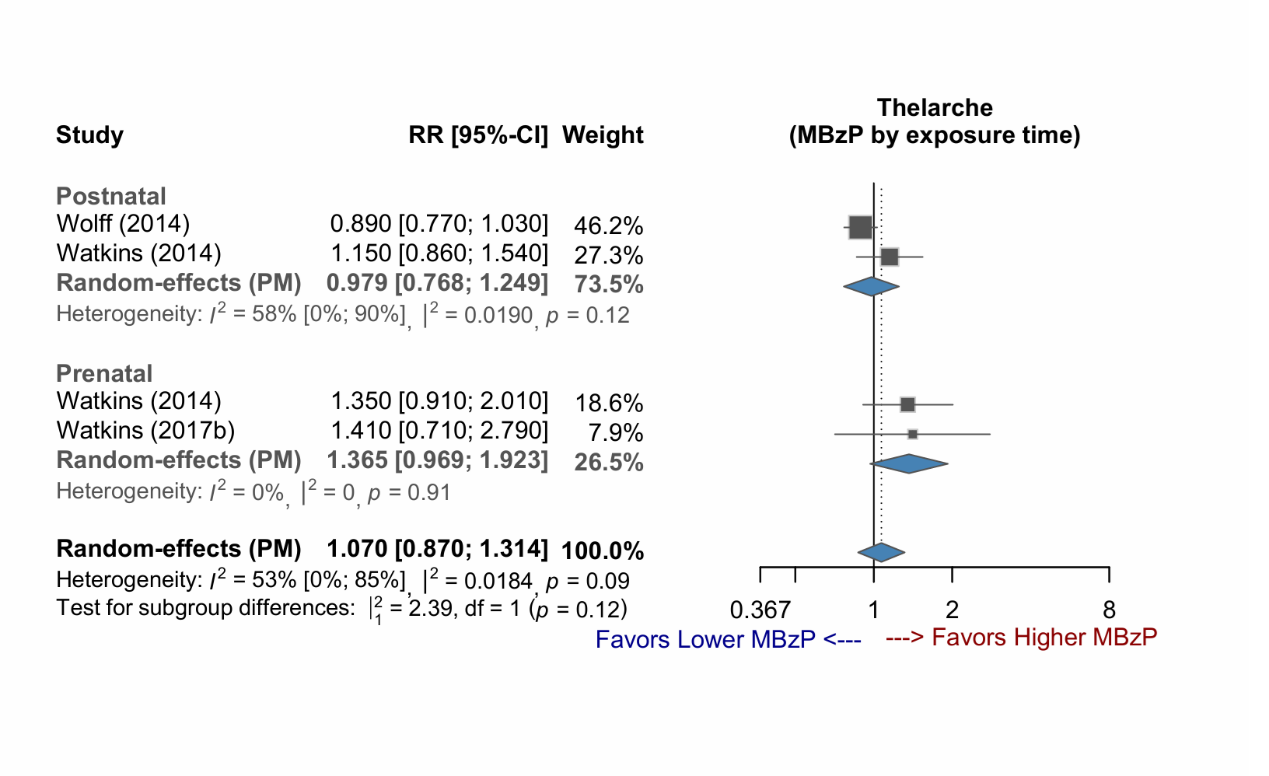

Supplementary Figure 5-5: Forest plot of studies demonstrating the RR of MCPP exposure for early puberty (thelarche), stratified by exposure time.

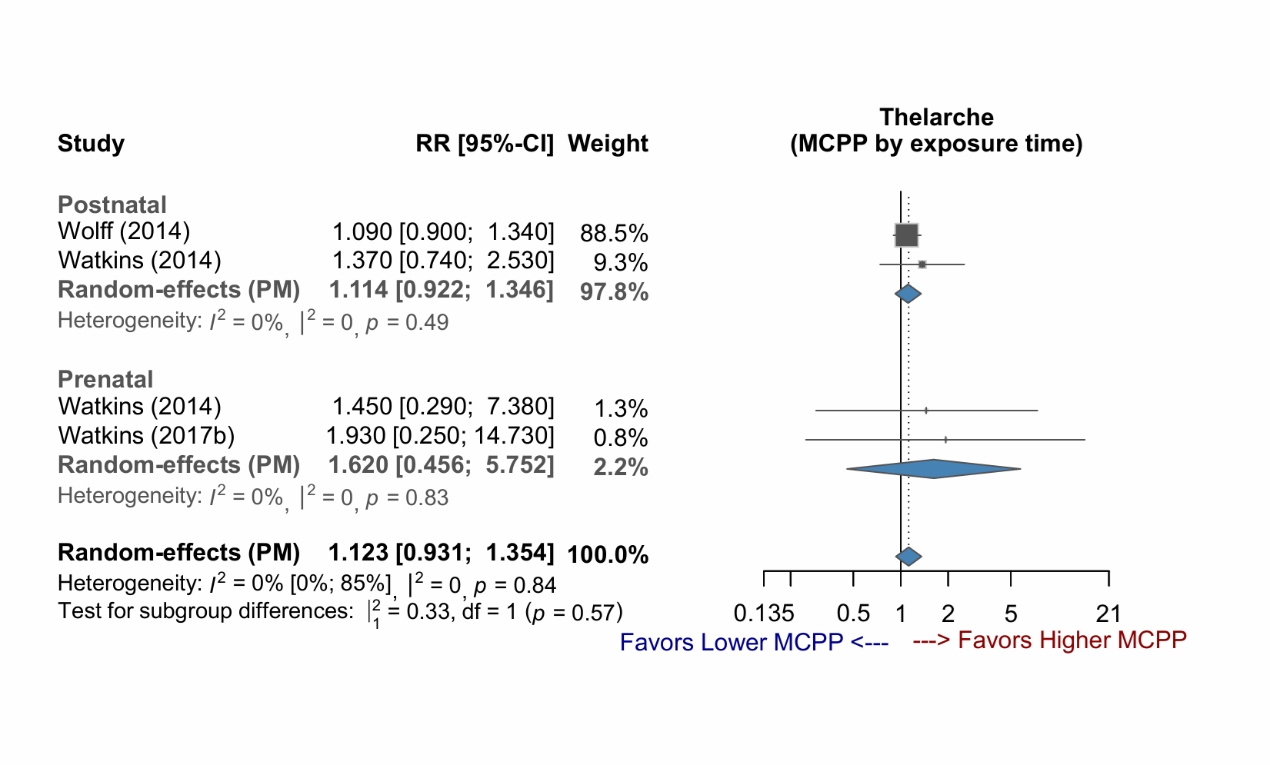

Supplementary Figure 5-6: Forest plot of studies demonstrating the RR of MEHP exposure for early puberty (thelarche), stratified by exposure time.

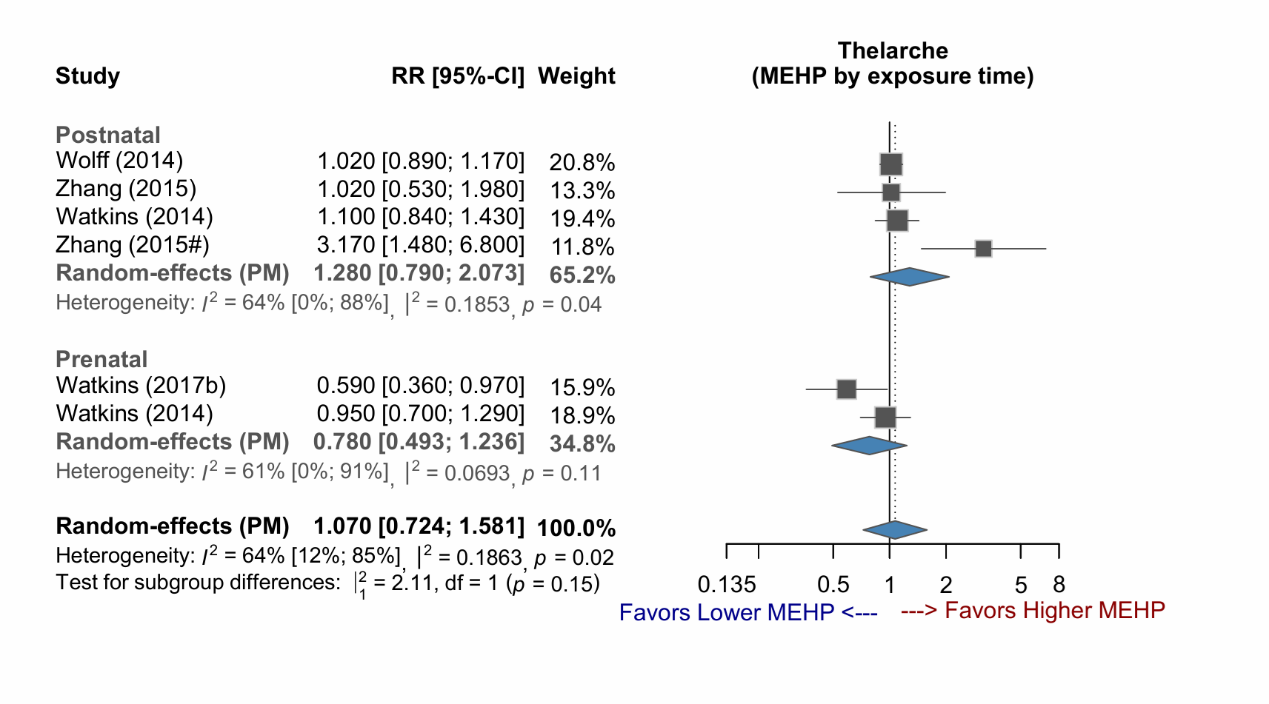

Supplementary Figure 5-7: Forest plot of studies demonstrating the RR of MEP exposure for early puberty (thelarche), stratified by exposure time.

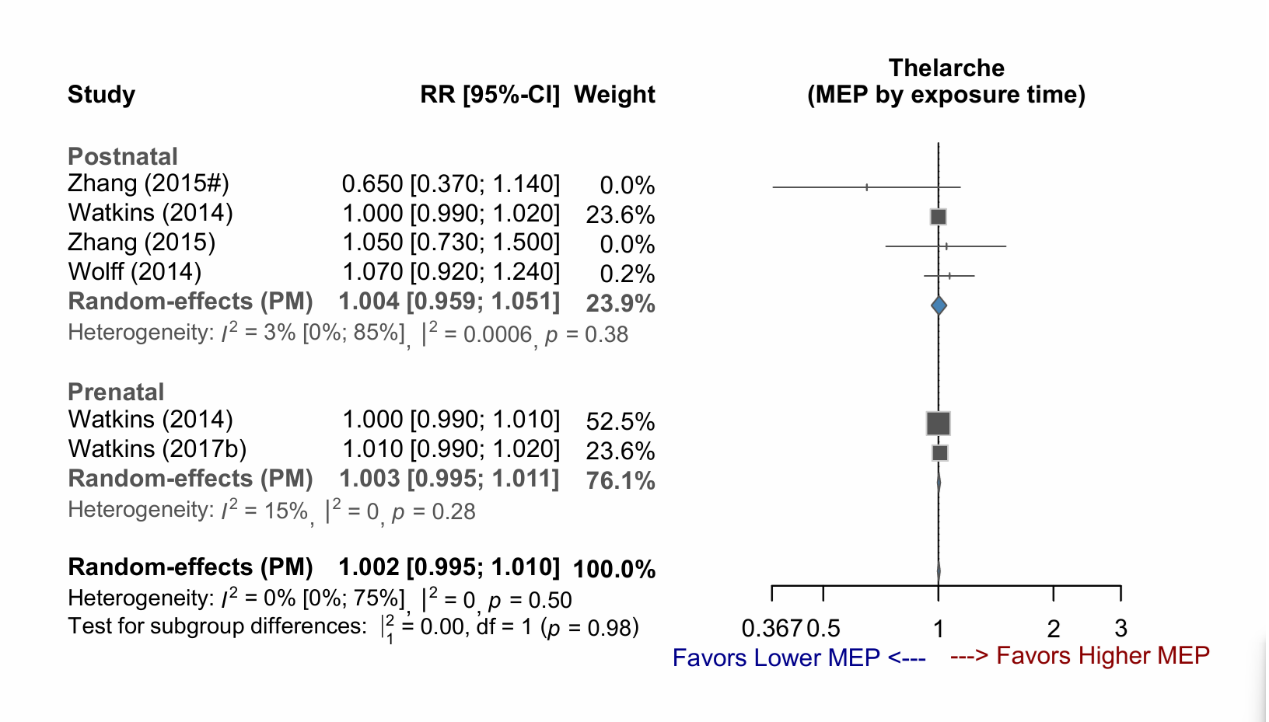

Supplementary Figure 5-8: Forest plot of studies demonstrating the RR of MiBP exposure for early puberty (thelarche), stratified by exposure time.

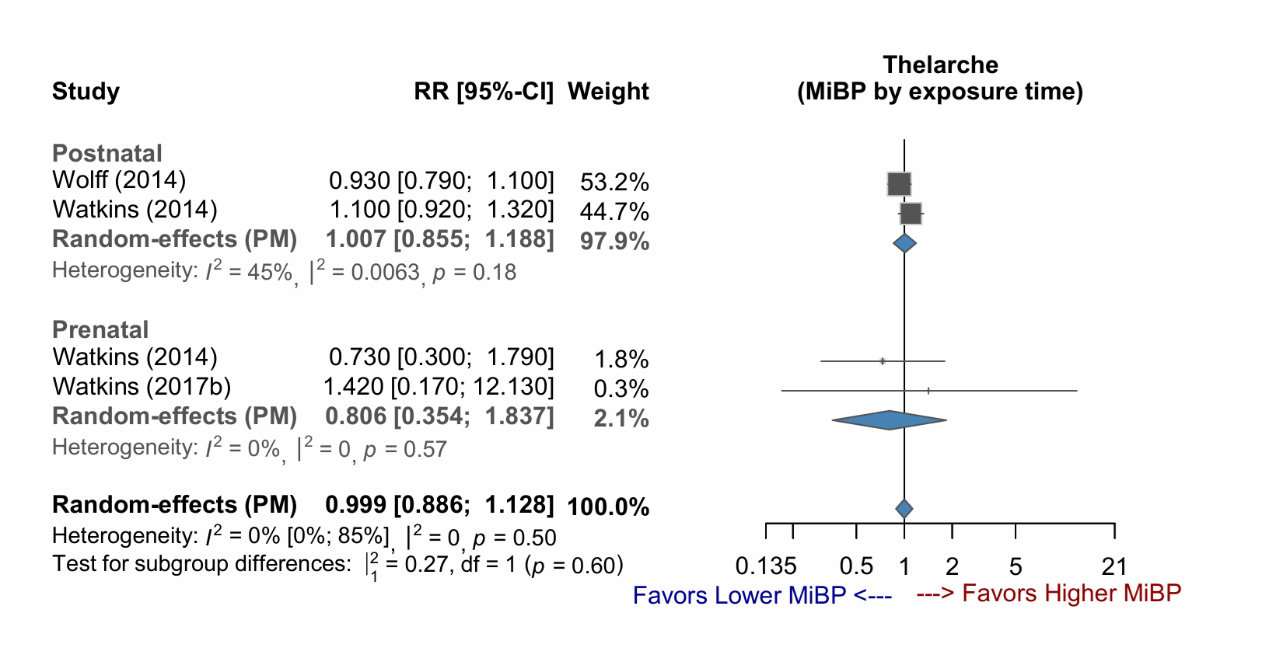

Supplementary Figure 5-9: Forest plot of studies demonstrating the RR of MMP exposure for early puberty (thelarche).

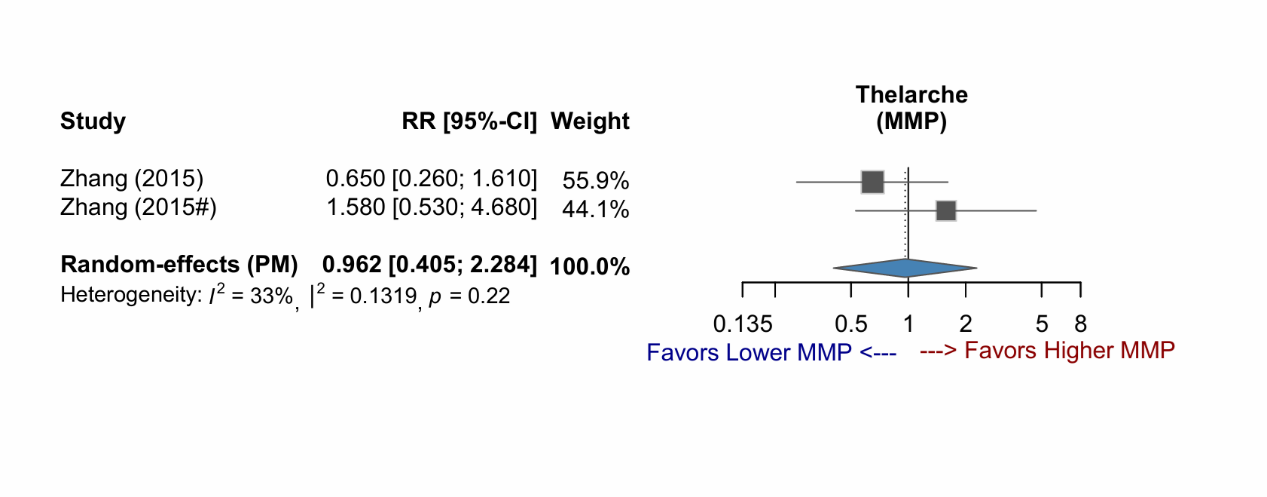

Supplementary Figure 5-10: Forest plot of studies demonstrating the RR of MnBP exposure for early puberty (thelarche), stratified by exposure time.

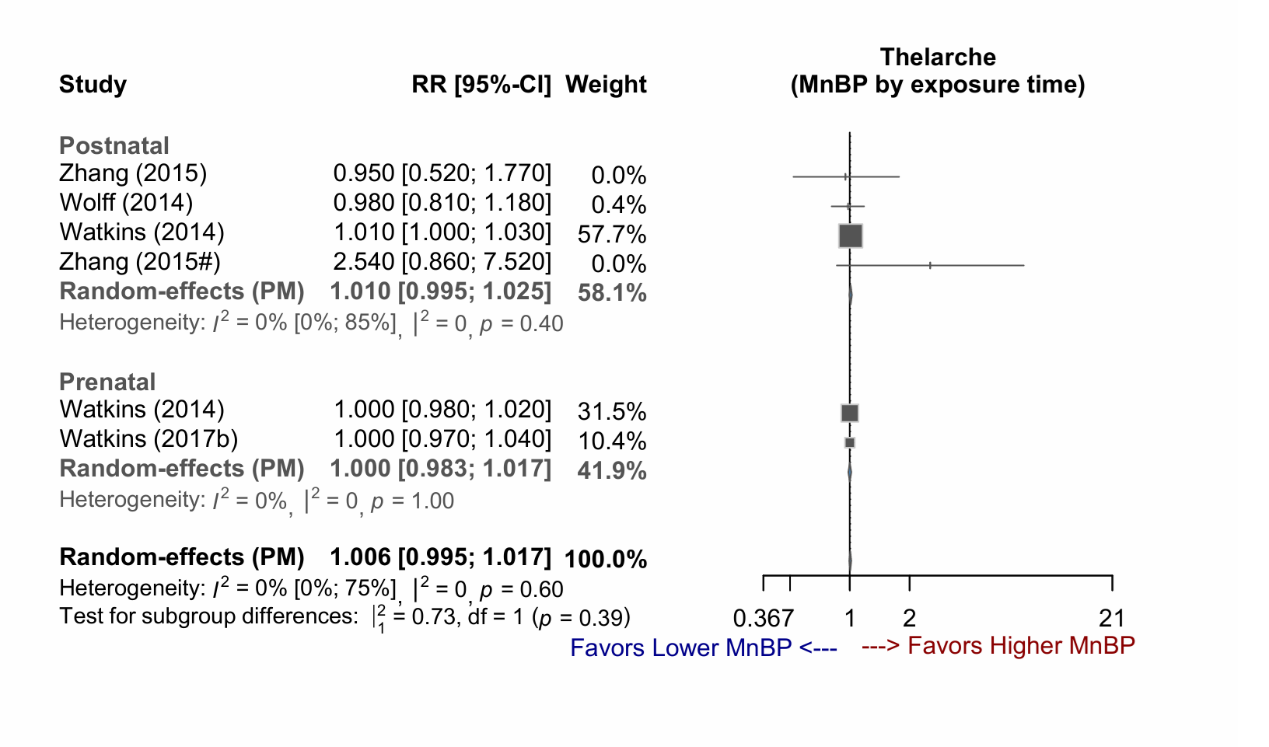

### **Supplementary Figure 6 (6-1~6-13) : Forest plots (outcome of menarche)**

Abbreviations:

2OH-MiBP, 2OH-mono-isobutyl phthalate; 3OH-MnBP, 3OH-mono-n-butyl phthalate; 5cx-MEPP, mono(2-ethyl-5-carboxypentyl)phthalate; 5OH-MEHP, mono(2-ethyl-5-hydroxyhexyl) phthalate; 5oxo-MEHP, mono(2-ethyl-5-oxohexyl)phthalate; cx-MiOP, monocarboxy isooctyl phthalate; MBzP, monobenzyl phthalate; MCPP, mono-(3-carboxypropyl) phthalate; MEHP, mono(2-ethylhexyl) phthalate; MEP, monoethyl phthalate; MiBP, mono-isobutyl phthalate; MMP, monomethyl phthalate; MnBP, mono-n-butyl phthalate

Supplementary Figure 6-1: Forest plot of studies demonstrating the RR of 2OH-MiBP exposure for early puberty (menarche).

Supplementary Figure 6-2: Forest plot of studies demonstrating the RR of 3OH-MnBP exposure for early puberty (menarche).

Supplementary Figure 6-3: Forest plot of studies demonstrating the RR of 5cx-MEPP exposure for early puberty (menarche).

Supplementary Figure 6-4: Forest plot of studies demonstrating the RR of 5OH-MEHP exposure for early puberty (menarche).

Supplementary Figure 6-5: Forest plot of studies demonstrating the RR of 5oxo-MEHP exposure for early puberty (menarche).

Supplementary Figure 6-6: Forest plot of studies demonstrating the RR of cx-MiOP exposure for early puberty (menarche).

Supplementary Figure 6-7: Forest plot of studies demonstrating the RR of MBzP exposure for early puberty (menarche).

Supplementary Figure 6-8: Forest plot of studies demonstrating the RR of MCPP exposure for early puberty (menarche).

Supplementary Figure 6-9: Forest plot of studies demonstrating the RR of MEHP exposure for early puberty (menarche).

Supplementary Figure 6-10: Forest plot of studies demonstrating the RR of MEP exposure for early puberty (menarche).

Supplementary Figure 6-11: Forest plot of studies demonstrating the RR of MiBP exposure for early puberty (menarche).

Supplementary Figure 6-12: Forest plot of studies demonstrating the RR of MMP exposure for early puberty (menarche).

Supplementary Figure 6-13: Forest plot of studies demonstrating the RR of MnBP exposure for early puberty (menarche).

### **Supplementary Figure 7 (7-1~7-10) : Forest plots (outcome of testicular volume)**

Abbreviations:

5cx-MEPP, mono(2-ethyl-5-carboxypentyl)phthalate; 5OH-MEHP, mono(2-ethyl-5-hydroxyhexyl) phthalate; 5oxo-MEHP, mono(2-ethyl-5-oxohexyl)phthalate; MBzP, monobenzyl phthalate; MCPP, mono-(3-carboxypropyl) phthalate; MEHP, mono(2-ethylhexyl) phthalate; MEP, monoethyl phthalate; MiBP, mono-isobutyl phthalate; MMP, monomethyl phthalate; MnBP, mono-n-butyl phthalate

Supplementary Figure 7-1: Forest plot of studies demonstrating the RR of 5cx-MEPP exposure for early puberty (testicular volume), stratified by exposure time.

Supplementary Figure 7-2: Forest plot of studies demonstrating the RR of 5OH-MEHP exposure for early puberty (testicular volume), stratified by exposure time.

Supplementary Figure 7-3: Forest plot of studies demonstrating the RR of 5oxo-MEHP exposure for early puberty (testicular volume), stratified by exposure time.

Supplementary Figure 7-4: Forest plot of studies demonstrating the RR of MBzP exposure for early puberty (testicular volume), stratified by exposure time.

Supplementary Figure 7-5: Forest plot of studies demonstrating the RR of MCPP exposure for early puberty (testicular volume), stratified by exposure time.

Supplementary Figure 7-6: Forest plot of studies demonstrating the RR of MEHP exposure for early puberty (testicular volume), stratified by exposure time.

Supplementary Figure 7-7: Forest plot of studies demonstrating the RR of MEP exposure for early puberty (testicular volume), stratified by exposure time.

Supplementary Figure 7-8: Forest plot of studies demonstrating the RR of MiBP exposure for early puberty (testicular volume), stratified by exposure time.

Supplementary Figure 7-9: Forest plot of studies demonstrating the RR of MMP exposure for early puberty (testicular volume).

Supplementary Figure 7-10: Forest plot of studies demonstrating the RR of MnBP exposure for early puberty (testicular volume), stratified by exposure time.
